# Risk factors in the first 1000 days of life associated with childhood obesity: a systematic review and risk factor quality assessment

**DOI:** 10.1101/2024.06.26.24309514

**Authors:** Sophia M Blaauwendraad, Arwen SJ Kamphuis, Francisco Javier Ruiz-Ojeda, Marco Brandimonte-Hernández, Eduard Flores-Ventura, Marieke Abrahamse-Berkeveld, Maria Carmen Collado, Janna A van Diepen, Patricia Iozzo, Karen Knipping, Carolien A van Loo-Bouwman, Ángel Gil, Romy Gaillard

## Abstract

**Background:** Adverse early life exposures might negatively affect foetal and infant development, predisposing children to obesity. We aimed to systematically identify and evaluate risk factors for childhood obesity in preconception, pregnancy, and infancy, and assess their potential as targets for future prediction and prevention strategies.

**Methods:** This systematic review (PROSPERO, CRD42022355152) included longitudinal studies from selected electronic databases published between inception and August 17^th^, 2022, identifying maternal, paternal, or infant risk factors from preconception until infancy for childhood obesity between 2 and 18 years. Screening and data extraction was performed through standardized extraction forms. We assessed risk factor quality on modifiability and predictive power using a piloted criteria template from ILSI-Europe-Marker-Validation-Initiative.

**Findings:** We identified 172 observational and 5 intervention studies involving n=1.879.971 children from 37, predominantly high-income, countries. 59%, 25% and 16% of studies measured childhood obesity between 2- <6 years, 6-10 years, and >10 -18 years respectively. Average reported childhood obesity prevalence was 11·1%. Pregnancy and infancy risk factors were mostly studied. We identified 59 potential risk factors, 24 of which were consistently associated with childhood obesity risk. Higher maternal prepregnancy weight (n=28/31 positive associations from 31 studies, respectively), higher gestational weight gain (n=18/21), maternal smoking during pregnancy (n=23/29), higher birth weight (n=20/28), LGA (n=17/18), no breastfeeding (n=20/31), and higher infant weight gain (n=12/12) were the strongest risk factors, which may aid in prediction or be targets for prevention. Level of evidence was generally moderate due to unreliable exposure measurement, short follow-up/loss-to-follow up, and risk of confounding.

**Interpretation:** We identified 7 early life risk factors, which were strongly associated with a higher risk of childhood obesity, and can contribute to future prediction and prevention strategies. These findings support implementation of prevention strategies targeting these early-life risk factors from a clinical and population perspective, where possible integrated with implementation studies.

**Funding:** This work was conducted by an expert group of the European branch of the International Life Sciences Institute, ILSI Europe.

## INTRODUCTION

Childhood obesity is a major public health challenge.^1^ Childhood obesity rates have rapidly increased over the past decades and continue to rise worldwide.^2^ Childhood obesity is related to increased risks of non-communicable diseases, including hypertension, diabetes, depression from childhood onwards and premature mortality in adulthood, leads to a reduced quality of life and large economic and societal burden .^3–5^

Accumulating evidence suggests that high susceptibility to obesity might already originate in early life.^6,7^ The Development Origins of Health and Disease (DOHaD) hypothesis states that in the early stages of human development, individuals are particularly vulnerable to an adverse environment.^8,9^ During preconception, egg cell maturation and sperm production occur. After conception, foetal life and early infancy are characterized by rapid cell division, organ development, and rapid growth. These stages define the blueprint of later health and disease; thus, adverse exposures during these critical developmental periods might predispose an individual to an increased risk of obesity in later life. As the prevalence of childhood obesity continues to rise, systematic identification of early-life risk factors for childhood obesity is urgently needed to enable development of improved prevention strategies with better early prediction of childhood obesity risk and potential novel modifiable targets for interventions at an individual and population level.

Therefore, we conducted a systematic review to first identify risk factors for childhood obesity in the preconception period, pregnancy, and infancy, thereby covering the critical first 1000 days of life. Second, we aimed to assess quality of potential crucial early-life risk factors for prediction of childhood obesity risk and as potential modifiable targets for future prevention strategies.

## METHODS

### Systematic review protocol development

This study was part of a large collaborative effort to perform a systematic review of risk factors in the first 1000 days of life for the development of various childhood cardiometabolic disorders. We developed a systematic review protocol to comprehensively include and evaluate individual research studies reporting on risk factors and noninvasive biomarkers during preconception, pregnancy and infancy for the development of various child and adolescent cardiometabolic disorders. For this study, we aimed to identify longitudinal observational or intervention studies that focused on association studies or prediction studies for early risk factors for childhood obesity between 2 and 18 years.

Risk factors and noninvasive biomarkers of interest, hereafter also referred to as “risk factors,” included sociodemographic factors, lifestyle factors, physical factors, environmental factors, pregnancy-related factors, and noninvasive biomarkers. We were interested in risk factors in mothers, fathers and offspring obtained during the crucial periods, preconception, pregnancy, and infancy until 2 years, together covering the first 1000 days of life. The outcome of interest was self-reported, physician or researcher-diagnosed obesity.

### Data sources, search strategy, and screening criteria

We registered our search strategy and systematic review protocol to PROSPERO CRD42022355152. We developed search terms for Medline, EMBASE, Web of Science, SCOPUS and Cochrane CENTRAL (**Text S1**) for eligible citations published in English language through August 17^th^, 2022. We included prospective and retrospective longitudinal observational studies identifying factors of incident offspring outcomes of interest. We excluded cross-sectional studies and studies among diseased populations only. We also included interventions prospectively comparing treatment effects on the outcome. Exclusion criteria comprised studies with outcomes before the age of 2 years, studies reporting only intermediate phenotypes or continuous traits, or studies that only assessed endpoints outside of the cardiometabolic outcomes of interest. 2 independent reviewers screened at the title and abstract level. For accepted citations, 2 independent reviewers screened the full manuscripts. Conflicts at all screening stages were resolved by a third reviewer. Conflicts that were not resolved by the third reviewer were discussed in the full group. All screenings were conducted in the Covidence online systematic review tracking platform.

### Data extraction and synthesis of results

We developed and piloted a data extraction template for eligible manuscripts, which included manuscript information, study level details and design, population enrolment and characteristics, exposure and outcome ascertainment and diagnosis criteria, and age at offspring outcome assessment. We classified exposures in 8 broad categories: (i) parental lifestyle factors, (ii) parental physical factors, (iii) environmental factors, (iv) pregnancy-related factors,(v) birth anthropometrics,(vi) feeding patterns, (vii) infant anthropometrics, and (viii) cord blood biomarkers.

### Risk of bias assessment for quality and synthesis

We assessed quality of each study using the Joanna Briggs Institute (JBI) critical appraisal tools for cohort studies, case-control studies, and randomized controlled trials (RCTs).^10^ For cohort and case-control studies, we assessed quality based on 11 and 9 items, respectively that evaluated population recruitment, exposure and outcome ascertainment, confounding, statistical methodology and follow-up. For the RCTs, we evaluated 11 items that assessed selection and allocation, intervention, administration, outcome ascertainment, follow-up, and statistical analysis. JBI items were categorized as, ‘Yes,’ ‘No,’ ‘Unclear,’ or ‘Not applicable’ following established guidelines. Any uncertainty in assessment was further discussed by the full research team. Studies with overall scores of 50% or less on questions answered with ‘Yes’, 50% until 70% questions answered with ‘Yes’, and 70% or more questions answered with ‘Yes’, were considered low, moderate and high quality, respectively.

### Quality assessment of risk factors for prediction and prevention

We took forward those risk factors we considered consistently associated with childhood obesity for quality assessment. We defined a consistently associated risk factor as a risk factor for which over 50% of studies reported an association in the same direction, in at least 2 studies of moderate or high quality. We developed and piloted a criteria template for risk factor quality assessment based on the ILSI Europe Marker Validation Initiative (**Table S1)**.^11^ Early life risk factors were scored on their methodological aspects, reflection of the study objective, prediction, and modifiability. Risk factors were scored by 3 independent reviewers at 4 different levels based on the criteria: very strong, strong, medium, and low. Conflicts were discussed in the full group until consensus was reached. Risk factors, which scored strong or very strong on prediction (65% of studies reported an association in the same direction in at least five moderate or high quality studies; >80% of studies reported an association in the same direction in at least five moderate or high quality studies, respectively) and strong or very strong on modifiability were considered to be most the most useful risk factors for prediction and prevention strategies. Modifiability was based on two components: theoretically modifiable and findings from intervention studies. If a risk factor scored strong or very strong on theoretically modifiable (risk factor is modifiable on individual level and implementation of modifications in daily life is complicated or easy, respectively) and low on intervention studies (No intervention studies have been conducted, or no potential effect of modification has been found), it was considered strong. Very strong on modifiability was considered if a risk factor scored strong or very strong on theoretically modifiable scored and at least medium on intervention studies (There is some evidence of potential effect of intervention on the outcome, but literature is controversial, or higher respectively).

## RESULTS

### Study selection and participant characteristics

**Figure 1** shows 35584 publications were identified for screening through searches of selected electronic databases. After removing 17974 duplicates, 17610 publications were subjected to title and abstract screening by 2 independent reviewers. 16059 publications were found to be irrelevant, and 1551 publications underwent full text screening. 177 studies met the inclusion criteria and were included in this review.

**Figure 1.**
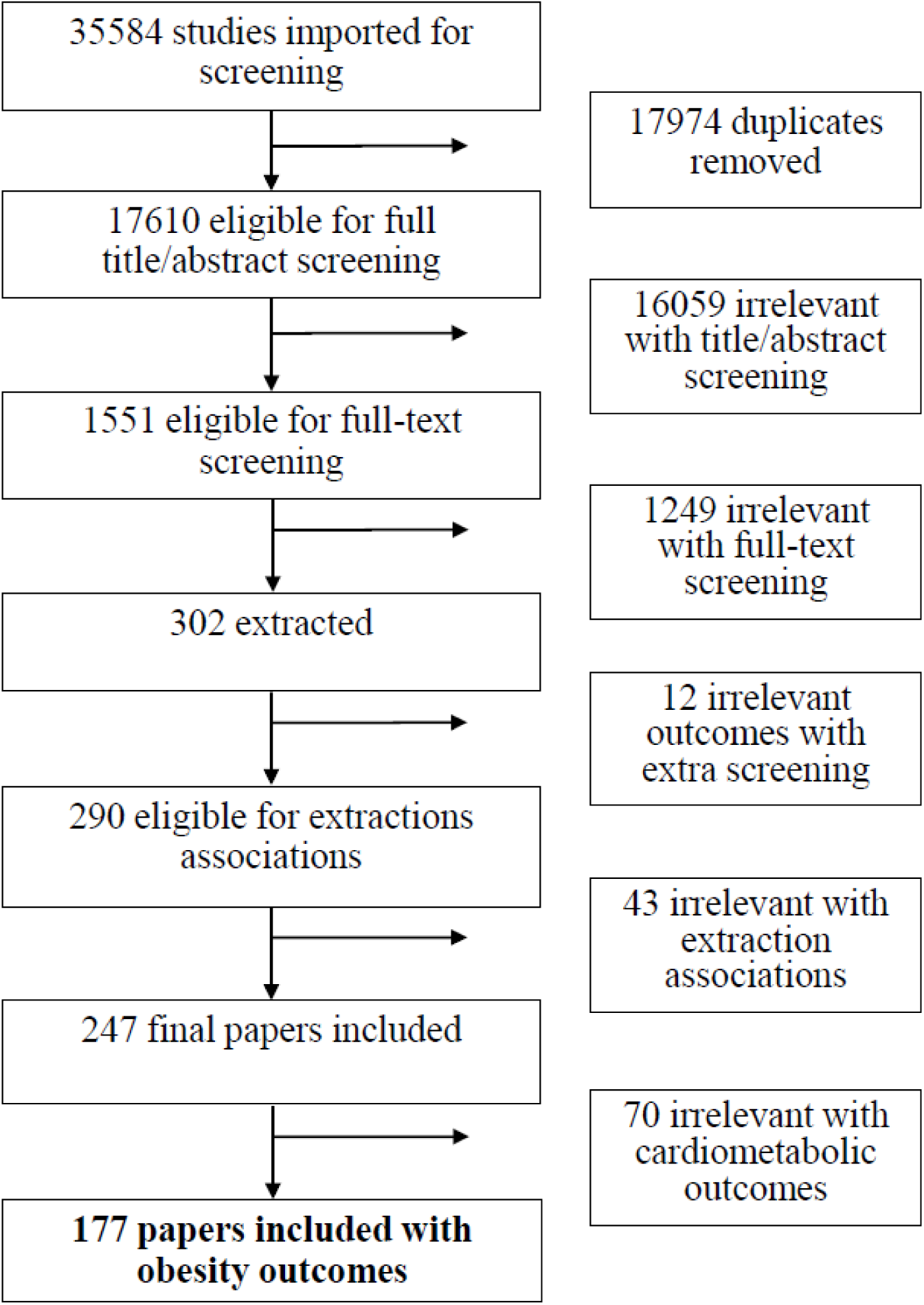
Flowchart of studies included in systematic review

165 were observational studies, 7 were case-control studies and 5 were intervention studies (**Table 1).** Studies were performed between 1970 and 2022. Sample sizes varied from 50 to 155411 participants, leading to 1879971 children included in the final study sample. 59% of studies measured childhood obesity between 2 to 6 years, 25% between 6 to 10 years, and 16% between 10 and 18 years. **Figure 2A** shows that studies included data from 37 different countries, predominantly high-income countries. The average reported prevalence of obesity was 11·1%, which ranged from 0·9% in Peru to 19·6% in Finland **(Figure 2B).**

**Figure 2.**
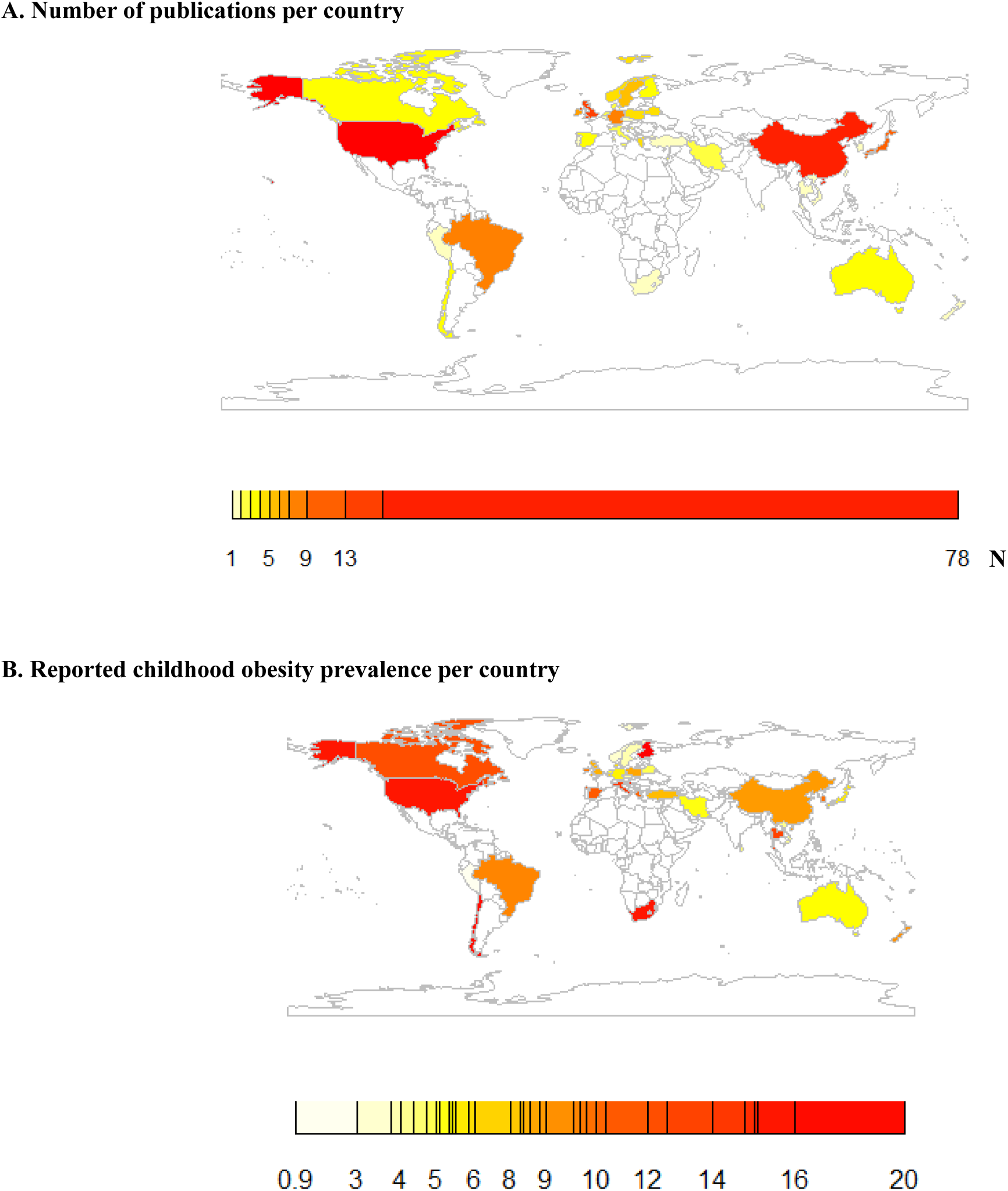
**World map of the included studies**

**Table 1.**
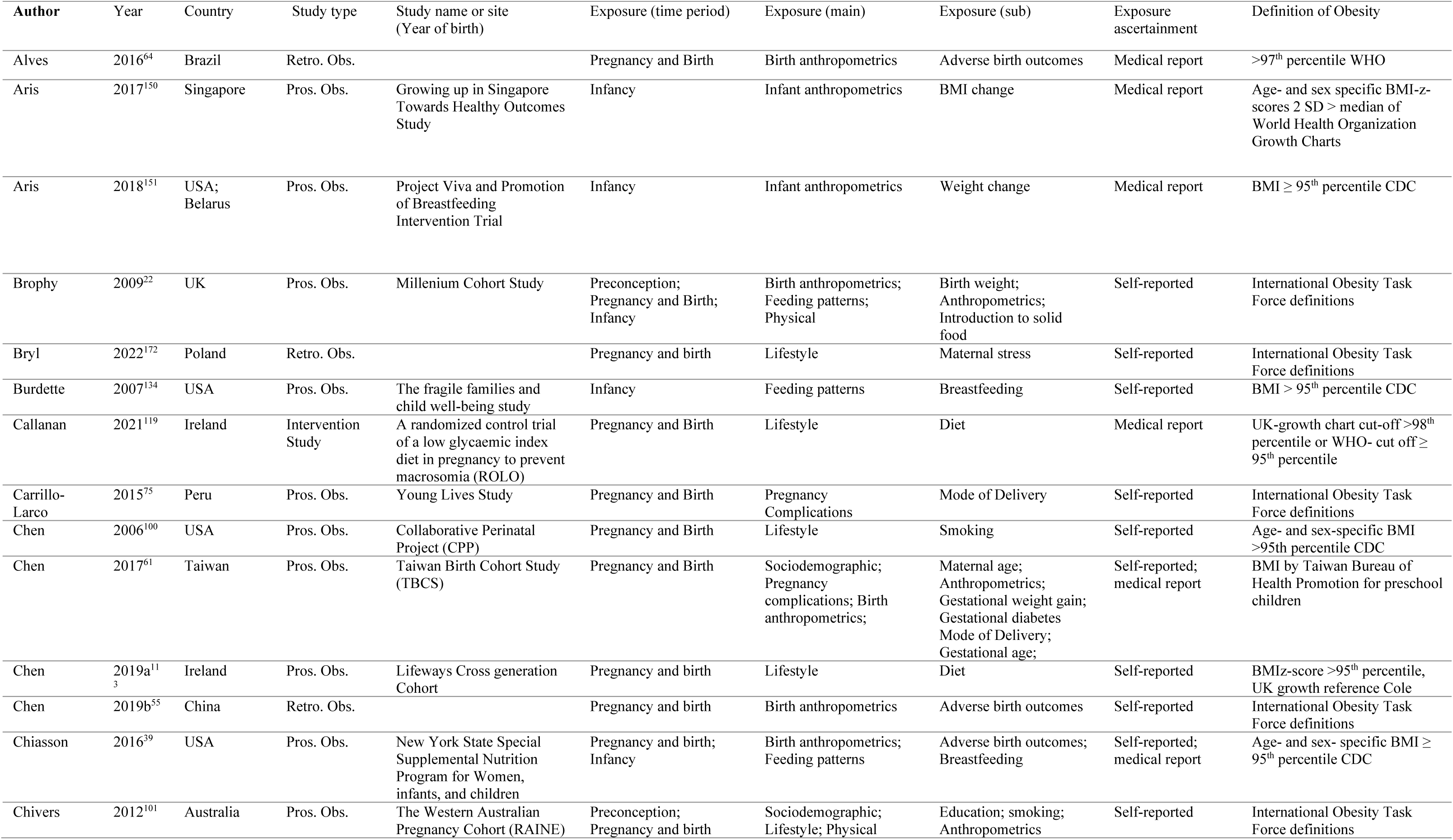
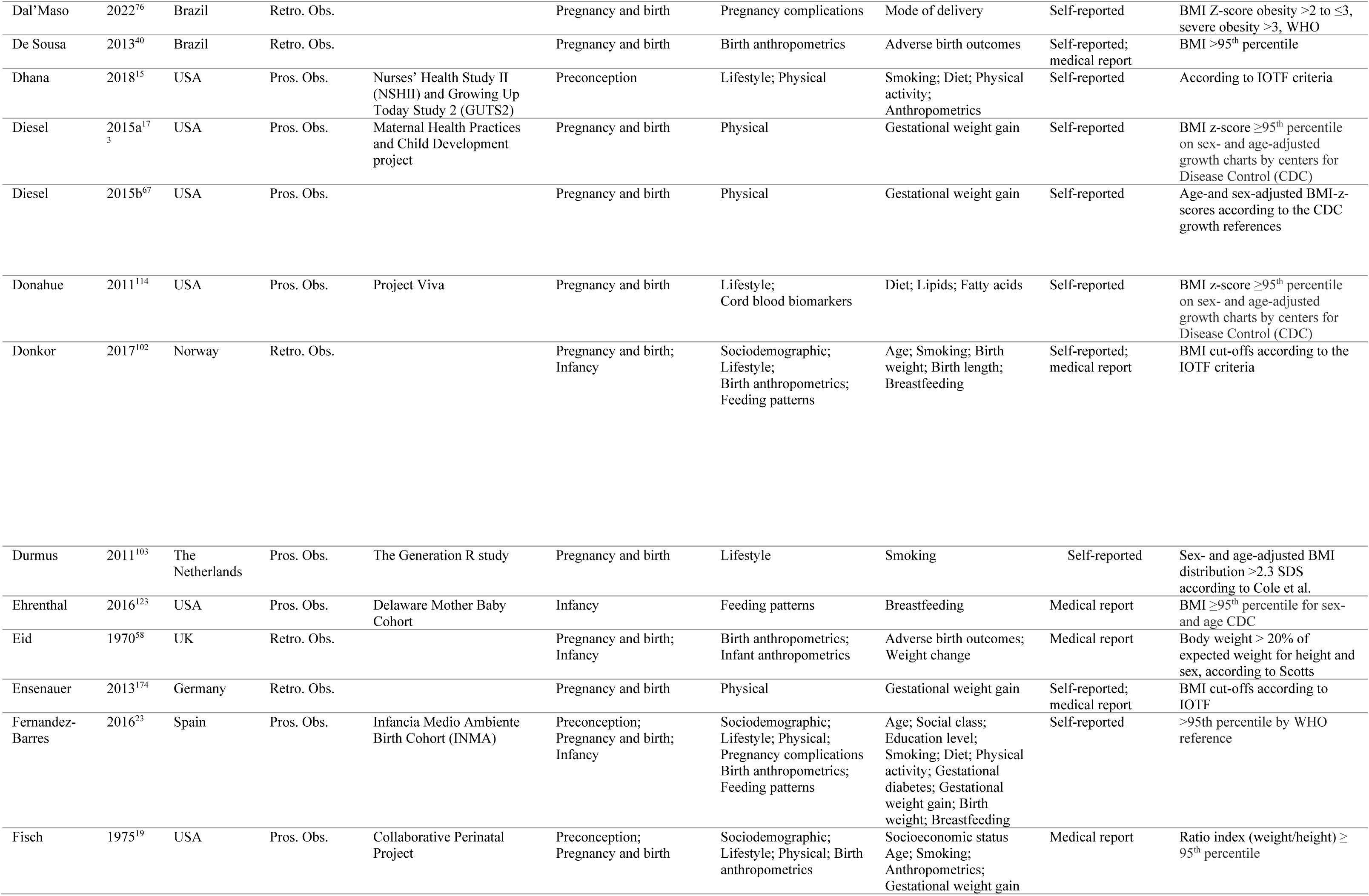
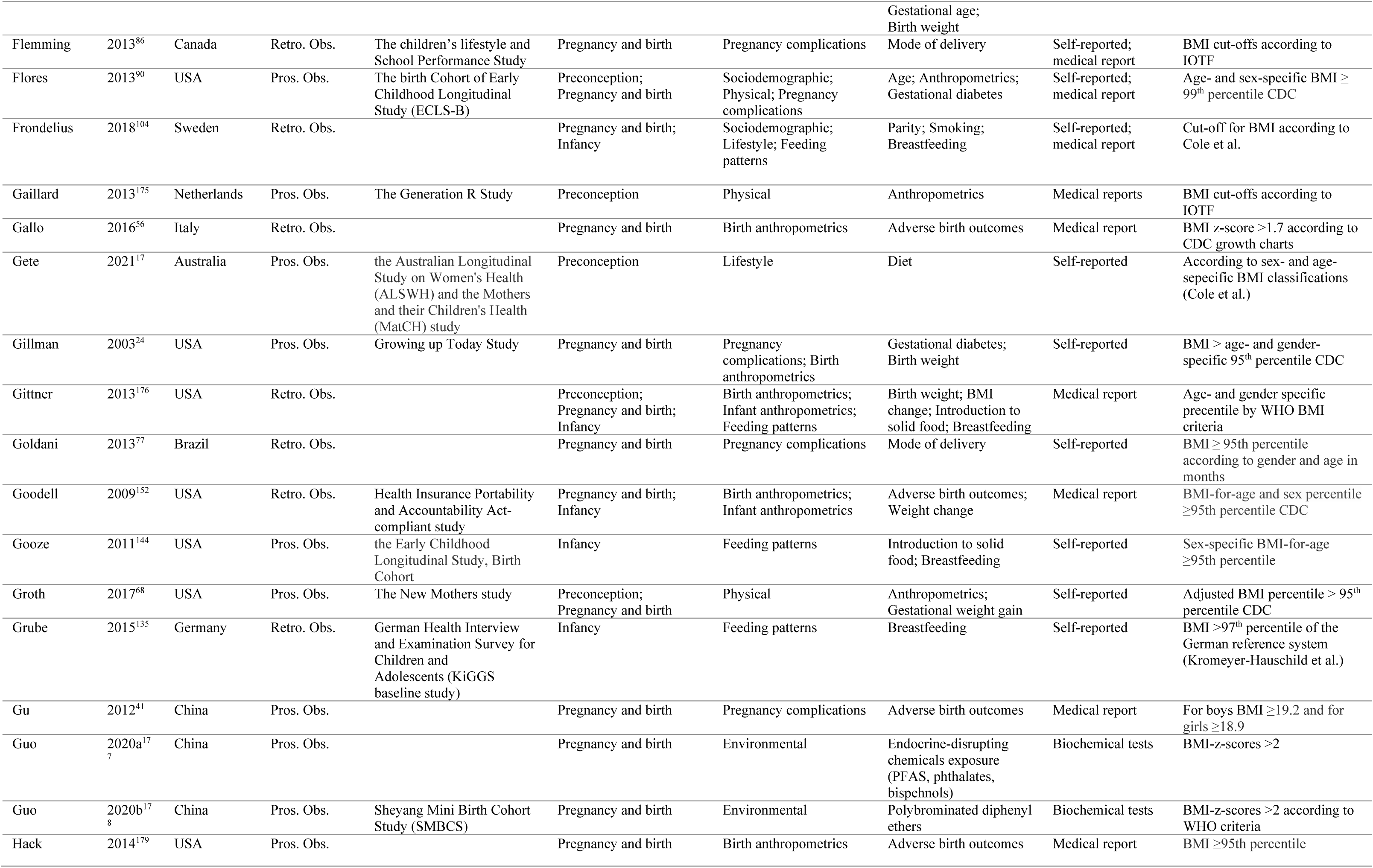
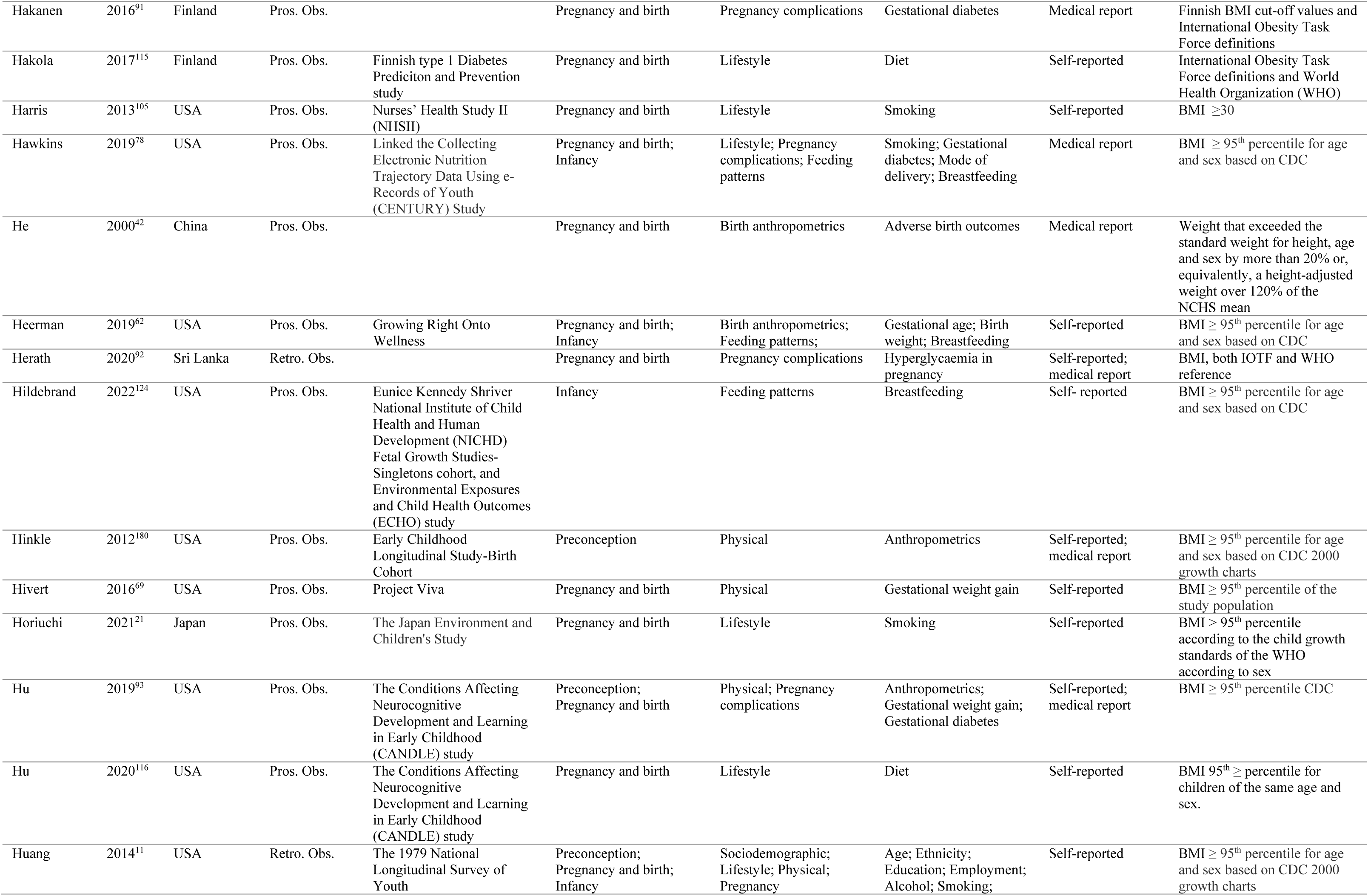
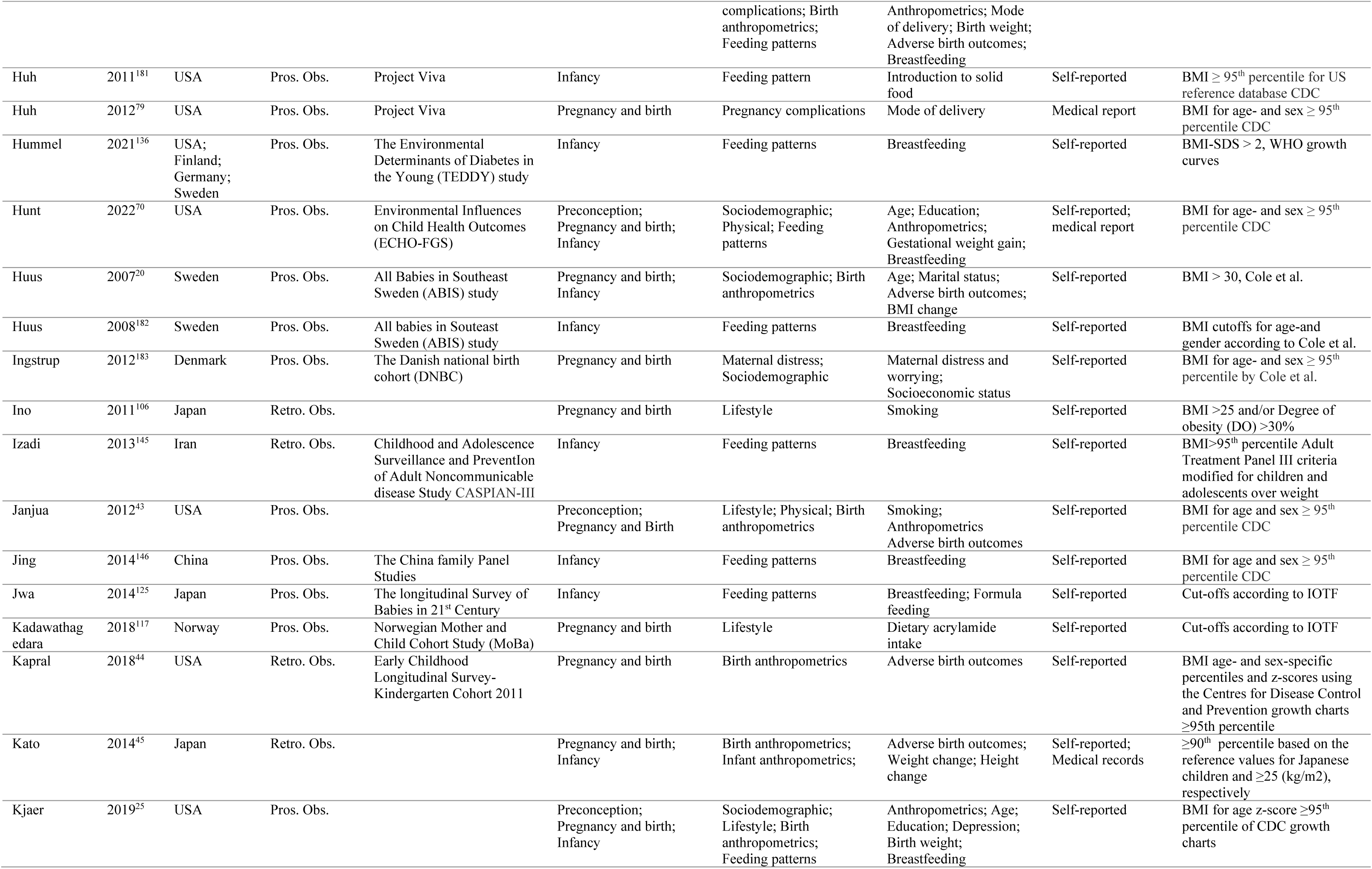
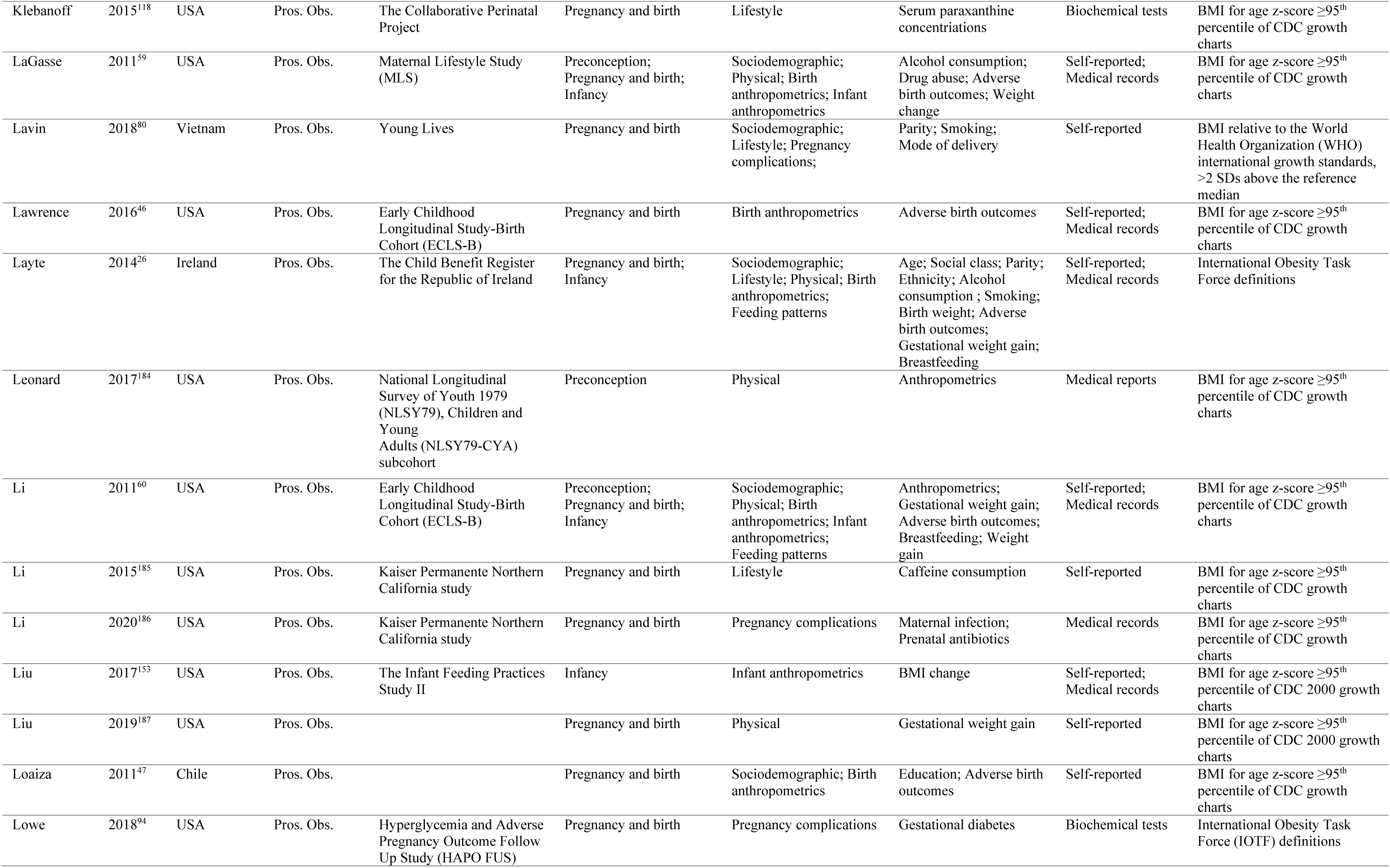
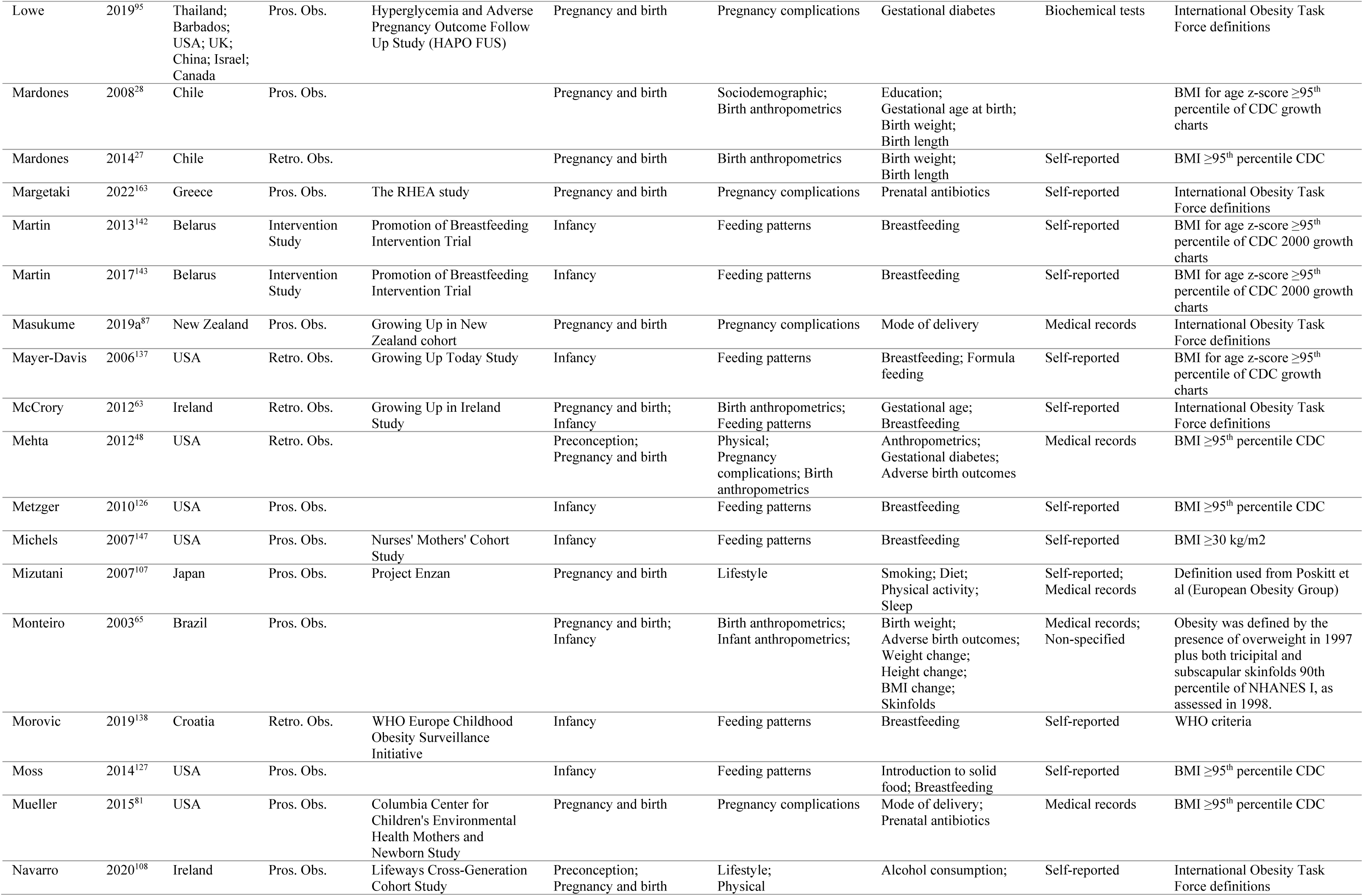
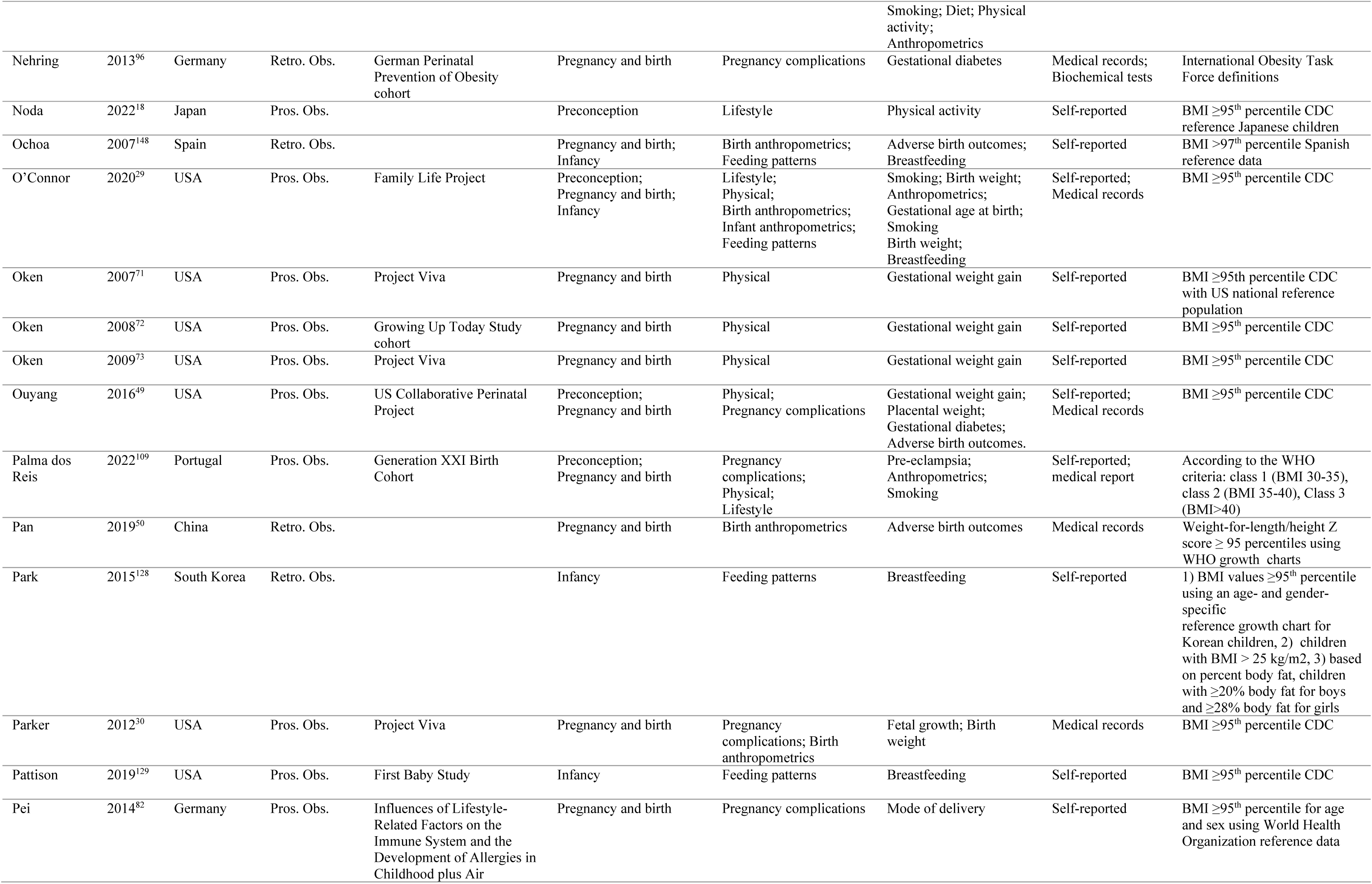
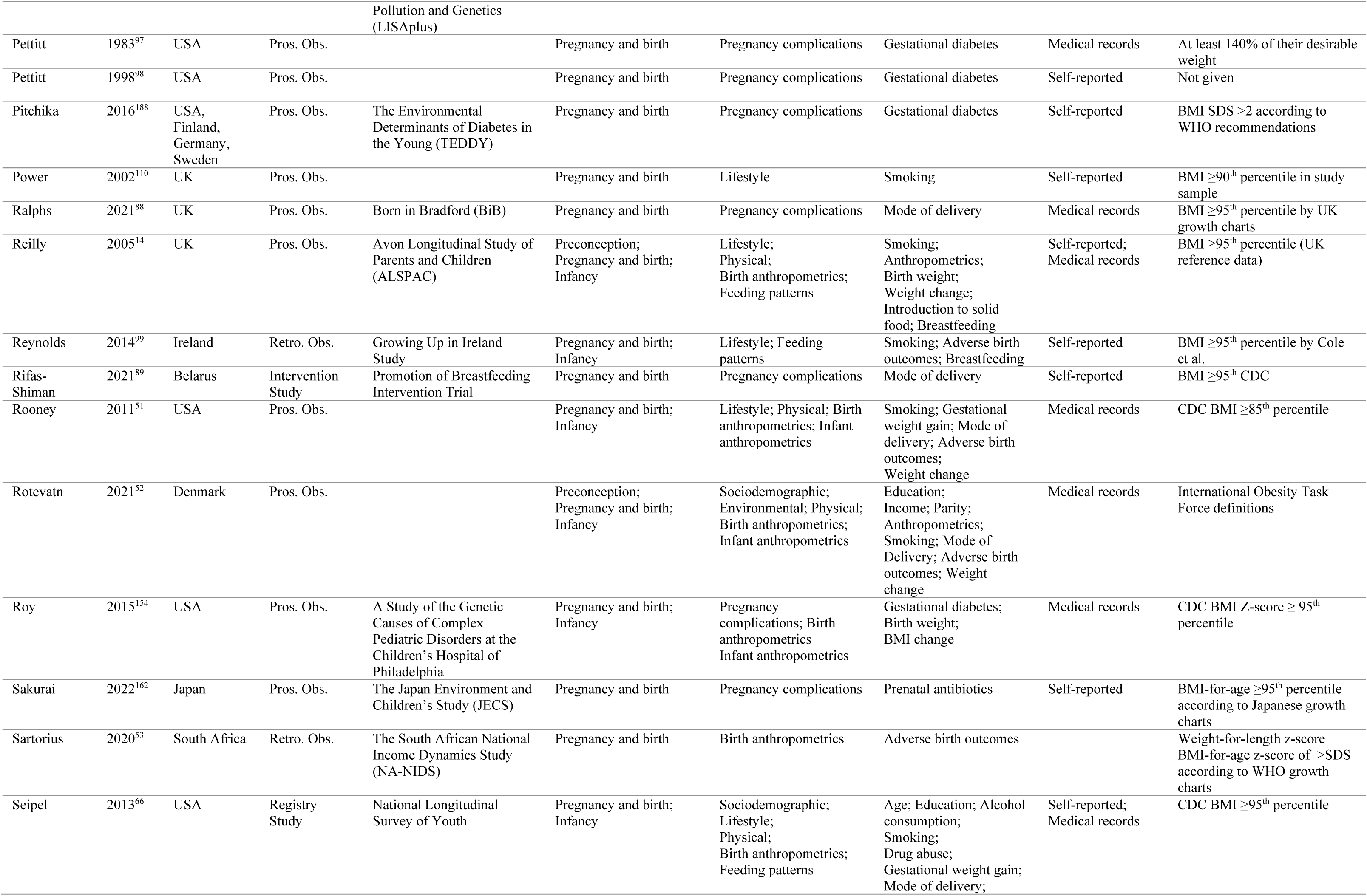
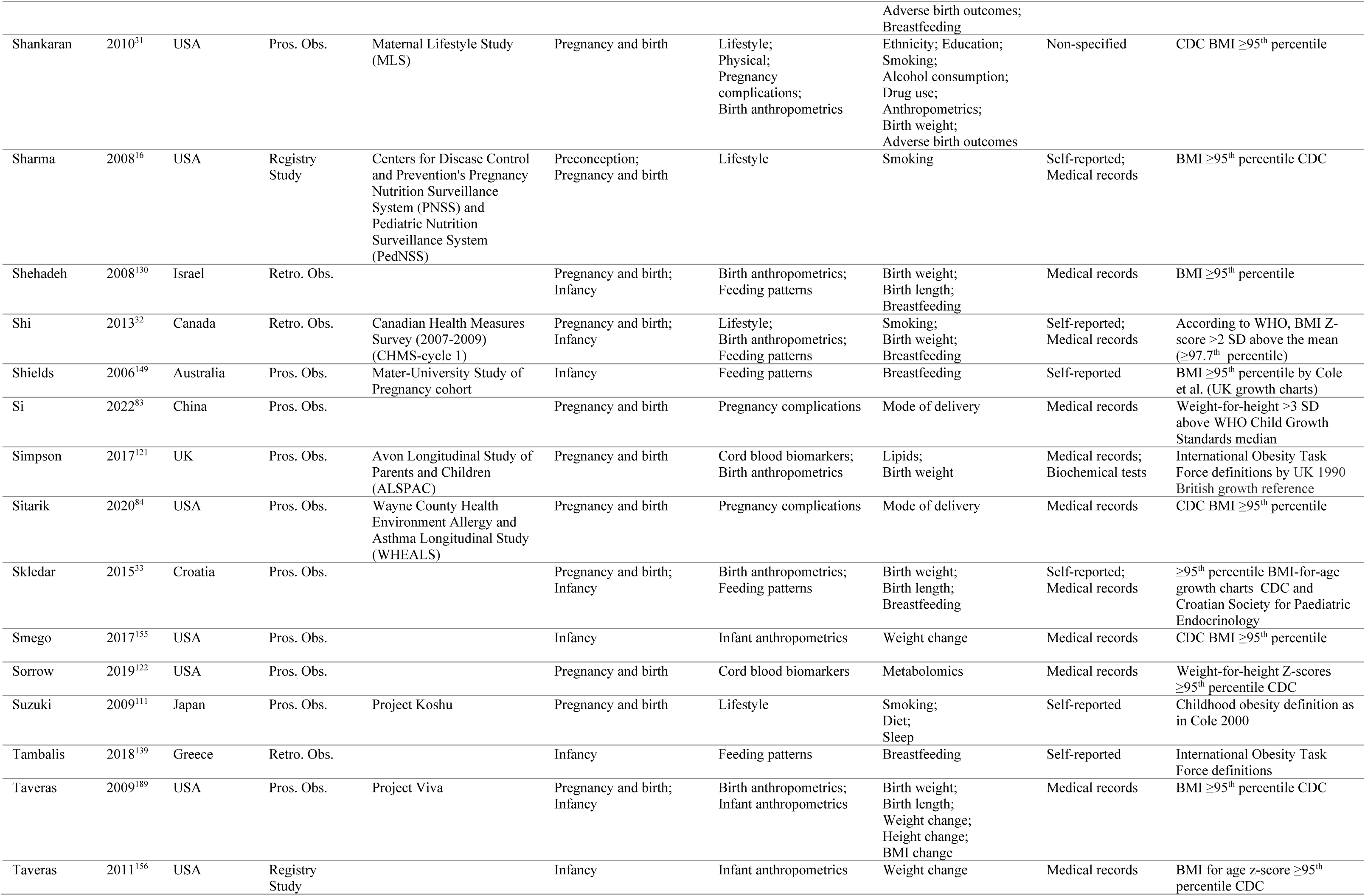
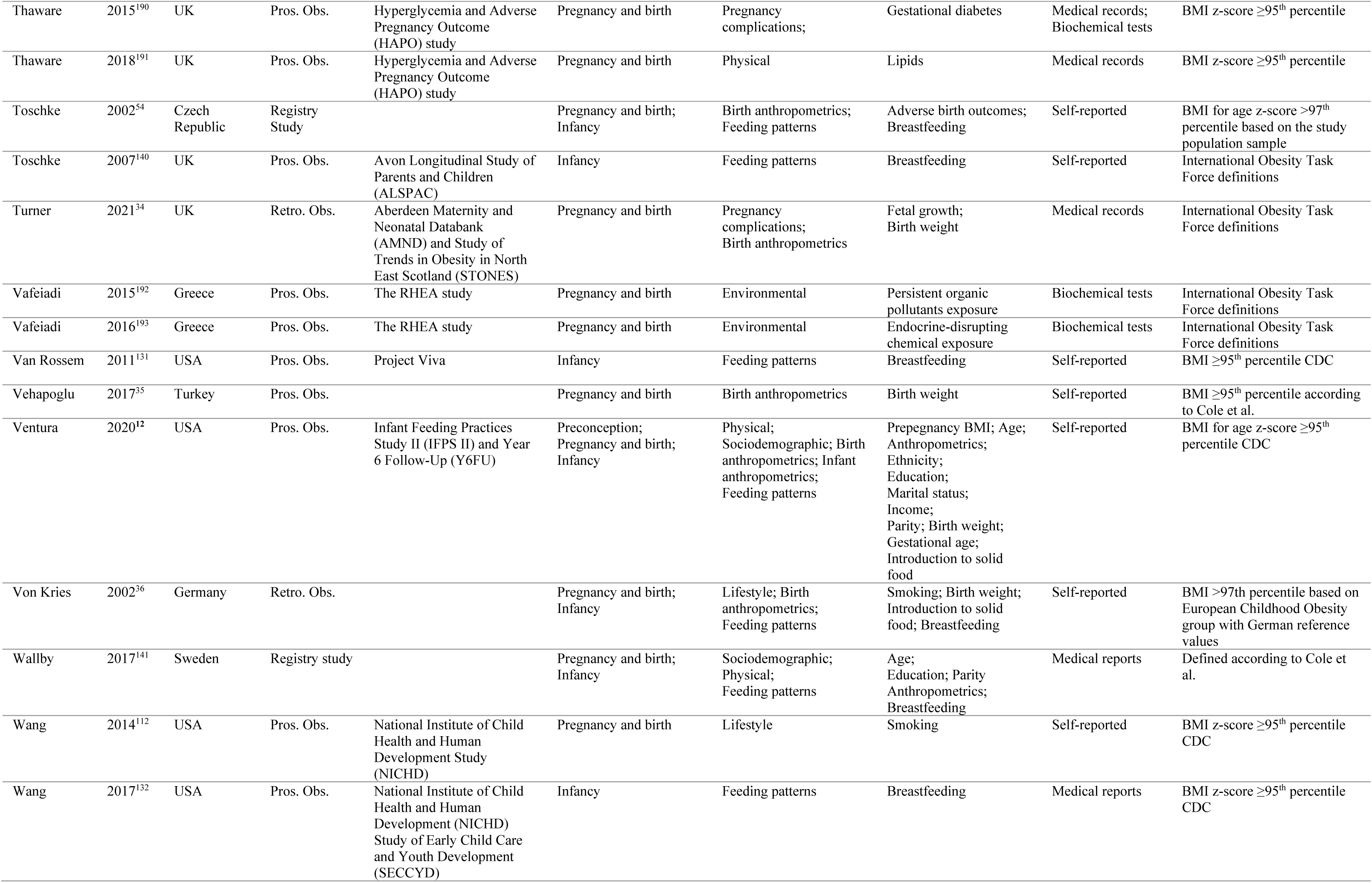
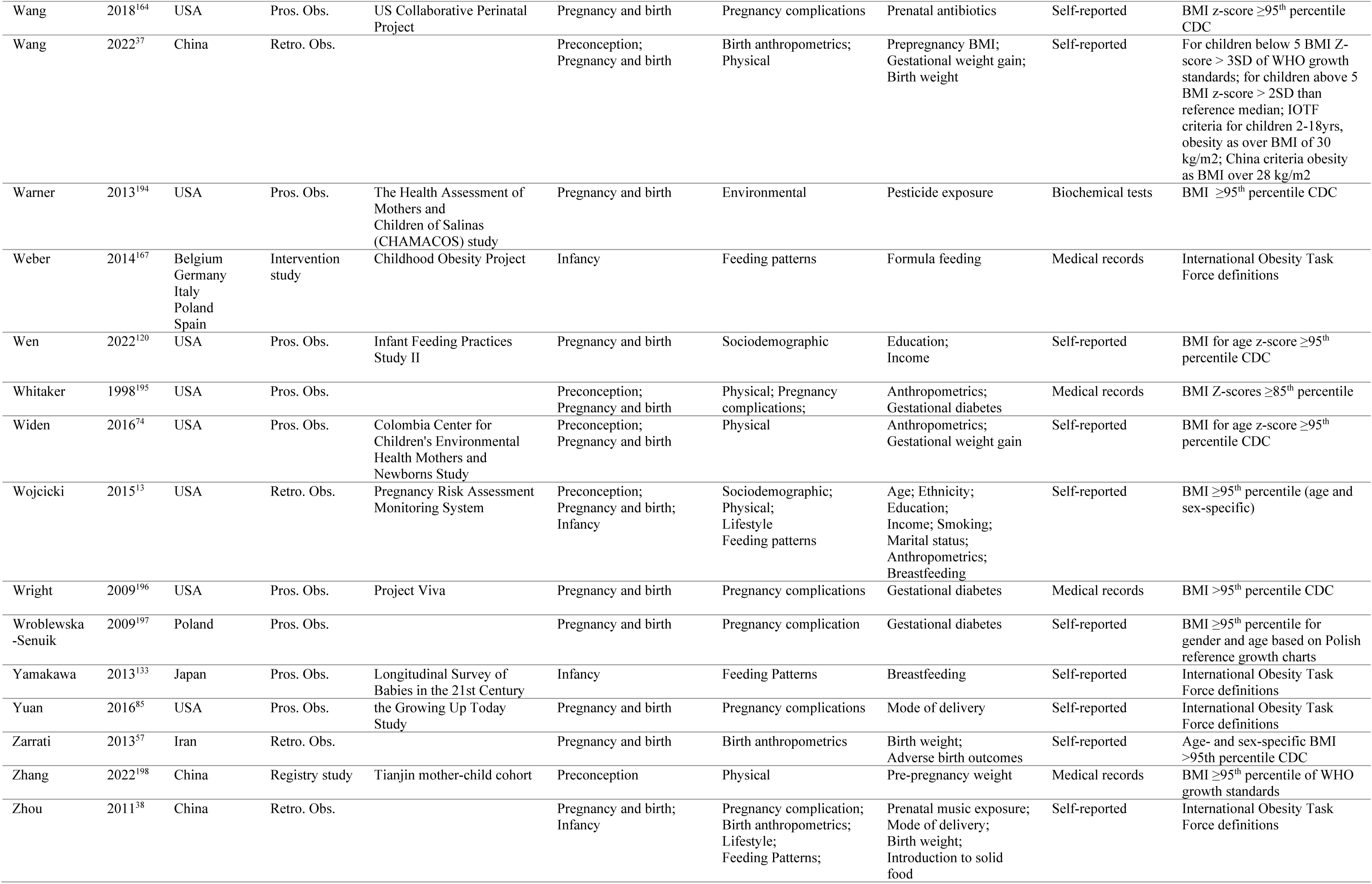
Characteristics of studies included in the systematic review.

| Author | Year | Country | Study type | Study name or site<br>(Year of birth) | Exposure (time period) | Exposure (main) | Exposure (sub) | Exposure<br>ascertainment | Definition of Obesity |
| --- | --- | --- | --- | --- | --- | --- | --- | --- | --- |
| Alves | 2016<sup>64</sup> | Brazil | Retro. Obs. |  | Pregnancy and Birth | Birth anthropometrics | Adverse birth outcomes | Medical report | >97<sup>th</sup> percentile WHO |
| Aris | 2017<sup>150</sup> | Singapore | Pros. Obs. | Growing up in Singapore<br>Towards Healthy Outcomes<br>Study | Infancy | Infant anthropometrics | BMI change | Medical report | Age- and sex specific BMI-z-<br>scores 2 SD > median of<br>World Health Organization<br>Growth Charts |
| Aris | 2018<sup>151</sup> | USA;<br>Belarus | Pros. Obs. | Project Viva and Promotion<br>of Breastfeeding<br>Intervention Trial | Infancy | Infant anthropometrics | Weight change | Medical report | BMI ≥ 95<sup>th</sup> percentile CDC |
| Brophy | 2009<sup>22</sup> | UK | Pros. Obs. | Millenium Cohort Study | Preconception;<br>Pregnancy and Birth;<br>Infancy | Birth anthropometrics;<br>Feeding patterns;<br>Physical | Birth weight;<br>Anthropometrics;<br>Introduction to solid<br>food | Self-reported | International Obesity Task<br>Force definitions |
| Bryl | 2022<sup>172</sup> | Poland | Retro. Obs. |  | Pregnancy and birth | Lifestyle | Maternal stress | Self-reported | International Obesity Task<br>Force definitions |
| Burdette | 2007<sup>134</sup> | USA | Pros. Obs. | The fragile families and<br>child well-being study | Infancy | Feeding patterns | Breastfeeding | Self-reported | BMI > 95<sup>th</sup> percentile CDC |
| Callanan | 2021<sup>119</sup> | Ireland | Intervention<br>Study | A randomized control trial<br>of a low glycaemic index<br>diet in pregnancy to prevent<br>macrosomia (ROLO) | Pregnancy and Birth | Lifestyle | Diet | Medical report | UK-growth chart cut-off >98<sup>th</sup><br>percentile or WHO- cut off ≥<br>95<sup>th</sup> percentile |
| Carrillo-Larco | 2015<sup>75</sup> | Peru | Pros. Obs. | Young Lives Study | Pregnancy and Birth | Pregnancy<br>Complications | Mode of Delivery | Self-reported | International Obesity Task<br>Force definitions |
| Chen | 2006<sup>100</sup> | USA | Pros. Obs. | Collaborative Perinatal<br>Project (CPP) | Pregnancy and Birth | Lifestyle | Smoking | Self-reported | Age- and sex-specific BMI<br>>95<sup>th</sup> percentile CDC |
| Chen | 2017<sup>61</sup> | Taiwan | Pros. Obs. | Taiwan Birth Cohort Study<br>(TBCS) | Pregnancy and Birth | Sociodemographic;<br>Pregnancy<br>complications; Birth<br>anthropometrics; | Maternal age;<br>Anthropometrics;<br>Gestational weight gain;<br>Gestational diabetes<br>Mode of Delivery;<br>Gestational age; | Self-reported;<br>medical report | BMI by Taiwan Bureau of<br>Health Promotion for preschool<br>children |
| Chen | 2019a<sup>113</sup> | Ireland | Pros. Obs. | Lifeways Cross generation<br>Cohort | Pregnancy and birth | Lifestyle | Diet | Self-reported | BMIz-score >95<sup>th</sup> percentile,<br>UK growth reference Cole |
| Chen | 2019b<sup>55</sup> | China | Retro. Obs. |  | Pregnancy and birth | Birth anthropometrics | Adverse birth outcomes | Self-reported | International Obesity Task<br>Force definitions |
| Chiasson | 2016<sup>39</sup> | USA | Pros. Obs. | New York State Special<br>Supplemental Nutrition<br>Program for Women,<br>infants, and children | Pregnancy and birth;<br>Infancy | Birth anthropometrics;<br>Feeding patterns | Adverse birth outcomes;<br>Breastfeeding | Self-reported;<br>medical report | Age- and sex- specific BMI ≥<br>95<sup>th</sup> percentile CDC |
| Chivers | 2012<sup>101</sup> | Australia | Pros. Obs. | The Western Australian<br>Pregnancy Cohort (RAINE) | Preconception;<br>Pregnancy and birth | Sociodemographic;<br>Lifestyle; Physical | Education; smoking;<br>Anthropometrics | Self-reported | International Obesity Task<br>Force definitions |
| Dal'Maso | 2022<sup>76</sup> | Brazil | Retro. Obs. |  | Pregnancy and birth | Pregnancy complications | Mode of delivery | Self-reported | BMI Z-score obesity >2 to ≤3, severe obesity >3, WHO |
| De Sousa | 2013<sup>40</sup> | Brazil | Retro. Obs. |  | Pregnancy and birth | Birth anthropometrics | Adverse birth outcomes | Self-reported; medical report | BMI >95<sup>th</sup> percentile |
| Dhana | 2018<sup>15</sup> | USA | Pros. Obs. | Nurses' Health Study II (NSHII) and Growing Up Today Study 2 (GUTS2) | Preconception | Lifestyle; Physical | Smoking; Diet; Physical activity; Anthropometrics | Self-reported | According to IOTF criteria |
| Diesel | 2015a<sup>173</sup> | USA | Pros. Obs. | Maternal Health Practices and Child Development project | Pregnancy and birth | Physical | Gestational weight gain | Self-reported | BMI z-score ≥95<sup>th</sup> percentile on sex- and age-adjusted growth charts by centers for Disease Control (CDC) |
| Diesel | 2015b<sup>67</sup> | USA | Pros. Obs. |  | Pregnancy and birth | Physical | Gestational weight gain | Self-reported | Age- and sex-adjusted BMI-z-scores according to the CDC growth references |
| Donahue | 2011<sup>114</sup> | USA | Pros. Obs. | Project Viva | Pregnancy and birth | Lifestyle; Cord blood biomarkers | Diet; Lipids; Fatty acids | Self-reported | BMI z-score ≥95<sup>th</sup> percentile on sex- and age-adjusted growth charts by centers for Disease Control (CDC) |
| Donkor | 2017<sup>102</sup> | Norway | Retro. Obs. |  | Pregnancy and birth; Infancy | Sociodemographic; Lifestyle; Birth anthropometrics; Feeding patterns | Age; Smoking; Birth weight; Birth length; Breastfeeding | Self-reported; medical report | BMI cut-offs according to the IOTF criteria |
| Durmus | 2011<sup>103</sup> | The Netherlands | Pros. Obs. | The Generation R study | Pregnancy and birth | Lifestyle | Smoking | Self-reported | Sex- and age-adjusted BMI distribution >2.3 SDS according to Cole et al. |
| Ehrenthal | 2016<sup>123</sup> | USA | Pros. Obs. | Delaware Mother Baby Cohort | Infancy | Feeding patterns | Breastfeeding | Medical report | BMI ≥95<sup>th</sup> percentile for sex- and age CDC |
| Eid | 1970<sup>58</sup> | UK | Retro. Obs. |  | Pregnancy and birth; Infancy | Birth anthropometrics; Infant anthropometrics | Adverse birth outcomes; Weight change | Medical report | Body weight > 20% of expected weight for height and sex, according to Scotts |
| Ensenauer | 2013<sup>174</sup> | Germany | Retro. Obs. |  | Pregnancy and birth | Physical | Gestational weight gain | Self-reported; medical report | BMI cut-offs according to IOTF |
| Fernandez-Barres | 2016<sup>23</sup> | Spain | Pros. Obs. | Infancia Medio Ambiente Birth Cohort (INMA) | Preconception; Pregnancy and birth; Infancy | Sociodemographic; Lifestyle; Physical; Pregnancy complications Birth anthropometrics; Feeding patterns | Age; Social class; Education level; Smoking; Diet; Physical activity; Gestational diabetes; Gestational weight gain; Birth weight; Breastfeeding | Self-reported | >95<sup>th</sup> percentile by WHO reference |
| Fisch | 1975<sup>19</sup> | USA | Pros. Obs. | Collaborative Perinatal Project | Preconception; Pregnancy and birth | Sociodemographic; Lifestyle; Physical; Birth anthropometrics | Socioeconomic status Age; Smoking; Anthropometrics; Gestational weight gain | Medical report | Ratio index (weight/height) ≥ 95<sup>th</sup> percentile |
|  |  |  |  |  |  |  | Gestational age;<br>Birth weight |  |  |
| Flemming | 2013<sup>86</sup> | Canada | Retro. Obs. | The children's lifestyle and School Performance Study | Pregnancy and birth | Pregnancy complications | Mode of delivery | Self-reported;<br>medical report | BMI cut-offs according to IOTF |
| Flores | 2013<sup>90</sup> | USA | Pros. Obs. | The birth Cohort of Early Childhood Longitudinal Study (ECLS-B) | Preconception;<br>Pregnancy and birth | Sociodemographic;<br>Physical; Pregnancy complications | Age; Anthropometrics;<br>Gestational diabetes | Self-reported;<br>medical report | Age- and sex-specific BMI ≥ 99<sup>th</sup> percentile CDC |
| Frondelius | 2018<sup>104</sup> | Sweden | Retro. Obs. |  | Pregnancy and birth;<br>Infancy | Sociodemographic;<br>Lifestyle; Feeding patterns | Parity; Smoking;<br>Breastfeeding | Self-reported;<br>medical report | Cut-off for BMI according to Cole et al. |
| Gaillard | 2013<sup>175</sup> | Netherlands | Pros. Obs. | The Generation R Study | Preconception | Physical | Anthropometrics | Medical reports | BMI cut-offs according to IOTF |
| Gallo | 2016<sup>56</sup> | Italy | Retro. Obs. |  | Pregnancy and birth | Birth anthropometrics | Adverse birth outcomes | Medical report | BMI z-score >1.7 according to CDC growth charts |
| Gete | 2021<sup>17</sup> | Australia | Pros. Obs. | the Australian Longitudinal Study on Women's Health (ALSWH) and the Mothers and their Children's Health (MatCH) study | Preconception | Lifestyle | Diet | Self-reported | According to sex- and age-specific BMI classifications (Cole et al.) |
| Gillman | 2003<sup>24</sup> | USA | Pros. Obs. | Growing up Today Study | Pregnancy and birth | Pregnancy complications; Birth anthropometrics | Gestational diabetes;<br>Birth weight | Self-reported | BMI > age- and gender-specific 95<sup>th</sup> percentile CDC |
| Gittner | 2013<sup>176</sup> | USA | Retro. Obs. |  | Preconception;<br>Pregnancy and birth;<br>Infancy | Birth anthropometrics;<br>Infant anthropometrics;<br>Feeding patterns | Birth weight; BMI change; Introduction to solid food; Breastfeeding | Medical report | Age- and gender specific percentile by WHO BMI criteria |
| Goldani | 2013<sup>77</sup> | Brazil | Retro. Obs. | | Pregnancy and birth | Pregnancy complications | Mode of delivery | Self-reported | BMI ≥ 95th percentile according to gender and age in months |
| Goodell | 2009<sup>152</sup> | USA | Retro. Obs. | Health Insurance Portability and Accountability Act-compliant study | Pregnancy and birth;<br>Infancy | Birth anthropometrics;<br>Infant anthropometrics | Adverse birth outcomes;<br>Weight change | Medical report | BMI-for-age and sex percentile ≥95th percentile CDC |
| Gooze | 2011<sup>144</sup> | USA | Pros. Obs. | the Early Childhood Longitudinal Study, Birth Cohort | Infancy | Feeding patterns | Introduction to solid food; Breastfeeding | Self-reported | Sex-specific BMI-for-age ≥95th percentile |
| Groth | 2017<sup>68</sup> | USA | Pros. Obs. | The New Mothers study | Preconception;<br>Pregnancy and birth | Physical | Anthropometrics;<br>Gestational weight gain | Self-reported | Adjusted BMI percentile > 95<sup>th</sup> percentile CDC |
| Grube | 2015<sup>135</sup> | Germany | Retro. Obs. | German Health Interview and Examination Survey for Children and Adolescents (KiGGS baseline study) | Infancy | Feeding patterns | Breastfeeding | Self-reported | BMI >97<sup>th</sup> percentile of the German reference system (Kromeyer-Hauschild et al.) |
| Gu | 2012<sup>41</sup> | China | Pros. Obs. | | Pregnancy and birth | Pregnancy complications | Adverse birth outcomes | Medical report | For boys BMI ≥19.2 and for girls ≥18.9 |
| Guo | 2020a<sup>177</sup> | China | Pros. Obs. |  | Pregnancy and birth | Environmental | Endocrine-disrupting chemicals exposure (PFAS, phthalates, bisphenols) | Biochemical tests | BMI-z-scores >2 |
| Guo | 2020b<sup>178</sup> | China | Pros. Obs. | Sheyang Mini Birth Cohort Study (SMBCS) | Pregnancy and birth | Environmental | Polybrominated diphenyl ethers | Biochemical tests | BMI-z-scores >2 according to WHO criteria |
| Hack | 2014<sup>179</sup> | USA | Pros. Obs. | | Pregnancy and birth | Birth anthropometrics | Adverse birth outcomes | Medical report | BMI ≥95th percentile |
| Hakanen | 2016<sup>91</sup> | Finland | Pros. Obs. |  | Pregnancy and birth | Pregnancy complications | Gestational diabetes | Medical report | Finnish BMI cut-off values and International Obesity Task Force definitions |
| Hakola | 2017<sup>115</sup> | Finland | Pros. Obs. | Finnish type 1 Diabetes Predicition and Prevention study | Pregnancy and birth | Lifestyle | Diet | Self-reported | International Obesity Task Force definitions and World Health Organization (WHO) |
| Harris | 2013<sup>105</sup> | USA | Pros. Obs. | Nurses' Health Study II (NHSII) | Pregnancy and birth | Lifestyle | Smoking | Self-reported | BMI $\geq 30$ |
| Hawkins | 2019<sup>78</sup> | USA | Pros. Obs. | Linked the Collecting Electronic Nutrition Trajectory Data Using e-Records of Youth (CENTURY) Study | Pregnancy and birth; Infancy | Lifestyle; Pregnancy complications; Feeding patterns | Smoking; Gestational diabetes; Mode of delivery; Breastfeeding | Medical report | BMI $\geq 95^{\text{th}}$ percentile for age and sex based on CDC |
| He | 2000<sup>42</sup> | China | Pros. Obs. |  | Pregnancy and birth | Birth anthropometrics | Adverse birth outcomes | Medical report | Weight that exceeded the standard weight for height, age and sex by more than 20% or, equivalently, a height-adjusted weight over 120% of the NCHS mean |
| Heerman | 2019<sup>62</sup> | USA | Pros. Obs. | Growing Right Onto Wellness | Pregnancy and birth; Infancy | Birth anthropometrics; Feeding patterns; | Gestational age; Birth weight; Breastfeeding | Self-reported | BMI $\geq 95^{\text{th}}$ percentile for age and sex based on CDC |
| Herath | 2020<sup>92</sup> | Sri Lanka | Retro. Obs. |  | Pregnancy and birth | Pregnancy complications | Hyperglycaemia in pregnancy | Self-reported; medical report | BMI, both IOTF and WHO reference |
| Hildebrand | 2022<sup>124</sup> | USA | Pros. Obs. | Eunice Kennedy Shriver National Institute of Child Health and Human Development (NICHD) Fetal Growth Studies-Singletons cohort, and Environmental Exposures and Child Health Outcomes (ECHO) study | Infancy | Feeding patterns | Breastfeeding | Self-reported | BMI $\geq 95^{\text{th}}$ percentile for age and sex based on CDC |
| Hinkle | 2012<sup>180</sup> | USA | Pros. Obs. | Early Childhood Longitudinal Study-Birth Cohort | Preconception | Physical | Anthropometrics | Self-reported; medical report | BMI $\geq 95^{\text{th}}$ percentile for age and sex based on CDC 2000 growth charts |
| Hivert | 2016<sup>69</sup> | USA | Pros. Obs. | Project Viva | Pregnancy and birth | Physical | Gestational weight gain | Self-reported | BMI $\geq 95^{\text{th}}$ percentile of the study population |
| Horiuchi | 2021<sup>21</sup> | Japan | Pros. Obs. | The Japan Environment and Children's Study | Pregnancy and birth | Lifestyle | Smoking | Self-reported | BMI $> 95^{\text{th}}$ percentile according to the child growth standards of the WHO according to sex |
| Hu | 2019<sup>93</sup> | USA | Pros. Obs. | The Conditions Affecting Neurocognitive Development and Learning in Early Childhood (CANDLE) study | Preconception; Pregnancy and birth | Physical; Pregnancy complications | Anthropometrics; Gestational weight gain; Gestational diabetes | Self-reported; medical report | BMI $\geq 95^{\text{th}}$ percentile CDC |
| Hu | 2020<sup>116</sup> | USA | Pros. Obs. | The Conditions Affecting Neurocognitive Development and Learning in Early Childhood (CANDLE) study | Pregnancy and birth | Lifestyle | Diet | Self-reported | BMI $95^{\text{th}} \geq$ percentile for children of the same age and sex. |
| Huang | 2014<sup>11</sup> | USA | Retro. Obs. | The 1979 National Longitudinal Survey of Youth | Preconception; Pregnancy and birth; Infancy | Sociodemographic; Lifestyle; Physical; Pregnancy | Age; Ethnicity; Education; Employment; Alcohol; Smoking; | Self-reported | BMI $\geq 95^{\text{th}}$ percentile for age and sex based on CDC 2000 growth charts |
|  |  |  |  |  |  | complications; Birth anthropometrics; Feeding patterns | Anthropometrics; Mode of delivery; Birth weight; Adverse birth outcomes; Breastfeeding |  |  |
| Huh | 2011<sup>181</sup> | USA | Pros. Obs. | Project Viva | Infancy | Feeding pattern | Introduction to solid food | Self-reported | BMI ≥ 95<sup>th</sup> percentile for US reference database CDC |
| Huh | 2012<sup>79</sup> | USA | Pros. Obs. | Project Viva | Pregnancy and birth | Pregnancy complications | Mode of delivery | Medical report | BMI for age- and sex ≥ 95<sup>th</sup> percentile CDC |
| Hummel | 2021<sup>136</sup> | USA; Finland; Germany; Sweden | Pros. Obs. | The Environmental Determinants of Diabetes in the Young (TEDDY) study | Infancy | Feeding patterns | Breastfeeding | Self-reported | BMI-SDS > 2, WHO growth curves |
| Hunt | 2022<sup>70</sup> | USA | Pros. Obs. | Environmental Influences on Child Health Outcomes (ECHO-FGS) | Preconception; Pregnancy and birth; Infancy | Sociodemographic; Physical; Feeding patterns | Age; Education; Anthropometrics; Gestational weight gain; Breastfeeding | Self-reported; medical report | BMI for age- and sex ≥ 95<sup>th</sup> percentile CDC |
| Huus | 2007<sup>20</sup> | Sweden | Pros. Obs. | All Babies in Southeast Sweden (ABIS) study | Pregnancy and birth; Infancy | Sociodemographic; Birth anthropometrics | Age; Marital status; Adverse birth outcomes; BMI change | Self-reported | BMI > 30, Cole et al. |
| Huus | 2008<sup>182</sup> | Sweden | Pros. Obs. | All babies in Souteast Sweden (ABIS) study | Infancy | Feeding patterns | Breastfeeding | Self-reported | BMI cutoffs for age-and gender according to Cole et al. |
| Ingstrup | 2012<sup>183</sup> | Denmark | Pros. Obs. | The Danish national birth cohort (DNBC) | Pregnancy and birth | Maternal distress; Sociodemographic | Maternal distress and worrying; Socioeconomic status | Self-reported | BMI for age- and sex ≥ 95<sup>th</sup> percentile by Cole et al. |
| Ino | 2011<sup>106</sup> | Japan | Retro. Obs. |  | Pregnancy and birth | Lifestyle | Smoking | Self-reported | BMI >25 and/or Degree of obesity (DO) >30% |
| Izadi | 2013<sup>145</sup> | Iran | Retro. Obs. | Childhood and Adolescence Surveillance and Prevention of Adult Noncommunicable disease Study CASPIAN-III | Infancy | Feeding patterns | Breastfeeding | Self-reported | BMI>95<sup>th</sup> percentile Adult Treatment Panel III criteria modified for children and adolescents over weight |
| Janjua | 2012<sup>43</sup> | USA | Pros. Obs. | | Preconception; Pregnancy and Birth | Lifestyle; Physical; Birth anthropometrics | Smoking; Anthropometrics Adverse birth outcomes | Self-reported | BMI for age and sex ≥ 95<sup>th</sup> percentile CDC |
| Jing | 2014<sup>146</sup> | China | Pros. Obs. | The China family Panel Studies | Infancy | Feeding patterns | Breastfeeding | Self-reported | BMI for age and sex ≥ 95<sup>th</sup> percentile CDC |
| Jwa | 2014<sup>125</sup> | Japan | Pros. Obs. | The longitudinal Survey of Babies in 21<sup>st</sup> Century | Infancy | Feeding patterns | Breastfeeding; Formula feeding | Self-reported | Cut-offs according to IOTF |
| Kadawathag edara | 2018<sup>117</sup> | Norway | Pros. Obs. | Norwegian Mother and Child Cohort Study (MoBa) | Pregnancy and birth | Lifestyle | Dietary acrylamide intake | Self-reported | Cut-offs according to IOTF |
| Kapral | 2018<sup>44</sup> | USA | Retro. Obs. | Early Childhood Longitudinal Survey- Kindergarten Cohort 2011 | Pregnancy and birth | Birth anthropometrics | Adverse birth outcomes | Self-reported | BMI age- and sex-specific percentiles and z-scores using the Centres for Disease Control and Prevention growth charts ≥95<sup>th</sup> percentile |
| Kato | 2014<sup>45</sup> | Japan | Retro. Obs. | | Pregnancy and birth; Infancy | Birth anthropometrics; Infant anthropometrics; | Adverse birth outcomes; Weight change; Height change | Self-reported; Medical records | ≥90<sup>th</sup> percentile based on the reference values for Japanese children and ≥25 (kg/m<sup>2</sup>), respectively |
| Kjaer | 2019<sup>25</sup> | USA | Pros. Obs. | | Preconception; Pregnancy and birth; Infancy | Sociodemographic; Lifestyle; Birth anthropometrics; Feeding patterns | Anthropometrics; Age; Education; Depression; Birth weight; Breastfeeding | Self-reported | BMI for age z-score ≥95<sup>th</sup> percentile of CDC growth charts |
| Klebanoff | 2015<sup>118</sup> | USA | Pros. Obs. | The Collaborative Perinatal Project | Pregnancy and birth | Lifestyle | Serum paraxanthine concentrations | Biochemical tests | BMI for age z-score $\geq 95^{\text{th}}$ percentile of CDC growth charts |
| LaGasse | 2011<sup>59</sup> | USA | Pros. Obs. | Maternal Lifestyle Study (MLS) | Preconception; Pregnancy and birth; Infancy | Sociodemographic; Physical; Birth anthropometrics; Infant anthropometrics | Alcohol consumption; Drug abuse; Adverse birth outcomes; Weight change | Self-reported; Medical records | BMI for age z-score $\geq 95^{\text{th}}$ percentile of CDC growth charts |
| Lavin | 2018<sup>80</sup> | Vietnam | Pros. Obs. | Young Lives | Pregnancy and birth | Sociodemographic; Lifestyle; Pregnancy complications; | Parity; Smoking; Mode of delivery | Self-reported | BMI relative to the World Health Organization (WHO) international growth standards, >2 SDs above the reference median |
| Lawrence | 2016<sup>46</sup> | USA | Pros. Obs. | Early Childhood Longitudinal Study-Birth Cohort (ECLS-B) | Pregnancy and birth | Birth anthropometrics | Adverse birth outcomes | Self-reported; Medical records | BMI for age z-score $\geq 95^{\text{th}}$ percentile of CDC growth charts |
| Layte | 2014<sup>26</sup> | Ireland | Pros. Obs. | The Child Benefit Register for the Republic of Ireland | Pregnancy and birth; Infancy | Sociodemographic; Lifestyle; Physical; Birth anthropometrics; Feeding patterns | Age; Social class; Parity; Ethnicity; Alcohol consumption ; Smoking; Birth weight; Adverse birth outcomes; Gestational weight gain; Breastfeeding | Self-reported; Medical records | International Obesity Task Force definitions |
| Leonard | 2017<sup>184</sup> | USA | Pros. Obs. | National Longitudinal Survey of Youth 1979 (NLSY79), Children and Young Adults (NLSY79-CYA) subcohort | Preconception | Physical | Anthropometrics | Medical reports | BMI for age z-score $\geq 95^{\text{th}}$ percentile of CDC growth charts |
| Li | 2011<sup>60</sup> | USA | Pros. Obs. | Early Childhood Longitudinal Study-Birth Cohort (ECLS-B) | Preconception; Pregnancy and birth; Infancy | Sociodemographic; Physical; Birth anthropometrics; Infant anthropometrics; Feeding patterns | Anthropometrics; Gestational weight gain; Adverse birth outcomes; Breastfeeding; Weight gain | Self-reported; Medical records | BMI for age z-score $\geq 95^{\text{th}}$ percentile of CDC growth charts |
| Li | 2015<sup>185</sup> | USA | Pros. Obs. | Kaiser Permanente Northern California study | Pregnancy and birth | Lifestyle | Caffeine consumption | Self-reported | BMI for age z-score $\geq 95^{\text{th}}$ percentile of CDC growth charts |
| Li | 2020<sup>186</sup> | USA | Pros. Obs. | Kaiser Permanente Northern California study | Pregnancy and birth | Pregnancy complications | Maternal infection; Prenatal antibiotics | Medical records | BMI for age z-score $\geq 95^{\text{th}}$ percentile of CDC growth charts |
| Liu | 2017<sup>153</sup> | USA | Pros. Obs. | The Infant Feeding Practices Study II | Infancy | Infant anthropometrics | BMI change | Self-reported; Medical records | BMI for age z-score $\geq 95^{\text{th}}$ percentile of CDC 2000 growth charts |
| Liu | 2019<sup>187</sup> | USA | Pros. Obs. | | Pregnancy and birth | Physical | Gestational weight gain | Self-reported | BMI for age z-score $\geq 95^{\text{th}}$ percentile of CDC 2000 growth charts |
| Loaiza | 2011<sup>47</sup> | Chile | Pros. Obs. | | Pregnancy and birth | Sociodemographic; Birth anthropometrics | Education; Adverse birth outcomes | Self-reported | BMI for age z-score $\geq 95^{\text{th}}$ percentile of CDC growth charts |
| Lowe | 2018<sup>94</sup> | USA | Pros. Obs. | Hyperglycemia and Adverse Pregnancy Outcome Follow Up Study (HAPO FUS) | Pregnancy and birth | Pregnancy complications | Gestational diabetes | Biochemical tests | International Obesity Task Force (IOTF) definitions |
| Lowe | 2019<sup>95</sup> | Thailand; Barbados; USA; UK; China; Israel; Canada | Pros. Obs. | Hyperglycemia and Adverse Pregnancy Outcome Follow Up Study (HAPO FUS) | Pregnancy and birth | Pregnancy complications | Gestational diabetes | Biochemical tests | International Obesity Task Force definitions |
| Mardones | 2008<sup>28</sup> | Chile | Pros. Obs. | | Pregnancy and birth | Sociodemographic; Birth anthropometrics | Education; Gestational age at birth; Birth weight; Birth length | | BMI for age z-score $\geq 95^{\text{th}}$ percentile of CDC growth charts |
| Mardones | 2014<sup>27</sup> | Chile | Retro. Obs. | | Pregnancy and birth | Birth anthropometrics | Birth weight; Birth length | Self-reported | BMI $\geq 95^{\text{th}}$ percentile CDC |
| Margetaki | 2022<sup>163</sup> | Greece | Pros. Obs. | The RHEA study | Pregnancy and birth | Pregnancy complications | Prenatal antibiotics | Self-reported | International Obesity Task Force definitions |
| Martin | 2013<sup>142</sup> | Belarus | Intervention Study | Promotion of Breastfeeding Intervention Trial | Infancy | Feeding patterns | Breastfeeding | Self-reported | BMI for age z-score $\geq 95^{\text{th}}$ percentile of CDC 2000 growth charts |
| Martin | 2017<sup>143</sup> | Belarus | Intervention Study | Promotion of Breastfeeding Intervention Trial | Infancy | Feeding patterns | Breastfeeding | Self-reported | BMI for age z-score $\geq 95^{\text{th}}$ percentile of CDC 2000 growth charts |
| Masukume | 2019a<sup>87</sup> | New Zealand | Pros. Obs. | Growing Up in New Zealand cohort | Pregnancy and birth | Pregnancy complications | Mode of delivery | Medical records | International Obesity Task Force definitions |
| Mayer-Davis | 2006<sup>137</sup> | USA | Retro. Obs. | Growing Up Today Study | Infancy | Feeding patterns | Breastfeeding; Formula feeding | Self-reported | BMI for age z-score $\geq 95^{\text{th}}$ percentile of CDC growth charts |
| McCrory | 2012<sup>63</sup> | Ireland | Retro. Obs. | Growing Up in Ireland Study | Pregnancy and birth; Infancy | Birth anthropometrics; Feeding patterns | Gestational age; Breastfeeding | Self-reported | International Obesity Task Force definitions |
| Mehta | 2012<sup>48</sup> | USA | Retro. Obs. | | Preconception; Pregnancy and birth | Physical; Pregnancy complications; Birth anthropometrics | Anthropometrics; Gestational diabetes; Adverse birth outcomes | Medical records | BMI $\geq 95^{\text{th}}$ percentile CDC |
| Metzger | 2010<sup>126</sup> | USA | Pros. Obs. | | Infancy | Feeding patterns | Breastfeeding | Self-reported | BMI $\geq 95^{\text{th}}$ percentile CDC |
| Michels | 2007<sup>147</sup> | USA | Pros. Obs. | Nurses' Mothers' Cohort Study | Infancy | Feeding patterns | Breastfeeding | Self-reported | BMI $\geq 30$ kg/m<sup>2</sup> |
| Mizutani | 2007<sup>107</sup> | Japan | Pros. Obs. | Project Enzan | Pregnancy and birth | Lifestyle | Smoking; Diet; Physical activity; Sleep | Self-reported; Medical records | Definition used from Poskitt et al (European Obesity Group) |
| Monteiro | 2003<sup>65</sup> | Brazil | Pros. Obs. |  | Pregnancy and birth; Infancy | Birth anthropometrics; Infant anthropometrics; | Birth weight; Adverse birth outcomes; Weight change; Height change; BMI change; Skinfolds | Medical records; Non-specified | Obesity was defined by the presence of overweight in 1997 plus both tricipital and subscapular skinfolds 90th percentile of NHANES I, as assessed in 1998. |
| Morovic | 2019<sup>138</sup> | Croatia | Retro. Obs. | WHO Europe Childhood Obesity Surveillance Initiative | Infancy | Feeding patterns | Breastfeeding | Self-reported | WHO criteria |
| Moss | 2014<sup>127</sup> | USA | Pros. Obs. | | Infancy | Feeding patterns | Introduction to solid food; Breastfeeding | Self-reported | BMI $\geq 95^{\text{th}}$ percentile CDC |
| Mueller | 2015<sup>81</sup> | USA | Pros. Obs. | Columbia Center for Children's Environmental Health Mothers and Newborn Study | Pregnancy and birth | Pregnancy complications | Mode of delivery; Prenatal antibiotics | Medical records | BMI $\geq 95^{\text{th}}$ percentile CDC |
| Navarro | 2020<sup>108</sup> | Ireland | Pros. Obs. | Lifeways Cross-Generation Cohort Study | Preconception; Pregnancy and birth | Lifestyle; Physical | Alcohol consumption; | Self-reported | International Obesity Task Force definitions |
|  |  |  |  |  |  |  | Smoking; Diet; Physical activity; Anthropometrics |  |  |
| Nehring | 2013<sup>96</sup> | Germany | Retro. Obs. | German Perinatal Prevention of Obesity cohort | Pregnancy and birth | Pregnancy complications | Gestational diabetes | Medical records; Biochemical tests | International Obesity Task Force definitions |
| Noda | 2022<sup>18</sup> | Japan | Pros. Obs. |  | Preconception | Lifestyle | Physical activity | Self-reported | BMI ≥95<sup>th</sup> percentile CDC reference Japanese children |
| Ochoa | 2007<sup>148</sup> | Spain | Retro. Obs. |  | Pregnancy and birth; Infancy | Birth anthropometrics; Feeding patterns | Adverse birth outcomes; Breastfeeding | Self-reported | BMI >97<sup>th</sup> percentile Spanish reference data |
| O'Connor | 2020<sup>29</sup> | USA | Pros. Obs. | Family Life Project | Preconception; Pregnancy and birth; Infancy | Lifestyle; Physical; Birth anthropometrics; Infant anthropometrics; Feeding patterns | Smoking; Birth weight; Anthropometrics; Gestational age at birth; Smoking Birth weight; Breastfeeding | Self-reported; Medical records | BMI ≥95<sup>th</sup> percentile CDC |
| Oken | 2007<sup>71</sup> | USA | Pros. Obs. | Project Viva | Pregnancy and birth | Physical | Gestational weight gain | Self-reported | BMI ≥95<sup>th</sup> percentile CDC with US national reference population |
| Oken | 2008<sup>72</sup> | USA | Pros. Obs. | Growing Up Today Study cohort | Pregnancy and birth | Physical | Gestational weight gain | Self-reported | BMI ≥95<sup>th</sup> percentile CDC |
| Oken | 2009<sup>73</sup> | USA | Pros. Obs. | Project Viva | Pregnancy and birth | Physical | Gestational weight gain | Self-reported | BMI ≥95<sup>th</sup> percentile CDC |
| Ouyang | 2016<sup>49</sup> | USA | Pros. Obs. | US Collaborative Perinatal Project | Preconception; Pregnancy and birth | Physical; Pregnancy complications | Gestational weight gain; Placental weight; Gestational diabetes; Adverse birth outcomes. | Self-reported; Medical records | BMI ≥95<sup>th</sup> percentile CDC |
| Palma dos Reis | 2022<sup>109</sup> | Portugal | Pros. Obs. | Generation XXI Birth Cohort | Preconception; Pregnancy and birth | Pregnancy complications; Physical; Lifestyle | Pre-eclampsia; Anthropometrics; Smoking | Self-reported; medical report | According to the WHO criteria: class 1 (BMI 30-35), class 2 (BMI 35-40), Class 3 (BMI>40) |
| Pan | 2019<sup>50</sup> | China | Retro. Obs. |  | Pregnancy and birth | Birth anthropometrics | Adverse birth outcomes | Medical records | Weight-for-length/height Z score ≥ 95 percentiles using WHO growth charts |
| Park | 2015<sup>128</sup> | South Korea | Retro. Obs. |  | Infancy | Feeding patterns | Breastfeeding | Self-reported | 1) BMI values ≥95<sup>th</sup> percentile using an age- and gender-specific reference growth chart for Korean children, 2) children with BMI > 25 kg/m<sup>2</sup>, 3) based on percent body fat, children with ≥20% body fat for boys and ≥28% body fat for girls |
| Parker | 2012<sup>30</sup> | USA | Pros. Obs. | Project Viva | Pregnancy and birth | Pregnancy complications; Birth anthropometrics | Fetal growth; Birth weight | Medical records | BMI ≥95<sup>th</sup> percentile CDC |
| Pattison | 2019<sup>129</sup> | USA | Pros. Obs. | First Baby Study | Infancy | Feeding patterns | Breastfeeding | Self-reported | BMI ≥95<sup>th</sup> percentile CDC |
| Pei | 2014<sup>82</sup> | Germany | Pros. Obs. | Influences of Lifestyle-Related Factors on the Immune System and the Development of Allergies in Childhood plus Air | Pregnancy and birth | Pregnancy complications | Mode of delivery | Self-reported | BMI ≥95<sup>th</sup> percentile for age and sex using World Health Organization reference data |

|  |  |  |  | Pollution and Genetics (LISAplus) |  |  |  |  |  |
| --- | --- | --- | --- | --- | --- | --- | --- | --- | --- |
| Pettitt | 1983<sup>97</sup> | USA | Pros. Obs. |  | Pregnancy and birth | Pregnancy complications | Gestational diabetes | Medical records | At least 140% of their desirable weight |
| Pettitt | 1998<sup>98</sup> | USA | Pros. Obs. |  | Pregnancy and birth | Pregnancy complications | Gestational diabetes | Self-reported | Not given |
| Pitchika | 2016<sup>188</sup> | USA, Finland, Germany, Sweden | Pros. Obs. | The Environmental Determinants of Diabetes in the Young (TEDDY) | Pregnancy and birth | Pregnancy complications | Gestational diabetes | Self-reported | BMI SDS >2 according to WHO recommendations |
| Power | 2002<sup>110</sup> | UK | Pros. Obs. |  | Pregnancy and birth | Lifestyle | Smoking | Self-reported | BMI ≥90<sup>th</sup> percentile in study sample |
| Ralphs | 2021<sup>88</sup> | UK | Pros. Obs. | Born in Bradford (BiB) | Pregnancy and birth | Pregnancy complications | Mode of delivery | Medical records | BMI ≥95<sup>th</sup> percentile by UK growth charts |
| Reilly | 2005<sup>14</sup> | UK | Pros. Obs. | Avon Longitudinal Study of Parents and Children (ALSPAC) | Preconception; Pregnancy and birth; Infancy | Lifestyle; Physical; Birth anthropometrics; Feeding patterns | Smoking; Anthropometrics; Birth weight; Weight change; Introduction to solid food; Breastfeeding | Self-reported; Medical records | BMI ≥95<sup>th</sup> percentile (UK reference data) |
| Reynolds | 2014<sup>99</sup> | Ireland | Retro. Obs. | Growing Up in Ireland Study | Pregnancy and birth; Infancy | Lifestyle; Feeding patterns | Smoking; Adverse birth outcomes; Breastfeeding | Self-reported | BMI ≥95<sup>th</sup> percentile by Cole et al. |
| Rifas-Shiman | 2021<sup>89</sup> | Belarus | Intervention Study | Promotion of Breastfeeding Intervention Trial | Pregnancy and birth | Pregnancy complications | Mode of delivery | Self-reported | BMI ≥95<sup>th</sup> CDC |
| Rooney | 2011<sup>51</sup> | USA | Pros. Obs. |  | Pregnancy and birth; Infancy | Lifestyle; Physical; Birth anthropometrics; Infant anthropometrics | Smoking; Gestational weight gain; Mode of delivery; Adverse birth outcomes; Weight change | Medical records | CDC BMI ≥85<sup>th</sup> percentile |
| Rotevatn | 2021<sup>52</sup> | Denmark | Pros. Obs. |  | Preconception; Pregnancy and birth; Infancy | Sociodemographic; Environmental; Physical; Birth anthropometrics; Infant anthropometrics | Education; Income; Parity; Anthropometrics; Smoking; Mode of Delivery; Adverse birth outcomes; Weight change | Medical records | International Obesity Task Force definitions |
| Roy | 2015<sup>154</sup> | USA | Pros. Obs. | A Study of the Genetic Causes of Complex Pediatric Disorders at the Children’s Hospital of Philadelphia | Pregnancy and birth; Infancy | Pregnancy complications; Birth anthropometrics; Infant anthropometrics | Gestational diabetes; Birth weight; BMI change | Medical records | CDC BMI Z-score ≥ 95<sup>th</sup> percentile |
| Sakurai | 2022<sup>162</sup> | Japan | Pros. Obs. | The Japan Environment and Children’s Study (JECS) | Pregnancy and birth | Pregnancy complications | Prenatal antibiotics | Self-reported | BMI-for-age ≥95<sup>th</sup> percentile according to Japanese growth charts |
| Sartorius | 2020<sup>53</sup> | South Africa | Retro. Obs. | The South African National Income Dynamics Study (NA-NIDS) | Pregnancy and birth | Birth anthropometrics | Adverse birth outcomes |  | Weight-for-length z-score<br>BMI-for-age z-score of >SDS according to WHO growth charts |
| Seipel | 2013<sup>66</sup> | USA | Registry Study | National Longitudinal Survey of Youth | Pregnancy and birth; Infancy | Sociodemographic; Lifestyle; Physical; Birth anthropometrics; Feeding patterns | Age; Education; Alcohol consumption; Smoking; Drug abuse; Gestational weight gain; Mode of delivery; | Self-reported; Medical records | CDC BMI ≥95<sup>th</sup> percentile |
|  |  |  |  |  |  |  | Adverse birth outcomes;<br>Breastfeeding |  |  |
| Shankaran | 2010<sup>31</sup> | USA | Pros. Obs. | Maternal Lifestyle Study (MLS) | Pregnancy and birth | Lifestyle;<br>Physical;<br>Pregnancy complications;<br>Birth anthropometrics | Ethnicity; Education;<br>Smoking;<br>Alcohol consumption;<br>Drug use;<br>Anthropometrics;<br>Birth weight;<br>Adverse birth outcomes | Non-specified | CDC BMI $\geq 95^{\text{th}}$ percentile |
| Sharma | 2008<sup>16</sup> | USA | Registry Study | Centers for Disease Control and Prevention's Pregnancy Nutrition Surveillance System (PNSS) and Pediatric Nutrition Surveillance System (PedNSS) | Preconception;<br>Pregnancy and birth | Lifestyle | Smoking | Self-reported;<br>Medical records | BMI $\geq 95^{\text{th}}$ percentile CDC |
| Shehadeh | 2008<sup>130</sup> | Israel | Retro. Obs. | | Pregnancy and birth;<br>Infancy | Birth anthropometrics;<br>Feeding patterns | Birth weight;<br>Birth length;<br>Breastfeeding | Medical records | BMI $\geq 95^{\text{th}}$ percentile |
| Shi | 2013<sup>32</sup> | Canada | Retro. Obs. | Canadian Health Measures Survey (2007-2009) (CHMS-cycle 1) | Pregnancy and birth;<br>Infancy | Lifestyle;<br>Birth anthropometrics;<br>Feeding patterns | Smoking;<br>Birth weight;<br>Breastfeeding | Self-reported;<br>Medical records | According to WHO, BMI Z-score $>2$ SD above the mean ($\geq 97.7^{\text{th}}$ percentile) |
| Shields | 2006<sup>149</sup> | Australia | Pros. Obs. | Mater-University Study of Pregnancy cohort | Infancy | Feeding patterns | Breastfeeding | Self-reported | BMI $\geq 95^{\text{th}}$ percentile by Cole et al. (UK growth charts) |
| Si | 2022<sup>83</sup> | China | Pros. Obs. | | Pregnancy and birth | Pregnancy complications | Mode of delivery | Medical records | Weight-for-height $>3$ SD above WHO Child Growth Standards median |
| Simpson | 2017<sup>121</sup> | UK | Pros. Obs. | Avon Longitudinal Study of Parents and Children (ALSPAC) | Pregnancy and birth | Cord blood biomarkers;<br>Birth anthropometrics | Lipids;<br>Birth weight | Medical records;<br>Biochemical tests | International Obesity Task Force definitions by UK 1990 British growth reference |
| Sitarik | 2020<sup>84</sup> | USA | Pros. Obs. | Wayne County Health Environment Allergy and Asthma Longitudinal Study (WHEALS) | Pregnancy and birth | Pregnancy complications | Mode of delivery | Medical records | CDC BMI $\geq 95^{\text{th}}$ percentile |
| Skledar | 2015<sup>33</sup> | Croatia | Pros. Obs. | | Pregnancy and birth;<br>Infancy | Birth anthropometrics;<br>Feeding patterns | Birth weight;<br>Birth length;<br>Breastfeeding | Self-reported;<br>Medical records | $\geq 95^{\text{th}}$ percentile BMI-for-age growth charts CDC and Croatian Society for Paediatric Endocrinology |
| Smego | 2017<sup>155</sup> | USA | Pros. Obs. | | Infancy | Infant anthropometrics | Weight change | Medical records | CDC BMI $\geq 95^{\text{th}}$ percentile |
| Sorrow | 2019<sup>122</sup> | USA | Pros. Obs. | | Pregnancy and birth | Cord blood biomarkers | Metabolomics | Medical records | Weight-for-height Z-scores $\geq 95^{\text{th}}$ percentile CDC |
| Suzuki | 2009<sup>111</sup> | Japan | Pros. Obs. | Project Koshu | Pregnancy and birth | Lifestyle | Smoking;<br>Diet;<br>Sleep | Self-reported | Childhood obesity definition as in Cole 2000 |
| Tambalis | 2018<sup>139</sup> | Greece | Retro. Obs. |  | Infancy | Feeding patterns | Breastfeeding | Self-reported | International Obesity Task Force definitions |
| Taveras | 2009<sup>189</sup> | USA | Pros. Obs. | Project Viva | Pregnancy and birth;<br>Infancy | Birth anthropometrics;<br>Infant anthropometrics | Birth weight;<br>Birth length;<br>Weight change;<br>Height change;<br>BMI change | Medical records | BMI $\geq 95^{\text{th}}$ percentile CDC |
| Taveras | 2011<sup>156</sup> | USA | Registry Study | | Infancy | Infant anthropometrics | Weight change | Medical records | BMI for age z-score $\geq 95^{\text{th}}$ percentile CDC |
| Thaware | 2015<sup>190</sup> | UK | Pros. Obs. | Hyperglycemia and Adverse Pregnancy Outcome (HAPO) study | Pregnancy and birth | Pregnancy complications; | Gestational diabetes | Medical records; Biochemical tests | BMI z-score $\geq 95^{\text{th}}$ percentile |
| Thaware | 2018<sup>191</sup> | UK | Pros. Obs. | Hyperglycemia and Adverse Pregnancy Outcome (HAPO) study | Pregnancy and birth | Physical | Lipids | Medical records | BMI z-score $\geq 95^{\text{th}}$ percentile |
| Toschke | 2002<sup>54</sup> | Czech Republic | Registry Study | | Pregnancy and birth; Infancy | Birth anthropometrics; Feeding patterns | Adverse birth outcomes; Breastfeeding | Self-reported | BMI for age z-score $> 97^{\text{th}}$ percentile based on the study population sample |
| Toschke | 2007<sup>140</sup> | UK | Pros. Obs. | Avon Longitudinal Study of Parents and Children (ALSPAC) | Infancy | Feeding patterns | Breastfeeding | Self-reported | International Obesity Task Force definitions |
| Turner | 2021<sup>34</sup> | UK | Retro. Obs. | Aberdeen Maternity and Neonatal Databank (AMND) and Study of Trends in Obesity in North East Scotland (STONES) | Pregnancy and birth | Pregnancy complications; Birth anthropometrics | Fetal growth; Birth weight | Medical records | International Obesity Task Force definitions |
| Vafeiadi | 2015<sup>192</sup> | Greece | Pros. Obs. | The RHEA study | Pregnancy and birth | Environmental | Persistent organic pollutants exposure | Biochemical tests | International Obesity Task Force definitions |
| Vafeiadi | 2016<sup>193</sup> | Greece | Pros. Obs. | The RHEA study | Pregnancy and birth | Environmental | Endocrine-disrupting chemical exposure | Biochemical tests | International Obesity Task Force definitions |
| Van Rossem | 2011<sup>131</sup> | USA | Pros. Obs. | Project Viva | Infancy | Feeding patterns | Breastfeeding | Self-reported | BMI $\geq 95^{\text{th}}$ percentile CDC |
| Vehapoglu | 2017<sup>35</sup> | Turkey | Pros. Obs. | | Pregnancy and birth | Birth anthropometrics | Birth weight | Self-reported | BMI $\geq 95^{\text{th}}$ percentile according to Cole et al. |
| Ventura | 2020<sup>12</sup> | USA | Pros. Obs. | Infant Feeding Practices Study II (IFPS II) and Year 6 Follow-Up (Y6FU) | Preconception; Pregnancy and birth; Infancy | Physical; Sociodemographic; Birth anthropometrics; Infant anthropometrics; Feeding patterns | Prepregnancy BMI; Age; Anthropometrics; Ethnicity; Education; Marital status; Income; Parity; Birth weight; Gestational age; Introduction to solid food | Self-reported | BMI for age z-score $\geq 95^{\text{th}}$ percentile CDC |
| Von Kries | 2002<sup>36</sup> | Germany | Retro. Obs. | | Pregnancy and birth; Infancy | Lifestyle; Birth anthropometrics; Feeding patterns | Smoking; Birth weight; Introduction to solid food; Breastfeeding | Self-reported | BMI $> 97^{\text{th}}$ percentile based on European Childhood Obesity group with German reference values |
| Wallby | 2017<sup>141</sup> | Sweden | Registry study |  | Pregnancy and birth; Infancy | Sociodemographic; Physical; Feeding patterns | Age; Education; Parity Anthropometrics; Breastfeeding | Medical reports | Defined according to Cole et al. |
| Wang | 2014<sup>112</sup> | USA | Pros. Obs. | National Institute of Child Health and Human Development Study (NICHD) | Pregnancy and birth | Lifestyle | Smoking | Self-reported | BMI z-score $\geq 95^{\text{th}}$ percentile CDC |
| Wang | 2017<sup>132</sup> | USA | Pros. Obs. | National Institute of Child Health and Human Development (NICHD) Study of Early Child Care and Youth Development (SECCYD) | Infancy | Feeding patterns | Breastfeeding | Medical reports | BMI z-score $\geq 95^{\text{th}}$ percentile CDC |
| Wang | 2018<sup>164</sup> | USA | Pros. Obs. | US Collaborative Perinatal Project | Pregnancy and birth | Pregnancy complications | Prenatal antibiotics | Self-reported | BMI z-score $\geq 95^{\text{th}}$ percentile CDC |
| Wang | 2022<sup>37</sup> | China | Retro. Obs. |  | Preconception;<br>Pregnancy and birth | Birth anthropometrics;<br>Physical | Prepregnancy BMI;<br>Gestational weight gain;<br>Birth weight | Self-reported | For children below 5 BMI Z-score > 3SD of WHO growth standards; for children above 5 BMI z-score > 2SD than reference median; IOTF criteria for children 2-18yrs, obesity as over BMI of 30 kg/m2; China criteria obesity as BMI over 28 kg/m2 |
| Warner | 2013<sup>194</sup> | USA | Pros. Obs. | The Health Assessment of Mothers and Children of Salinas (CHAMACOS) study | Pregnancy and birth | Environmental | Pesticide exposure | Biochemical tests | BMI $\geq 95^{\text{th}}$ percentile CDC |
| Weber | 2014<sup>167</sup> | Belgium<br>Germany<br>Italy<br>Poland<br>Spain | Intervention study | Childhood Obesity Project | Infancy | Feeding patterns | Formula feeding | Medical records | International Obesity Task Force definitions |
| Wen | 2022<sup>120</sup> | USA | Pros. Obs. | Infant Feeding Practices Study II | Pregnancy and birth | Sociodemographic | Education;<br>Income | Self-reported | BMI for age z-score $\geq 95^{\text{th}}$ percentile CDC |
| Whitaker | 1998<sup>195</sup> | USA | Pros. Obs. | | Preconception;<br>Pregnancy and birth | Physical; Pregnancy complications; | Anthropometrics;<br>Gestational diabetes | Medical records | BMI Z-scores $\geq 85^{\text{th}}$ percentile |
| Widen | 2016<sup>74</sup> | USA | Pros. Obs. | Colombia Center for Children's Environmental Health Mothers and Newborns Study | Preconception;<br>Pregnancy and birth | Physical | Anthropometrics;<br>Gestational weight gain | Self-reported | BMI for age z-score $\geq 95^{\text{th}}$ percentile CDC |
| Wojcicki | 2015<sup>13</sup> | USA | Retro. Obs. | Pregnancy Risk Assessment Monitoring System | Preconception;<br>Pregnancy and birth;<br>Infancy | Sociodemographic;<br>Physical;<br>Lifestyle<br>Feeding patterns | Age; Ethnicity;<br>Education;<br>Income; Smoking;<br>Marital status;<br>Anthropometrics;<br>Breastfeeding | Self-reported | BMI $\geq 95^{\text{th}}$ percentile (age and sex-specific) |
| Wright | 2009<sup>196</sup> | USA | Pros. Obs. | Project Viva | Pregnancy and birth | Pregnancy complications | Gestational diabetes | Medical records | BMI >95<sup>th</sup> percentile CDC |
| Wroblewska-Senuik | 2009<sup>197</sup> | Poland | Pros. Obs. | | Pregnancy and birth | Pregnancy complication | Gestational diabetes | Self-reported | BMI $\geq 95^{\text{th}}$ percentile for gender and age based on Polish reference growth charts |
| Yamakawa | 2013<sup>133</sup> | Japan | Pros. Obs. | Longitudinal Survey of Babies in the 21st Century | Infancy | Feeding Patterns | Breastfeeding | Self-reported | International Obesity Task Force definitions |
| Yuan | 2016<sup>85</sup> | USA | Pros. Obs. | the Growing Up Today Study | Pregnancy and birth | Pregnancy complications | Mode of delivery | Self-reported | International Obesity Task Force definitions |
| Zarrati | 2013<sup>57</sup> | Iran | Retro. Obs. |  | Pregnancy and birth | Birth anthropometrics | Birth weight;<br>Adverse birth outcomes | Self-reported | Age- and sex-specific BMI >95<sup>th</sup> percentile CDC |
| Zhang | 2022<sup>198</sup> | China | Registry study | Tianjin mother-child cohort | Preconception | Physical | Pre-pregnancy weight | Medical records | BMI $\geq 95^{\text{th}}$ percentile of WHO growth standards |
| Zhou | 2011<sup>38</sup> | China | Retro. Obs. |  | Pregnancy and birth;<br>Infancy | Pregnancy complication;<br>Birth anthropometrics;<br>Lifestyle;<br>Feeding Patterns; | Prenatal music exposure;<br>Mode of delivery;<br>Birth weight;<br>Introduction to solid food | Self-reported | International Obesity Task Force definitions |

### Risk factors for childhood obesity in preconception

We identified 34 observational studies and no RCTs that assessed 6 risk factors associated with childhood obesity in the preconception period ((**Figure 3A_1_&3B_1_, Table S2)**. Maternal physical factors were most frequently studied. 19 observational studies reported that higher maternal preconception weight was associated with an increased risk of childhood obesity, whereas 3 observational studies^11–13^ found no association. Samples sizes varied from 205 to 5156. All identified studies showed that maternal prepregnancy overweight or obesity was associated with a higher risk of childhood obesity. Only 1 study^14^ reported paternal overweight or obesity as risk factor for childhood obesity. 4 observational studies^15–18^ focused on the associations of maternal lifestyle factors during preconception with childhood obesity risk. 2 observational studies^15,16^ showed that maternal smoking in the preconception period was associated with an increased risk of childhood obesity, whereas no associations with the risk of childhood obesity were reported for maternal preconception diet^15,17^ or physical activity^15,18^.

**Figure 3A.**
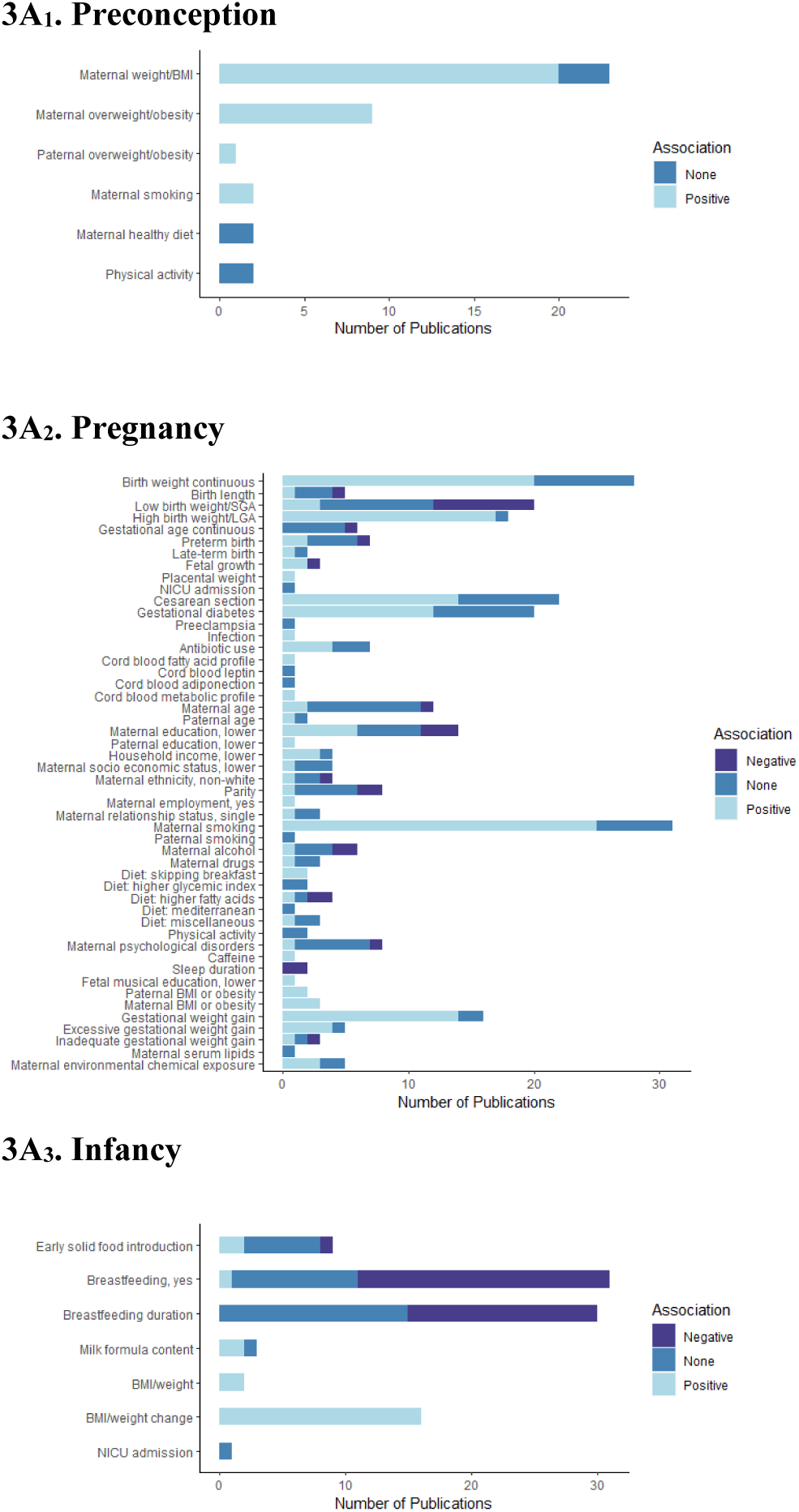
Number of positive, negative, and null associations for each identified exposure in all included publications.

**Figure 3B.**
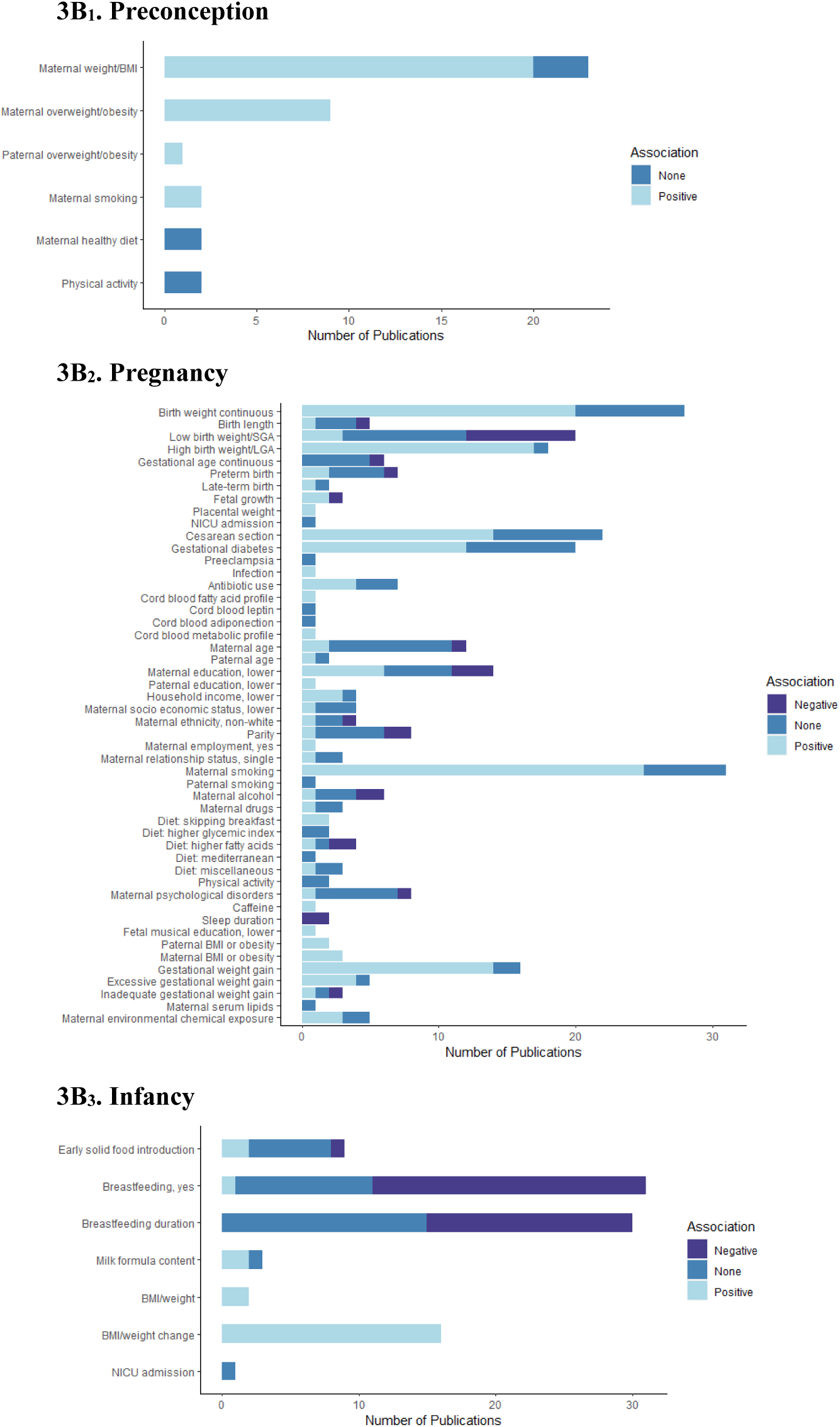
Number of positive, negative, and null associations for each identified exposure within only moderate to high quality publications.

### Risk factors for childhood obesity during pregnancy

We identified 135 observational studies and 2 RCTs that assessed 46 different risk factors during pregnancy (**Figure 3A_2_&3B_2_, Table S2)**. Most frequently studied characteristics were birth characteristics and parental lifestyle factors. Most risk factors were assessed in mother and child. Only 4 studies reported paternal data^13,19–21^.

The most commonly reported risk factor was birthweight, assessed continuously, with 20^12,14,19,22–38^ of 28 observational studies reporting that a higher birthweight was associated with a higher risk for childhood obesity, and 17^20,39–54^ of 18 observational studies identifying large for gestational age (LGA) as a risk factor for childhood obesity. For small for gestational age (SGA) at birth, findings were less consistent. 4 observational studies^31,55–57^ identified SGA as a risk for childhood obesity, however, eight observational studies^11,39,49,52,53,58–60^ reported that SGA was associated with a lower risk of childhood obesity. Most observational studies assessing gestational age^12,29,61–63^ at birth and or pre- and late-term birth^26,64–66^ reported no evidence for higher risk for childhood obesity. 29 observational studies focused on parental physical factors. The majority investigated the association of (excessive) maternal gestational weight gain with childhood obesity, 14^26,37,51,60,61,66–74^ of 16 observational studies reported an increased risk of childhood obesity associated with higher gestational weight gain.

Most studies on pregnancy complications have focused on mode of delivery and gestational diabetes. Within 14^51,52,61,75–85^ out of 22 studied populations, caesarean section (CS) was identified as a risk factor for increased childhood obesity. 7 observational studies^11,38,66,77,86–88^ and 1 RCT^89^ reported no association between CS and childhood obesity. Gestational diabetes was identified as risk factor for childhood obesity in 12 out of 21 observational studies^49,61,78,90–98^ . 25^11,14,16,26,29,32,36,51,52,66,78,99–112^ out of 31 observational studies showed that maternal smoking during pregnancy was associated with a higher risk for childhood obesity. 10 observational studies^23,107,108,111,113–118^ and 1 RCT^119^ on maternal diet during pregnancy reached no consensus on the association with childhood obesity, partly because of the diversity of diets investigated. No consistent evidence was found for maternal age, socioeconomic status, ethnicity, parity, employment, relationship status, maternal or paternal education, and paternal age as risk factors for childhood obesity. Lower household income^13,52,120^ was reported within 3 out of 4 studies as risk factor for obesity in childhood.

Only 3 publications^114,121,122^ explored different cord blood biomarkers in relation to childhood obesity. Risk factors were fatty acids^114^, leptin and adiponectin^121^ and differences in metabolomics^122^, and no consensus on associations with childhood obesity was reached.

### Risk factors for childhood obesity during infancy

We identified 76 observational studies and 3 RCTs that assessed 7 different exposures during infancy (**Figure 3A_3_&3B_3_**). The most frequently studied characteristic was feeding pattern. All risk factors were assessed in the child. 20^11,32,36,39,54,60,66,70,78,123–133^ out of 30 observational studies on breastfeeding reported that a breastfed child had lower risks of obesity as compared to a non-breastfed child. Furthermore, 15 observational studies^13,26,63,99,102,124,131,134–141^ reported that longer breastfeeding duration was associated with less childhood obesity, however 2 RCTs^142,143^ and 13 observational studies^12,23,33,62,104,129,132,144–149^ did not report an effect of the duration of breastfeeding on childhood obesity. All 16 observational studies^12,29,45,51,52,58–60,65,150–156^ focusing on body mass index (BMI) or weight gain during infancy reported that higher BMI or weight gain was associated with a higher risk of childhood obesity.

### Risk of bias assessment for quality and synthesis

Level of evidence was generally scored as moderate quality, mainly due suboptimal assessment of the exposure or outcome, inadequate blinding of participants and researchers in RCTs, high risk of residual confounding, short duration of follow-up, and high loss-to-follow up (**Figure 4**). 6% of included studies was of low quality, consisting of 1 RCT and 10 cohort studies. **Supplemental figures 1A-C--3A-D** show the detailed bias assessments for all included studies based on the JBI checklist.

**Figure 4.**
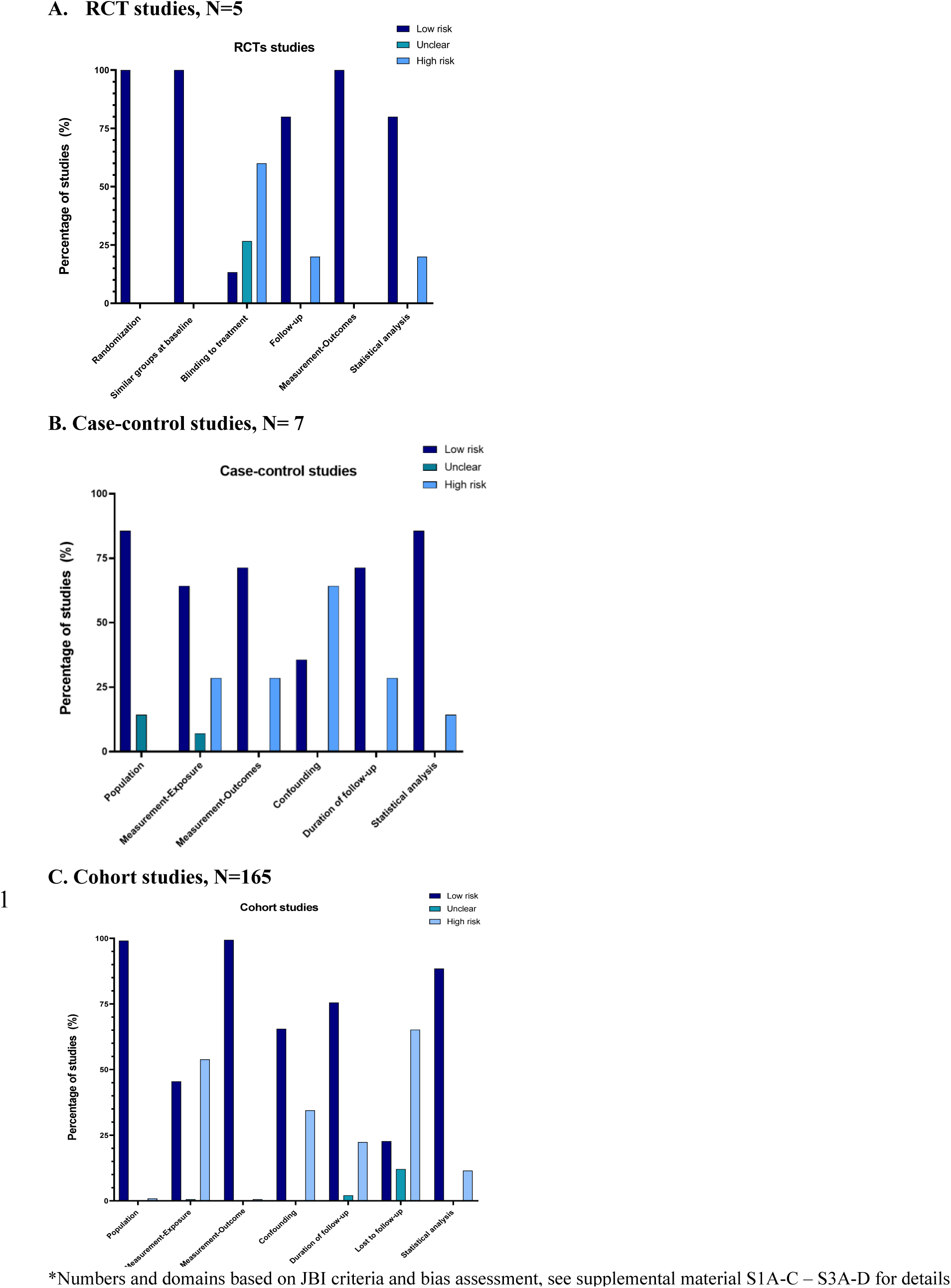
**Risk for bias in different domains\***

### Quality assessment of risk factors for prediction and prevention

In total, we identified 24 risk factors consistently associated with childhood obesity, which included: maternal prepregnancy/pregnancy weight/overweight/obesity (hereafter maternal prepregnancy and pregnancy weight status), prepregnancy/pregnancy smoking, caesarean section, gestational diabetes, antibiotic use, education, household income, sleep duration, gestational weight gain/excessive gestational weight gain (hereafter maternal gestational weight gain), environmental chemical exposure, paternal prepregnancy/pregnancy weight/overweight/obesity, continuous foetal growth, birthweight/high birthweight/LGA, breastfeeding/duration of breastfeeding, milk formula content, infant BMI/weight/and BMI/weight gain.

We evaluated these risk factors for their potential for prediction and prevention strategies (**Figure S4)**. In preconception, only maternal weight status scored very strong on its predictability and strong on modifiability. In pregnancy, maternal smoking, weight status, gestational weight gain, and birthweight/LGA scored strong or higher on prediction and modifiability. In infancy, only breastfeeding and infant BMI/weight change scored strong or very strong for their potential as predictor and to be modifiable on individual level.

## DISCUSSION

In this systematic review, we identified 172 observational and 5 intervention studies from 37 countries that studied risk factors for childhood obesity in the first 1000 days of life. Most studies were performed in high-income countries and focused on identification of risk factors in pregnancy. 24 risk factors were consistently associated with the risk of childhood obesity. Higher maternal prepregnancy weight, higher gestational weight gain, maternal smoking during pregnancy, higher birthweight, LGA, no breastfeeding, and higher infant weight gain were the strongest risk factors for childhood obesity and need to be considered in future prediction or prevention strategies. Overall level of evidence was generally moderate due to unreliable measurements of exposures, short follow-up duration and loss to follow-up, and risk of residual confounding.

This current review provides the most comprehensive, systematic, up-to-date information on family-based risk factors for childhood obesity from preconception until infancy, thereby covering the full first 1000 days of life.^157^ Due to the large heterogeneity between studies, we could not perform meta-analyses. Despite increasing awareness on the importance of the preconception period for offspring health, only a remarkable low number of studies focused on risk factors in this potential critical period. Maternal prepregnancy weight status was consistently associated with childhood obesity, but no consistent effects were found for other maternal lifestyle factors. The transgenerational effects of increased maternal prepregnancy weight on childhood obesity risk poses a major public health concern, as obesity often progresses into adulthood, subsequently causing increased risks for the next generation.^2^ Only 1 publication focused on paternal factors and reported that higher paternal prepregnancy BMI was associated with a higher childhood obesity risk . Adverse paternal sociodemographic and lifestyle factors have already been shown to be associated with adverse pregnancy outcomes.^158^ Animal studies have shown that these paternal factors also increase the risk of childhood obesity, with the strongest effects for paternal obesity, but also for paternal hyperglycaemia and paternal smoking.^159^

As expected, pregnancy was the most studied period for risk factors for childhood obesity. Most studies focused on maternal and foetal risk factors. Suboptimal maternal gestational weight status and gestational weight gain were consistently associated with the risk of childhood obesity. Gestational diabetes and gestational hypertensive disorders tended to be associated with a higher risk of childhood obesity, but this was not consistent in moderate to high quality studies. Indeed, a previous individual-participant-data meta-analysis among 160757 mother-offspring pairs from European and North-American birth cohorts, also reported that no effect of gestational diabetes or gestational hypertensive disorders was present on childhood obesity after adjustment for maternal prepregnancy BMI.^160^ A higher birthweight and LGA were strong risk factors for childhood obesity, whereas associations for SGA and preterm birth were inconclusive. Children born SGA may be constitutionally small or have suffered from intra-uterine foetal growth restriction. Differences in this pathophysiology may partly explain these inconsistent findings.^161^ A lower household income and lower maternal and paternal education level tended to be associated with a higher risk of childhood obesity, but only inconsistent in less than 5 studies of at least moderate quality. As these findings are less consistent, generalizability is more difficult. Possibly, these effects are more country-specific and influenced by community and social support practices per region. Several moderate and high quality studies reported an association between CS and a higher risk for childhood obesity. All were observational studies, and the only included RCT did not support that association, suggesting that residual confounding may play a role in these observed associations. Maternal antibiotics during pregnancy was associated with childhood obesity in 50% of studies, but the number of studies was low, and associations were only found in females^162^, or not in all investigated age groups^163,164^, so no generalized conclusion can be drawn. No consistent associations were present for maternal or paternal age, ethnicity or parity.

In infancy, infant weight gain and feeding patterns were mostly studied with regards to the development of childhood obesity. Strongest associations were present for higher infant weight or BMI gain with higher risks of obesity. The infant period reflects the greatest proportional weight gain in postnatal life and might be a critical period for development of energy balance mechanisms. Early exposure to excess and dysfunctional fat mass might trigger the development of metabolic changes in childhood.^165^ Additionally, early weight gain may reflect the beginning influence of a genetic predisposition to overweight.^166^ Being breast-fed and a longer duration of breastfeeding were associated with lower risks of childhood obesity within the majority of reported observational studies. The only two intervention studies^142,143^, did not identify a significant association between the duration of breastfeeding and childhood obesity. Causality between the protective role of breastfeeding on development of childhood obesity can therefore not be properly determined based on these findings. It is remarkable that we identified no studies on the role of different components of breastmilk and only 3 studies on differences in infant formula composition.^125,137,167^

Our findings are important from an etiological perspective and provide novels insights to improve prediction and prevention strategies from the start of life onwards. Improved risk selection from early-life onwards is crucial to enable more targeted delivery of prevention strategies. Prevention strategies focused on improving these identified early-life risk factors from a clinical and population perspective is needed targeting those families at higher risk of offspring obesity, preferable integrated with implementation studies to evaluate risk selection, intervention effects and optimize delivery methods. The first 1000 days of life offer an are unique opportunity for prevention, because of parental motivation and contact with the health care system.^168,169^

Our systematic review also identified major areas for further research. First, studies were conducted in mainly high-income, countries. To increase global generalizability, more studies should be performed in low- and middle income countries, in which the rise of childhood obesity is most rapidly increasing and prevalence of identified risk factors may vary as compared to higher income countries.^2^ Second, we only identified few studies in preconception and infancy, whereas both periods are increasingly recognized as critical periods for childhood obesity development.^170^ Additionally, limited number of studies focused on paternal factors, environmental factors, metabolomics, and genetic factors. Evidence is increasing on the role of environmental exposures in human health, and investigation of their role in development of childhood obesity is warranted, as they are ubiquitous and represent an opportunity for governmental and political intervention. The field of omics is expanding rapidly, and further insight in the value of metabolomics and genomics for prediction of childhood obesity is of interest. Third, our findings are mainly based on observational studies, and causality cannot be established from observational evidence.^171^ Quality assessment showed many studies were at risk of residual confounding. We identified only 5 intervention studies, which included childhood obesity as outcome. 1 RCT investigated the role of maternal low glycaemic index diet during pregnancy^119^, 2 RCT’s investigated the duration of breastfeeding^142,143^ and 1 RCT investigated the difference between infant formula with high protein and low protein^167^. Especially in pregnancy, maternal lifestyle interventions have been conducted in RCTs, but these studies often did not include childhood obesity as a primary or secondary outcome and suffered from high attrition. Our systematic review underlines the urgent importance of well-designed intervention studies targeting parental lifestyle from preconception onwards with longer follow-up of the offspring and inclusion of clinical cardiometabolic outcomes.

## CONCLUSION

We showed that 24 risk factors in early life are consistently associated with a higher risk of childhood obesity. Higher maternal prepregnancy weight and gestational weight gain, maternal smoking during pregnancy, higher birthweight and large-size-for-gestational-age-at-birth, no breastfeeding, and higher infant weight gain are most strongly related to childhood obesity risk. These findings are relevant for early life risk prediction and insight into the strongest potential modifiable factors from a clinical, governmental, and industrial perspective.

## Data Availability

All data produced in the present study are available upon reasonable request to the authors

## ACKNOWLEDGEMENTS

RG received funding from the Netherlands Organization for Health Research and Development (ZonMW, grant number 543003109) and funding from the European Union’s Horizon 2020 research and innovation program under the ERA-NET Cofund action (No. 727565), European Joint Programming Initiative “A Healthy Diet for a Healthy Life” (JPI HDHL), EndObesity, ZonMW Netherlands (No. 529051026). SMB was supported by the KNAW Ter Meulen Grant/KNAW Medical Science Fund, Royal Netherlands Academy of Arts & Sciences, and by ILSI Europe. ASJK was supported by ISLI Europe. MCC acknowledges the support by the Spanish Ministry of Science and Innovation (MCIN) research grant (ref. PID2022-139475OB-I00) and the award from MCIN/AEI to the Institute of Agrochemistry and Food Technology (IATA-CSIC) as Centre of Excellence Severo Ochoa (CEX2021-001189-S MCIN/AEI /10.13039/ 501100011033). EFV was supported by a Predoctoral grant awarded by the Spanish Ministry of Science and Innovation and the European Social Fund Plus (ESF+) for the training of doctors within the framework of the State Plan for Scientific, Technical and Innovation Research 2021 - 2023. (ref. CEX2021-001189-S-20-1). FJRO was supported by the Spanish Goverment (Juan de la Cierva programm, Ministry of Science and Innovation). This Project was executed in collaboration with Early Nutrition and Long-term Health Task Force of the European Branch of the International Life Sciences Institute (ILSI Europe). Industry members of this task force are listed on the website www.ilsi.eu. For more information, please contact ILSI Europe by or call +32 2 771.00.14. The opinions expressed herein, and the conclusion of the project are those of the authors and do not necessarily represent the view of ILSI Europe nor those of its member companies.

## Funding

This work was conducted by an expert group of the European branch of the International Life Sciences Institute, ILSI Europe. This publication was coordinated by the Early Nutrition and Long- term Health Task Force. Industry members of this task force are listed on the ILSI Europe website at https://ilsi.eu/scientific-activities/nutrition/early-nutrition-and-long-term-health/. Experts are not paid for the time spent on this work. However, the nonindustry members within the expert group were offered support for travel on costs from the Early Nutrition and Long-term Health Task Force to attend 1 live meeting to discuss the manuscript. The expert group carried out the work, that is collecting/analysing data/information and writing the scientific paper separate to other activities of the task force. The research reported is the result of a scientific evaluation in line with ILSI Europe’s framework to provide a precompetitive setting for public-private partnership. ILSI Europe facilitated scientific meetings and coordinated the overall project management and administrative tasks relating to the completion of this work. For further information about ILSI Europe, please or call +3227710014. The opinions expressed herein and the conclusions of this publication are those of the authors and do not necessarily represent the views of ILSI Europe nor those of its member companies or any regulatory authority.

## AUTHOR CONTRIBUTIONS

SMB: Drafted the initial version of the manuscript, completed the review, extraction, and quality assessment of papers.

ASJK: Drafted the initial version of the manuscript, completed the review, extraction, and quality assessment of papers.

FJRO: Completed the review, extraction, and quality assessment of papers, provided critical intellectual feedback.

MBH: provided extraction of the data and provided critical intellectual feedback and completed quality assessment of papers.

EFV: provided extraction of the data and provided critical intellectual feedback. MAB: provided extraction of the data and provided critical intellectual feedback. MCC: provided extraction of the data and provided critical intellectual feedback.

JvD: Set up the initial aim of the study and study protocol, provided extraction of the data and provided critical intellectual feedback.

PI: provided extraction of the data and provided critical intellectual feedback. KK: provided extraction of the data and provided critical intellectual feedback.

CAvLB: provided extraction of the data and provided critical intellectual feedback.

AG: Completed the review, extraction, and quality assessment of papers and provided critical intellectual feedback.

RG: Set up the initial aim of the study and study protocol, completed the review, extraction, and quality assessment of papers, drafted the initial version of the manuscript.

All authors approved the final version for publication.

## COMPETING INTERESTS STATEMENT

All authors declare no competing interests.

