## Supplemental material for "Risk factors in the first 1000 days of life associated with childhood obesity: a systematic review and risk factor quality assessment"

**Text S1.** Search terms for the systematic review.

**Table S1.** Guideline on quality assessment of risk factors.

**Table S2.** Associations of childhood obesity with risk factors per period.

**Figure S1-3.** Bias assessment of the included studies.

**Figure S4.** Quality assessment of the 24 consistently associated risk factors with childhood obesity.

**Text S1.** Search terms for the systematic review,

**EMBASE**

**https://www.embase.com/#advancedSearch/default – 10659 records**

('pregnancy'/exp OR 'pregnancy' OR 'pregnant woman'/exp OR 'pregnant woman' OR 'mother'/exp OR 'mother' OR 'prenatal exposure'/exp OR 'prenatal exposure' OR 'pregnancy outcome'/exp OR 'pregnancy outcome' OR 'pregnancy disorder'/exp OR 'pregnancy disorder' OR 'pregnancy complication'/exp OR 'pregnancy complication' OR 'high-risk pregnancy'/exp OR ‘adolescent pregnancy’/exp OR ‘maternal smoking’/exp OR 'prenatal period'/exp OR 'prenatal period' OR 'prenatal growth'/exp OR 'prenatal growth' OR 'fetus disease'/exp OR 'fetus disease' OR 'newborn disease'/exp OR 'newborn disease' OR 'low birth weight'/exp OR 'low birth weight' OR 'umbilical cord blood'/exp OR 'umbilical cord blood' OR 'infant disease'/exp OR 'infant disease' OR 'adolescent pregnancy'/exp OR 'adolescent pregnancy' OR gravidit*:ab,ti,kw OR gestation*:ab,ti,kw OR pregnan*:ab,ti,kw OR mother*:ab,ti,kw OR prenatal*:ab,ti,kw OR preeclamp*:ab,ti,kw OR 'pre eclamp*':ab,ti,kw OR (((prematur* OR preterm* OR live*) NEAR/3 (labor OR birth OR labour OR delivery OR ruptur*)):ab,ti,kw) OR birthweight*:ab,ti,kw OR (((birth OR neonat* OR newborn OR infant*) NEAR/3 weight*):ab,ti,kw) OR (((infant* OR child*) NEAR/3 ('breast feed*' OR breastfeed* OR 'milk composit*' OR weaning)):ab,ti,kw) OR ((infant* NEAR/3 (biomarker* OR saliva OR fecal OR urine OR hair)):ab,ti,kw) OR postpartum:ab,ti,kw OR 'post partum':ab,ti,kw OR puerperium:ab,ti,kw OR (((fetal OR foetus OR fetus OR newborn OR neonat* OR infant* OR intrauterin* OR 'intra uterin*') NEAR/3 (diseas* OR abnormal* OR growth* OR infect*)):ab,ti,kw) OR ((gestational NEAR/3 (diabet* OR hypertens* OR weight*)):ab,ti,kw) OR ((placenta* NEAR/3 abruption):ab,ti,kw) OR ((('small for date' OR 'small for gestational age' OR 'large for date' OR 'large for gestational age') NEAR/3 infant*):ab,ti,kw) OR preconcept*:ab,ti,kw OR maternal*:ab,ti,kw OR paternal*:ab,ti,kw OR father*:ab,ti,kw OR parent*:ab,ti,kw OR 'cord blood*':ab,ti,kw OR 'fetal blood*':ab,ti,kw OR lifestyle*:ab,ti,kw OR sociodemographic*:ab,ti,kw) **AND** ('follow up'/exp OR 'follow up' OR 'longitudinal study'/exp OR 'longitudinal study' OR 'prospective study'/exp OR 'prospective study' OR 'retrospective study'/exp OR 'retrospective study' OR 'cohort analysis'/exp OR 'cohort analysis' OR 'prediction'/exp OR 'prediction' OR followup*:ab,ti,kw OR 'follow up*':ab,ti,kw OR longitudinal*:ab,ti,kw OR prospective*:ab,ti,kw OR retrospective*:ab,ti,kw OR cohort*:ab,ti,kw OR predict*:ab,ti,kw) **AND** ((((obes* OR hypertension* OR diabetes* OR diabetic* OR bmi OR 'body mass*' OR 'waist circum*' OR 'fat distribut*' OR dyslipidemia* OR lipidemia OR lipoedema OR 'glucos* intoleranc*' OR 'glucos* toleranc*' OR overweight* OR overnutrition*) NEAR/6 (child* OR progeny OR offspring*)):ab,ti,kw) OR (((metabolic* OR cardiometabolic* OR 'insulin resist*' OR dysmetabolic* OR metabolism*) NEAR/2 (syndrom* OR diseas* OR outcome* OR disorder*) NEAR/9 (child* OR progeny OR offspring*)):ab,ti,kw) OR (((increas* OR elevat* OR high*) NEAR/2 (bloodpress* OR 'blood press*') NEAR/9 (child* OR progeny OR offspring*)):ab,ti,kw)) **NOT** (('animal'/exp OR animal OR animal*:de OR 'nonhuman'/exp OR nonhuman) NOT ('human'/exp OR 'human')) AND [english]/lim NOT ([conference abstract]/lim OR [preprint]/lim) NOT ('case report'/exp OR 'case report' OR 'case report*':ti)

**Medline**

**https://ovidsp.dc1.ovid.com/ovid-b/ovidweb.cgi – 11364 results.**

(exp Pregnancy/ OR Pregnant Women/ OR exp Parents/ OR exp Pregnancy Complications/ OR Pregnancy Outcome/ OR exp Infant, Newborn, Diseases/ OR exp Infant, Low Birth Weight/ OR Fetal Blood/ OR exp Pregnancy, High-Risk/ OR exp Pregnancy in Adolescence/ OR exp Biomarkers/ OR exp Cardiometablic Risk Factors/ OR exp sociodemographic factors/ OR exp maternal exposure/ OR exp Feeding Behavior/ OR exp Infant, Newborn, Disease/ OR (gravidit* OR gestation* OR pregnan* OR prenatal* OR maternal* OR preeclamp* OR pre-eclamp* OR ((prematur* OR preterm* OR live*) ADJ3 (labor OR birth OR labour OR delivery OR ruptur*)) OR birthweight* OR ((birth OR neonat* OR newborn OR infant*) ADJ3 (weight*)) OR ((infant* OR child*) ADJ3 (breast-feed* OR breastfeed* OR milk-composit* OR weaning)) OR ((infant*) ADJ3 (biomarker* OR saliva OR fecal OR urine OR hair)) OR postpartum OR post-partum OR puerperium OR ((fetal OR foetus OR fetus OR newborn OR neonat* OR infant* OR intrauterin* OR intra-uterin*) ADJ3 (diseas* OR abnormal* OR growth* OR infect*)) OR ((gestational) ADJ3 (diabet* OR hypertens* OR weight*)) OR ((placenta*) ADJ3 (abruption)) OR ((small-for-date OR small-for-gestational-age OR large-for-date OR large-for-gestational-age) ADJ3 (infant*)) OR preconcept* OR maternal* OR paternal* OR father* OR parent* OR cord-blood* OR fetal-blood* OR lifestyle* OR sociodemographic*).ab,ti,kf.) **AND** (exp Cohort Studies/ OR (followup* OR follow-up* OR longitudinal* OR prospective* OR retrospective* OR cohort* OR predict*).ab,ti,kf.) **AND** (exp Pediatric Obesity/ OR (((obes* OR hypertension* OR diabetes* OR diabetic* OR BMI OR body-mass* OR waist-circum* OR fat-distribut* OR dyslipidemia* OR lipidemia OR lipoedema OR glucos*-intoleranc* OR glucos*-toleranc* OR overweight* OR overnutrition*) ADJ6 (child* OR progeny OR offspring*)) OR ((metabolic* OR cardiometabolic* OR insulin-resist* OR dysmetabolic* OR metabolism*) ADJ2 (syndrom* OR diseas* OR outcome* OR disorder*) ADJ9 (child* OR progeny OR offspring*)) OR ((increas* OR elevat* OR high*) ADJ2 (bloodpress* OR blood-press*) ADJ9 (child* OR progeny OR offspring*))).ab,ti,kf.) NOT (exp animals/ NOT humans/) AND (english).lg NOT (news OR congres* OR abstract* OR book* OR chapter* OR dissertation abstract*).pt. NOT (Case Reports/ OR case-report*.ti.)

**Web of Science**

**https://www.webofscience.com/wos/woscc/advanced-search - 10034 records**

TS=(((gravidit* OR gestation* OR pregnan* OR mother* OR prenatal* OR preeclamp* OR pre-eclamp* OR ((prematur* OR preterm* OR live*) NEAR/2 (labor OR birth OR labour OR delivery OR ruptur*)) OR birthweight* OR ((birth OR neonat* OR newborn OR infant*) NEAR/2 (weight*)) OR ((infant* OR child*) NEAR/2 (breast-feed* OR breastfeed* OR milk-composit* OR weaning)) OR ((infant*) NEAR/2 (biomarker* OR saliva OR fecal OR urine OR hair)) OR postpartum OR post-partum OR puerperium OR ((fetal OR foetus OR fetus OR newborn OR neonat* OR infant* OR intrauterin* OR intra-uterin*) NEAR/2 (diseas* OR abnormal* OR growth* OR infect*)) OR ((gestational) NEAR/2 (diabet* OR hypertens* OR weight*)) OR ((placenta*) NEAR/2 (abruption)) OR ((small-for-date OR small-for-gestational-age OR large-for-date OR large-for-gestational-age) NEAR/2 (infant*)) OR preconcept* OR maternal* OR paternal* OR father* OR parent* OR cord-blood* OR fetal-blood* OR lifestyle* OR sociodemographic*)) AND ((followup* OR follow-up* OR longitudinal* OR prospective* OR retrospective* OR cohort* OR predict*)) AND ((((obes* OR hypertension* OR diabetes* OR diabetic* OR BMI OR body-mass* OR waist-circum* OR fat-distribut* OR dyslipidemia* OR lipidemia OR lipoedema OR glucos*-intoleranc* OR glucos*-toleranc* OR overweight* OR overnutrition* ) NEAR/5 (child* OR progeny OR offspring*)) OR ((metabolic* OR cardiometabolic* OR insulin-resist* OR dysmetabolic* OR metabolism*) NEAR/2 (syndrom* OR diseas* OR outcome* OR disorder*) NEAR/9 (child* OR progeny OR offspring*)) OR ((increas* OR elevat* OR high*) NEAR/2 (bloodpress* OR blood-press*) NEAR/9 (child* OR progeny OR offspring*)))) NOT ((animal* OR rat OR rats OR mouse OR mice OR murine OR dog OR dogs OR canine OR cat OR cats OR feline OR rabbit OR cow OR cows OR bovine OR rodent* OR sheep OR ovine OR pig OR swine OR porcine OR veterinar* OR chick* OR zebrafish* OR baboon* OR nonhuman* OR primate* OR cattle* OR goose OR geese OR duck OR macaque* OR avian* OR bird* OR fish*) NOT (human* OR patient* OR women OR woman OR men OR man))) AND LA=(English) AND DT=(Article OR Review OR Letter OR Early Access)

**Scopus**

**Scopus - Advanced search – 1631 records**

( TITLE-ABS-KEY ( ( gravidit* OR gestation* OR pregnan* OR mother* OR prenatal* OR preeclamp* OR pre-eclamp* OR ( ( prematur* OR preterm* OR live* ) PRE/3 ( labor OR birth OR labour OR delivery OR ruptur* ) ) OR birthweight* OR ( ( birth OR neonat* OR newborn OR infant* ) PRE/3 ( weight ) ) OR ( ( infant* OR child* ) PRE/3 ( breast-feed* OR breastfeed* OR milk-composit* OR weaning ) ) OR ( ( infant* ) PRE/3 ( biomarker* OR saliva OR fecal OR urine OR hair ) ) OR postpartum OR post-partum OR puerperium OR ( ( fetal OR foetus OR fetus OR newborn OR neonat* OR infant* OR intrauterin* OR intra-uterin* ) PRE/3 ( diseas* OR abnormal* OR growth* OR infect* ) ) OR ( ( gestational ) PRE/3 ( diabet* OR hypertens* OR weight* ) ) OR ( ( placenta* ) PRE/3 ( abruption ) ) OR ( ( small-for-date OR small-for-gestational-age OR large-for-date OR large-for-gestational-age ) PRE/3 ( infant* ) ) OR preconcept* OR maternal* OR paternal* OR father* OR parent* OR cord-blood* OR fetal-blood* OR lifestyle* OR sociodemographic* ) ) ) AND ( TITLE-ABS-KEY ( "follow up" OR "longitudinal AND study" OR "prospective AND study" OR "retrospective AND study" OR "cohort AND analysis" OR "prediction" OR ( followup* OR follow-up* OR longitudinal* OR prospective* OR retrospective* OR cohort* OR predict* ) ) ) AND ( TITLE-ABS-KEY ( ( ( obes* OR hypertension* OR diabetes* OR diabetic* OR bmi OR body-mass* OR waist-circum* OR fat-distribut* OR dyslipidemia* OR lipidemia OR lipoedema OR glucos*-intoleranc* OR glucos*-toleranc* OR overweight* OR overnutrition*) PRE/6 ( child* OR progeny OR offspring* ) ) OR ( metabolic* OR cardiometabolic* OR insulin-resist* OR dymetabolic* OR metabolism* ) PRE/2 ( syndrom* OR diseas* OR outcome* OR disorder* ) PRE/9 ( child* OR progeny OR offspring* ) ) OR ( ( increas* OR elevat* OR high* ) PRE/2 ( bloodpress* OR blood-press* ) PRE/9 ( child* OR progeny OR offspring* ) ) ) ) AND

NOT ( ( INDEXTERMS ( animals OR animal ) ) AND NOT ( INDEXTERMS ( humans OR human ) ) ) AND ( LIMIT-TO ( LANGUAGE,"English" ) ) AND ( EXCLUDE ( DOCTYPE,"cp" ) OR EXCLUDE ( DOCTYPE,"ch" ) OR EXCLUDE ( DOCTYPE,"le" ) )

**Cochrane**
**https://www.cochranelibrary.com/advanced-search/search-manager - 1896 records**
((gravidit* OR gestation* OR pregnan* OR mother* OR prenatal* OR preeclamp* OR pre NEXT/1 eclamp* OR ((prematur* OR preterm* OR live*) NEAR/3 (labor OR birth OR labour OR delivery OR ruptur*)) OR birthweight* OR ((birth OR neonat* OR newborn OR infant*) NEAR/3 (weight*)) OR ((infant* OR child*) NEAR/3 (breast NEXT/1 feed* OR breastfeed* OR milk NEXT/1 composit* OR weaning)) OR ((infant*) NEAR/3 (biomarker* OR saliva OR fecal OR urine OR hair)) OR postpartum OR post NEXT/1 partum OR puerperium OR ((fetal OR foetus OR fetus OR newborn OR neonat* OR infant* OR intrauterin* OR intra NEXT/1 uterin*) NEAR/3 (diseas* OR abnormal* OR growth* OR infect*)) OR ((gestational) NEAR/3 (diabet* OR hypertens* OR weight*)) OR ((placenta*) NEAR/3 (abruption)) OR ((small NEXT/1 for NEXT/1 date OR small NEXT/1 for NEXT/1 gestational NEXT/1 age OR large NEXT/1 for NEXT/1 date OR large NEXT/1 for NEXT/1 gestational NEXT/1 age) NEAR/3 (infant*)) OR preconcept* OR maternal* OR paternal* OR father* OR parent* OR cord NEXT/1 blood* OR fetal NEXT/1 blood* OR lifestyle* OR sociodemographic*):ab,ti,kw) **AND** ((followup* OR follow NEXT/1 up* OR longitudinal* OR prospective* OR retrospective* OR cohort* OR predict*):ab,ti,kw) **AND** ((((obes* OR hypertension* OR diabetes* OR diabetic* OR BMI OR body NEXT/1 mass* OR waist NEXT/1 circum* OR fat NEXT/1 distribut* OR dyslipidemia* OR lipidemia OR lipoedema OR glucos* NEXT/1 intoleranc* OR glucos* NEXT/1 toleranc* OR overweight* OR overnutrition*) NEAR/6 (child* OR progeny OR offspring*)) OR ((metabolic* OR cardiometabolic* OR insulin NEXT/1 resist* OR dysmetabolic* OR metabolism*) NEAR/2 (syndrom* OR diseas* OR outcome* OR disorder*) NEAR/9 (child* OR progeny OR offspring*)) OR ((increas* OR elevat* OR high*) NEAR/2 (bloodpress* OR blood NEXT/1 press*) NEAR/9 (child* OR progeny OR offspring*))):ab,ti,kw)

**Table S1.** Criteria template for quality assessment of the risk factors.

| | **Methodological aspects (excluding study design)** | | **Reflect/mark the study objective** | **Modifiable** | | **Prediction** |
|---|---|---|---|---|---|---|
| **Levels** | **Reproducibility, accuracy, standardization, stability (quality of the sample) and technical variation** | **Biological variation** | **A change in the marker is linked with a change in the endpoint in one or more target population(s)** | **Theoretically modifiable** | **Intervention studies** | |
| Very strong (+++) | Marker is highly reproducible (intra class correlation coefficient >0.9) and accurate (less than 1% deviation from 'correct' value), assay is highly standardized, sample is stable or can easily be made stable | Minimal variation and relevant effects highly superior to variation: effects likely to be observed between groups of tens of individuals | Generally, accepted marker (marker changes consistently linked with a change in the endpoint) | The risk factor is modifiable on an individual level and implementation of modifications in daily life is easy | A systematic review on intervention on this risk factor has been conducted and shows a positive effect of modification on the outcome | >80% of moderate or high quality papers report an association in the same direction and at least 5 publications |
| Strong (++) | Marker is reproducible and accurate enough to detect biological meaningful changes, assay is standardized, sample is stable or can be made stable | High variation explainable (for example, circadian cycle, age, sex, BMI, ethnicity and genotype) and possible to correct it and relevant effects reproducibly superior to variation: effects likely to be observed between groups of fifties to hundreds of individuals | Described as a cause-and effect relationship, but not (yet) generally accepted as a marker, due to a lack of (specific) studies | The risk factor is modifiable on an individual level and implementation of modifications in daily life is complicated | Multiple intervention studies have been conducted showing a potential positive effect of modification on the outcome | >65% of moderate or high quality papers report an association in the same direction and at least 5 publications |
| Medium (+) | Marker is reproducible and accurate enough for specific applications, assay is somewhat standardized and needs to be extensively processed or analysed fast | High variation explainable (for example, circadian cycle, age, sex, BMI, ethnicity and genotype) and possible to correct it and relevant effects reproducibly close to variation: effects may be observed between groups of fifties to hundreds of individuals | Body of evidence suggesting correlation, but cause and effect not established | Modification of the risk factor is not possible on an individual and requires political, governmental, or medical interventions | There is some evidence of potential effect of intervention on the outcome, but literature is controversial | >50% of moderate or high quality report an association in the same direction or less than 5 publications |
| Low (0) | Reproducibility and accuracy of the marker, standardization of the assay and stability of the sample are either poor or not properly documented | High and unexplained variation in a short time span and relevant effects likely to be observed between groups of thousands of individuals | Plausible hypothesis, in use as an exploratory marker, but no substantial body of evidence yet. | The risk factor is not modifiable | No intervention studies have been conducted, or no potential effect of modification has been found | No consistent effect |

| **Table S2.** Associations of childhood obesity with each risk factor per period. | | | | | |
|---|---|---|---|---|---|
| **Author** | **Year** | **Country** | **Exposure (sub)** | **Prevalence Obesity (%)** | **Association** |
| **Preconception** | | | | | |
| **Physical** | | | | | |
| ***Maternal prepregnancy weight/BMI continuous*** | | | | | |
| *Brophy | 2009[24] | UK | Maternal prepregnancy weight | 5.8 | Positive |
| *Chivers | 2012[102] | Australia | Maternal prepregnancy BMI | 7.8 | Positive |
| *Dhana | 2018[17] | USA | BMI | 9.1 | Positive |
| *Fernández-Barrés | 2016[25] | Spain | Prepregnancy BMI | 12.4 | Positive |
| *Fisch | 1975[21] | USA | Maternal height;<br>Maternal weight;<br>Maternal BMI | 5.5 | Positive;<br>Positive;<br>Positive |
| *Gittner | 2013[166] | USA | Prepregnancy BMI | - | Positive |
| Groth | 2017[70] | USA | Prepregnancy BMI | 21.4 | Positive |
| *Huang | 2014[13] | USA | Prepregnancy weight | 26.6 | None |
| *Hunt | 2022[72] | USA | Prepregnancy BMI | 13.1 | Positive |
| *Kjaer | 2019[27] | USA | Prepregnancy BMI | 37.0 | Positive |
| *LaGasse | 2011[61] | USA | Prepregnancy BMI | 21.6 | Positive |
| *Mehta | 2012[50] | USA | Prepregnancy BMI | - | Positive |
| *O'Connor | 2020[31] | USA | Prepregnancy BMI | 17 | Positive |
| *Li | 2011[62] | USA | Prepregnancy weight | 18.0 | Positive |
| Ouyang | 2016[51] | USA | Prepregnancy BMI | 6.4 | Positive |
| *Palma dos Reis | 2022[12] | Portugal | Prepregnancy weight | ? | Positive |
| *Wang | 2022[39] | China | Maternal prepregnancy BMI | 10.8 | Positive |
| *Rotevatn | 2021[54] | Danmark | Prepregnancy BMI | 2.4 | Positive |
| *Ventura | 2020[14] | USA | Prepregnancy BMI | 10.9 | None |
| *Wojcicki | 2015[15] | USA | Maternal prepregnancy BMI; | 24.9 | None |
| *Widen | 2016[76] | USA | Maternal prepregnancy BMI | 22 | Positive |
| Zhang | 2022[192] | China | Maternal prepregnancy weight | ?? | Positive |
| ***Maternal prepregnancy overweight/obesity*** | | | | | |
| *Flores | 2013[91] | USA | Mother severely obese prepregnancy | 5.7 | Positive |
| *Gaillard | 2013[165] | Netherlands | Maternal obesity prepregnancy | 8.1 | Positive |
| Hinkle | 2012[170] | USA | Prepregnancy obesity | 15.8 | Positive |
| *Hu | 2019[94] | USA | Prepregnancy overweight/obesity | 16.6 | Positive |
| *Janjua | 2012[45] | USA | Prepregnancy BMI overweight/obese | 9.6 | Positive |

| | | | | | |
|---|---|---|---|---|---|
| Leonard | 2017[174] | USA | Prepregnancy overweight + excessive weight gain; Prepregnancy obesity + excessive weight gain | 18.0-24.0 | Positive |
| *Navarro | 2020[109] | Ireland | Prepregnancy BMI >30 | 8.8-29.3 | Positive |
| *Reilly | 2005[16] | UK | Maternal BMI >30 kg/m2 | 8.6 | Positive |
| *Whitaker | 1998[189] | USA | Maternal prepregnancy obesity | 20 | Positive |
| ***Paternal prepregnancy overweight/obesity*** | | | | | |
| *Reilly | 2005[16] | UK | Paternal BMI >30 kg/m2 | 8.6 | Positive |
| **Lifestyle** | | | | | |
| ***Smoking*** | | | | | |
| *Dhana | 2018[17] | USA | Smoking | 9.1 | Positive |
| *Sharma | 2008[18] | USA | Smoking | 14.6 | Positive |
| ***Diet*** | | | | | |
| *Dhana | 2018[17] | USA | Healthy diet | 9.1 | None |
| Gete | 2021[19] | Australia | Higher healthy eating index | 3.3 | None |
| ***Physical activity*** | | | | | |
| *Dhana | 2018[17] | USA | Exercise | 9.1 | None |
| Noda | 2022[20] | Japan | Maternal physical activity | 9.5 | None |
| **Pregnancy and birth** | | | | | |
| **Birth anthropometrics** | | | | | |
| ***Birth weight continuous*** | | | | | |
| *Brophy | 2009[24] | UK | Birth weight | 5.8 | Positive |
| *Donkor | 2017[103] | Norway | Birth weight | 3.5 | None |
| *Fisch | 1975[21] | USA | Birth weight | 5.5 | Positive |
| *Fernández-Barrés | 2016[25] | Spain | Birth weight | 12.4 | Positive |
| *Gillman | 2003[26] | USA | Birth weight | 6.7 | Positive |
| *Heerman | 2019[64] | USA | Birth weight | 34.5 | None |
| *Kjaer | 2019[27] | USA | Birth weight | 37.0 | Positive |
| *Layte | 2014[28] | Ireland | Birth weight | ? | Positive |
| *Mardones | 2008[30] | Chile | Birth weight | 17.9 | Positive |
| Mardones | 2014[29] | Chile | Birth weight | 16.0 | Positive |
| *Monteiro | 2003[67] | Brazil | Birth weight | 7.7 | None |
| *O'Connor | 2020[31] | USA | Birth weight | 17 | Positive |
| Parker | 2012[32] | USA | Birth weight | 9 | Positive |
| *Reilly | 2005[16] | UK | Birth weight | 8.6 | Positive |
| *Roy | 2015[154] | USA | Birth weight | 17 | None |
| *Shankaran | 2010[33] | USA | Birth weight | 21 | Positive |
| *Shehadeh | 2008[131] | Israel | Birth weight | - | None |

| | | | | | |
|---|---|---|---|---|---|
| *Shi | 2013[34] | Canada | Birth weight | 13 | Positive |
| *Simpson | 2017[122] | UK | Birth weight | 4.0-6.4 | None |
| *Skledar | 2015[35] | Croatia | Birth weight | 17.8 | Positive |
| *Taveras | 2009[181] | USA | Birth weight for length score | 9 | None |
| *Turner | 2021[36] | UK | Birth weight | 4 | Positive |
| Vehapoglu | 2017[37] | Turkey | Birth weight | 7.8 | Positive |
| *Ventura | 2020[14] | USA | Birth weight | 10.9 | Positive |
| *Von Kries | 2002[38] | Germany | Birth weight | 2.2-8.5 | Positive |
| *Wang | 2022[39] | China | Birth weight | 10.8 | Positive |
| Zarrati | 2013[59] | Iran | Birth weight | 5.3 | None |
| *Zhou | 2011[40] | China | Birth weight | 3.5 | Positive |
| ***Birth length continuous*** | | | | | |
| *Donkor | 2017[103] | Norway | Birth length | 3.5 | None |
| *Mardones | 2008[30] | Chile | Birth length | 17.9 | Negative |
| Mardones | 2014[29] | Chile | Birth length | 16.0 | None |
| *Shehadeh | 2008[131] | Israel | Birth length | - | None |
| *Skledar | 2015[35] | Croatia | Birth length | 17.8 | Positive |
| ***Low birth weight/small-for-gestational age*** | | | | | |
| Chen | 2019b[57] | China | Low birth weight (<2500 g) | 8.0 | Positive |
| *Chiasson | 2016[41] | USA | Low birth weight | 9.9 | Negative |
| De Sousa | 2013[42] | Brazil | Low birth weight | 22.0 | None |
| *Eid | 1970[60] | UK | Low birth weight | 7.7 | Negative |
| Gallo | 2016[58] | Italy | Small-for-gestational age | 12.7-29.1 | Positive |
| *Goodell | 2009[152] | USA | Low birth weight | 18.0 | None |
| Hack | 2014[169] | USA | Extreme low birth weight | 12.0-21.0 | None |
| *Huang | 2014[13] | USA | Low birth weight <2500 g | 26.6 | Negative |
| Kapral | 2018[46] | USA | Low birth weight <2500 g | 12.0-6.5 | None |
| *LaGasse | 2011[61] | USA | Small-for-gestational age | 21.6 | Negative |
| *Lawrence | 2016[48] | USA | Low birth weight | 14.3-36.4 | None |
| *Li | 2011[62] | USA | Low birth weight | 18.0 | Negative |
| *Monteiro | 2003[67] | Brazil | Small-for-gestational age (<P10) | 7.7 | None |
| *Ochoa | 2007[148] | Spain | Birth weight (<2800 g) | - | None |
| Ouyang | 2016[51] | USA | Small-for-gestational age | 6.4 | Negative |
| *Reynolds | 2014[100] | Ireland | Birth weight (<2500 g versus ≥2500 g) | 6.6 | None |
| *Rooney | 2011[53] | USA | Birth weight < 7pounds | 22.6-29.6 | None |
| *Rotevatn | 2021[54] | Denmark | Small-for-gestational age | 2.4 | Negative |
| *Sartorius | 2020[55] | South Africa | Birth weight <2500 g | 15.1 | Negative |
| *Shankaran | 2010[33] | USA | Intra-uterine growth restriction | 21 | Positive |

| | | | | | |
|---|---|---|---|---|---|
| Zarrati | 2013[59] | Iran | Low birth weight (<2500 g) | 5.3 | Positive |
| ***High birth weight/large-for-gestational age*** | | | | | |
| *Chiasson | 2016[41] | USA | High birth weight (>4000 g) | 9.9 | Positive |
| De Sousa | 2013[42] | Brazil | High birth weight | 22.0 | Positive |
| Gu | 2012[43] | China | Macrosomia (>4000 g) | 11.4 | Positive |
| *He | 2000[44] | China | Birth weight >4000 g | - | Positive |
| *Huang | 2014[13] | USA | High birth weight >4000 g | 26.6 | None |
| *Huus | 2007[22] | Sweden | High birth BMI (>95th percentile) | 4.3 | Positive |
| *Janjua | 2012[45] | USA | High birth weight (>85th percentile) | 9.6 | Positive |
| Kapral | 2018[46] | USA | High birth weight >4500 g | 12.0-6.5 | Positive |
| *Kato | 2014[47] | Japan | Obesity at birth | 5.6-8.4 | Positive |
| *Lawrence | 2016[48] | USA | High birth weight | 14.3-36.4 | Positive |
| *Loaiza | 2011[49] | Chile | Macrosomia (>4000 g) | 14.6 | Positive |
| *Mehta | 2012[50] | USA | Large-for-gestational age | - | Positive |
| Ouyang | 2016[51] | USA | Large-for-gestational age | 6.4 | Positive |
| Pan | 2019[52] | China | Macrosomia | 5.4 | Positive |
| *Rooney | 2011[53] | USA | Birth weight > 8.5 pounds | 22.6-29.6 | Positive |
| *Rotevatn | 2021[54] | Denmark | Large-for-gestational age | 2.4 | Positive |
| *Sartorius | 2020[55] | South Africa | Birth weight >4000 g | 15.1 | Positive |
| *Toschke | 2002[56] | Czech Republic | Birth weight > 4000 g | 3.8 | Positive |
| ***Gestational age at birth continuous*** | | | | | |
| *Chen | 2017[63] | Taiwan | Gestational age | 14.7 | None |
| *Heerman | 2019[64] | USA | Gestational age at birth | 34.5 | None |
| *McCrory | 2012[65] | Ireland | Gestational age | 6.6 | None |
| *O'Connor | 2020[31] | USA | Gestational age | 17 | None |
| *Ventura | 2020[14] | USA | Gestational age | 10.9 | None |
| *Mardones | 2008[30] | Chile | Gestational age | 17.9 | Negative |
| ***Preterm birth*** | | | | | |
| Alves | 2016[66] | Brazil | Preterm birth | 12.7 | None |
| *Layte | 2014[28] | Ireland | Gestational age <33 wks;<br>Gestational age <36 wks | ? | Positive;<br>None; |
| *Monteiro | 2003[67] | Brazil | Gestational age (<37 wks) | 7.7 | None |
| *Reynolds | 2014[100] | Ireland | Preterm birth (only ≤32 wks) | 6.6 | Negative |
| *Seipel | 2013[68] | USA | Preterm birth | 11-13 | None |
| *Shankaran | 2010[33] | USA | Preterm birth | 21 | Positive |
| ***Late-term birth*** | | | | | |
| *Chen | 2017[63] | Taiwan | Late-term ≥ 42 wks | 14.7 | Positive |
| *Layte | 2014[28] | Ireland | Gestational age >42 wks | ? | None |

| | | | | | |
|---|---|---|---|---|---|
| ***Foetal growth*** | | | | | |
| Parker | 2012[32] | USA | Estimated foetal growth | 9 | Positive |
| *Turner | 2021[36] | UK | Estimated foetal growth | 4 | Positive |
| ***Placental weight*** | | | | | |
| Ouyang | 2016[51] | USA | Placental weight | 6.4 | Positive |
| **Pregnancy complications** | | | | | |
| ***Mode of Delivery (CS)*** | | | | | |
| Carrillo-Larco | 2015[11] | Peru | Mode of delivery (CS) | 0.7 – 1.1 | Positive |
| *Chen | 2017[63] | Taiwan | Mode of delivery (CS) | 14.7 | Positive |
| Dal'Maso | 2022[77] | Brazil | Mode of delivery (CS) | 8.3 | Positive |
| Flemming | 2013[87] | Canada | Mode of delivery (CS) | 9.8 | None |
| Goldani | 2013[78] | Brazil | Mode of delivery (CS);<br>Mode of delivery (CS) | 13.0/2.1 | Positive;<br>None (two cohorts) |
| *Hawkins | 2019[79] | USA | Mode of delivery (CS) | 12.4 – 13.2 | Positive |
| *Huang | 2014[13] | USA | Mode of delivery (CS) | 26.6 | None |
| Huh | 2012[80] | USA | Mode of delivery (CS) | 9.2 | Positive |
| *Lavin | 2018[81] | Vietnam | Mode of delivery (CS) | 4.1 | Positive |
| Masukume | 2019a[88] | New Zealand | Mode of delivery (CS) | 8.6 | None |
| Mueller | 2015[82] | USA | Mode of delivery (CS) | 25.2 | Positive |
| Pei | 2014[83] | Germany | Mode of delivery (CS) | 11 | Positive (only 2 y) |
| Ralphs | 2021[89] | UK | Mode of delivery (CS) | 5.2 | None |
| Rifas-Shiman | 2021[90] | Belarus | Mode of delivery (CS) | 3.8 – 5.9 | None |
| *Rooney | 2011[53] | USA | Mode of delivery (CS) | 22.6-29.6 | Positive |
| *Rotevatn | 2021[54] | Danmark | Mode of delivery (CS) | 2.4 | Positive |
| *Seipel | 2013[68] | USA | Mode of delivery (CS) | 11-13 | None |
| Si | 2022[84] | China | Mode of delivery (CS) | 3.8 | Positive |
| Sitarik | 2020[85] | USA | Mode of delivery (planned CS) | 20.3 | Positive |
| Yuan | 2016[86] | USA | Mode of delivery (CS) | 13 | Positive |
| *Zhou | 2011[40] | China | Mode of delivery (CS) | 3.5 | None |
| ***Gestational diabetes*** | | | | | |
| *Chen | 2017[63] | Taiwan | Gestational diabetes | 14.7 | Positive |
| *Fernández-Barrés | 2016[25] | Spain | Gestational diabetes | 12.4 | None |
| *Flores | 2013[91] | USA | Gestational diabetes | 5.7 | Positive |
| *Gillman | 2003[26] | USA | Gestational diabetes | 6.7 | None |
| Hakanen | 2016[92] | Finland | Gestational diabetes | 3.0-12.8 | Positive |
| *Hawkins | 2019[79] | USA | Gestational diabetes | 12.4 – 13.2 | Positive |
| Herath | 2020[93] | Sri Lanka | Hyperglycaemia in pregnancy | 3.2 – 5.7 | Positive |
| *Hu | 2019[94] | USA | Gestational diabetes | 16.6 | Positive |
| Lowe | 2018[95] | USA | Gestational diabetes | 8.2 | Positive |

| | | | | | |
|---|---|---|---|---|---|
| Lowe | 2019[96] | Thailand; Barbados; USA; UK; China; Israel; Canada | Fasting glucose;<br>1 h glucose;<br>2 h glucose;<br>HbA1c | 12.3 | Positive;<br>Positive;<br>Positive;<br>Positive |
| *Mehta | 2012[50] | USA | Gestational diabetes | ? | None |
| Nehring | 2013[97] | Germany | Gestational diabetes | 2.6 | Positive |
| Ouyang | 2016[51] | USA | Gestational diabetes | 6.4 | Positive |
| Pettitt | 1983[98] | USA | Gestational diabetes | - | Positive |
| Pettitt | 1998[99] | USA | Glucose during pregnancy | - | Positive (at 5-9 y) |
| Pitchika | 2018[179] | USA, Finland, Germany, Sweden | Gestational diabetes;<br>Diabetes type 1<br>Diabetes type 2 | 5.7 | None;<br>Negative (only 6 y)<br>None |
| *Roy | 2015[154] | USA | Gestational diabetes | 17 | None |
| Thaware | 2015[182] | UK | Gestational diabetes | 12.7 | None |
| *Whitaker | 1998[189] | USA | Gestational Diabetes | 20 | None |
| Wright | 2009[190] | USA | Gestational Diabetes | 9.3 | None |
| Wroblewska-Senuik | 2009[191] | Poland | Gestational Diabetes | 13 | None |
| ***Preeclampsia*** | | | | | |
| *Palma dos Reis | 2022[12] | Portugal | Preeclampsia | ? | None |
| ***Infection*** | | | | | |
| Li | 2020[176] | USA | Infection without antibiotic use vs control | 16.2-21.1 | Positive |
| ***Antibiotic use*** | | | | | |
| Li | 2020[176] | USA | Antibiotic use (with or without infection) | 16.2-21.1 | None |
| Margetaki | 2022[178] | Greece | Prenatal antibiotics use | 7.8-12.0 | None (4 y);<br>Positive (6 y) |
| Mueller | 2015[82] | USA | Use of antibiotics | 25.2 | Positive |
| Sakurai | 2022[180] | Japan | Antibiotic use (in 2nd or 3rd trimester) | 9.4 | Positive (only in females) |
| Wang | 2018[186] | USA | Antibiotic use | 8.9 (4y)<br>4.6 (7y) | None (4y)<br>Positive (7y, 2nd trimester) |
| **Cord blood biomarkers** | | | | | |
| *Donahue | 2011[114] | USA | Higher DHA+EPA/n-3 fatty acids;<br>Higher n-6:n-3 ratio/AA:DHA + EPA ratio/LA:ALA ratio | 9.4 | Negative;<br>Positive |
| *Simpson | 2017[122] | UK | Leptin<br>Adiponectin | 4.0-6.4 | None |
| Sorrow | 2019[123] | USA | Metabolomics | - | Positive/negative |
| **Sociodemographic** | | | | | |
| ***Maternal age*** | | | | | |

| | | | | | |
|---|---|---|---|---|---|
| *Chen | 2017[63] | Taiwan | Age mother at birth | 14.7 | None |
| *Donkor | 2017[103] | Norway | Age mother at birth | 3.5 | None |
| *Fernández-Barrés | 2016[25] | Spain | Maternal age at birth | 12.4 | None |
| *Fisch | 1975[21] | USA | Age mother at birth | 5.5 | Positive |
| *Flores | 2013[91] | USA | Maternal age at birth | 5.7 | None |
| *Gittner | 2013[166] | USA | Age | - | None |
| *Huang | 2014[13] | USA | Age | 26.6 | None |
| *Hunt | 2022[72] | USA | Age | 13.1 | Negative |
| *Huus | 2007[22] | Sweden | Maternal age | 4.3 | None |
| *Kjaer | 2019[27] | USA | Age | 37.0 | None |
| *Layte | 2014[28] | Ireland | Age | ? | None |
| *Seipel | 2013[68] | USA | Maternal age at birth | 11-13 | Positive |
| *Ventura | 2020[14] | USA | Maternal age | 10.9 | None |
| *Wallby | 2017[120] | Sweden | Maternal age | 1.9 | None |
| *Wojcicki | 2015[15] | USA | Maternal age | 24.9 | None |
| ***Paternal age*** | | | | | |
| *Fisch | 1975[21] | USA | Age father at birth | 5.5 | Positive |
| *Huus | 2007[22] | Sweden | Paternal age | 4.3 | None |
| ***Maternal education*** | | | | | |
| *Chivers | 2012[102] | Australia | Education (lower) | 7.8 | Positive |
| *Fernández-Barrés | 2016[25] | Spain | Maternal educational level | 12.4 | None |
| *Huang | 2014[13] | USA | Education (lower) | 26.6 | None |
| *Hunt | 2022[72] | USA | Education | 13.1 | None |
| *Kjaer | 2019[27] | USA | Education | 37.0 | None |
| *Loaiza | 2011[49] | Chile | Finished elementary school | 14.6 | Negative |
| *Mardones | 2008[30] | Chile | Education (>8 years) | 17.9 | Positive |
| *Rotevatn | 2021[54] | Denmark | Maternal education (higher) | 2.4 | Negative |
| *Seipel | 2013[68] | USA | Maternal education (higher) | 11-13 | Negative |
| *Shankaran | 2010[33] | USA | Maternal education (higher) | 21 | Positive |
| *Ventura | 2020[14] | USA | Maternal education (higher) | 10.9 | None |
| *Wallby | 2017[120] | Sweden | Maternal educational level (lower) | 1.9 | Positive |
| *Wen | 2022[121] | USA | Maternal education (lower) | 14.9 | Positive |
| *Wojcicki | 2015[15] | USA | Maternal education (higher) | 24.9 | Positive |
| ***Paternal education*** | | | | | |
| *Wojcicki | 2015[15] | USA | Paternal education (lower) | 24.9 | Positive |
| ***Income*** | | | | | |
| *Rotevatn | 2021[54] | Denmark | Household income (higher) | 2.4 | Negative |
| *Ventura | 2020[14] | USA | Income | 10.9 | None |

| | | | | | |
|---|---|---|---|---|---|
| *Wen | 2022[121] | USA | Family income (lower) | 14.9 | Positive |
| *Wojcicki | 2015[15] | USA | Income (lower) | 24.9 | Positive |
| ***Maternal social economic status*** | | | | | |
| *Fernández-Barrés | 2016[25] | Spain | Social class | 12.4 | None |
| *Fisch | 1975[21] | USA | Socioeconomic index (higher) | 5.5 | None |
| *Ingstrup | 2012[173] | Denmark | Socioeconomic status (lower) | 9.9 | Positive |
| *Layte | 2014[28] | Ireland | Social class | ? | None |
| ***Maternal ethnicity*** | | | | | |
| *Layte | 2014[28] | Ireland | Non-white (African) versus white (Irish, UK, European) | ? | None |
| *Shankaran | 2010[33] | USA | Ethnicity (non-white) | 21 | Negative |
| *Ventura | 2020[14] | USA | Ethnicity (other than white) | 10.9 | None |
| *Wojcicki | 2015[15] | USA | Ethnicity (non-White) | 24.9 | Positive; |
| ***Parity*** | | | | | |
| *Frondelius | 2018[105] | Sweden | Parity | 3.0 | None |
| *Gittner | 2013[166] | USA | Parity | - | Negative |
| *Layte | 2014[28] | Ireland | Parity | ? | None |
| *Rotevatn | 2021[54] | Denmark | Parity | 2.4 | Positive |
| *Ventura | 2020[14] | USA | Parity | 10.9 | None |
| *Wallby | 2017[120] | Sweden | Parity | 1.9 | None |
| *Huus | 2007[22] | Sweden | Parity (any siblings) | 4.3 | None |
| *Lavin | 2018[81] | Vietnam | Multiparous | 4.1 | Negative |
| ***Maternal employment*** | | | | | |
| *Gittner | 2013[166] | USA | Employment (yes) | - | Positive |
| ***Maternal relationship status*** | | | | | |
| *Huus | 2007[22] | Sweden | Maternal relationship status (single) | 4.3 | None |
| *Ventura | 2020[14] | USA | Marital status | 10.9 | None |
| *Wojcicki | 2015[15] | USA | Marital status mother (not married) | 24.9 | Positive |
| **Lifestyle** | | | | | |
| ***Maternal smoking*** | | | | | |
| Chen | 2006[101] | USA | Smoking | 4.3 – 8.6 | Positive |
| *Chivers | 2012[102] | Australia | Smoking | 7.8 | Positive |
| *Donkor | 2017[103] | Norway | Smoking | 3.5 | Positive; |
| Durmus | 2011[104] | Netherlands | Smoking | 2.3 | Positive |
| *Fernández-Barrés | 2016[25] | Spain | Smoking | 12.4 | None |
| *Fisch | 1975[21] | USA | Smoking | 5.5 | None |
| *Frondelius | 2018[105] | Sweden | Smoking | 3.0 | Positive |
| Harris | 2013[106] | USA | Smoking | 18.9 | Positive |

| | | | | | |
|---|---|---|---|---|---|
| *Hawkins | 2019[79] | USA | Smoking | 12.4 – 13.2 | Positive |
| Horiuchi | 2021[23] | Japan | Maternal smoking | - | None |
| *Huang | 2014[13] | USA | Smoking | 26.6 | Positive |
| Ino | 2011[107] | Japan | Smoking | 6.0-16.0 | Positive |
| *Janjua | 2012[45] | USA | Smoking | 9.6 | None |
| *Layte | 2014[28] | Ireland | Smoking | ? | Positive |
| Mizutani | 2007[108] | Japan | Smoking | 2.7 | Positive |
| *Navarro | 2020[109] | Ireland | Smoking | 8.8-29.3 | Positive (for 5y) |
| *O'Connor | 2020[31] | USA | Smoking | 17 | Positive |
| *Palma dos Reis | 2022[12] | Portugal | Smoking | ? | Positive |
| Power | 2002[110] | UK | Smoking | - | Positive (only for 16y) |
| *Reilly | 2005[16] | UK | Smoking | 8.6 | Positive |
| *Reynolds | 2014[100] | Ireland | Smoking | 6.6 | Positive |
| *Rooney | 2011[53] | USA | Smoking | 22.6-29.6 | Positive |
| *Rotevatn | 2021[54] | Danmark | Smoking | 2.4 | Positive |
| *Sharma | 2008[18] | USA | Smoking | 14.6 | Positive |
| *Seipel | 2013[68] | USA | Smoking | 11-13 | Positive |
| *Shankaran | 2010[33] | USA | Smoking | 21 | None |
| *Shi | 2013[34] | Canada | Smoking | 13 | Positive |
| Suzuki | 2009[111] | Japan | Smoking | 3 | Positive |
| *Von Kries | 2002[38] | Germany | Smoking | 2.2-8.5 | Positive |
| Wang | 2014[112] | USA | Smoking | 23.2 | Positive |
| *Wojcicki | 2015[15] | USA | Smoking | 24.9 | None |
| ***Paternal smoking*** | | | | | |
| Horiuchi | 2021[23] | Japan | Paternal smoking | - | None |
| ***Maternal alcohol*** | | | | | |
| *Layte | 2014[28] | Ireland | Alcohol use (light) | ? | Negative |
| *Seipel | 2013[68] | USA | Alcohol use | 11-13 | Positive |
| *Shankaran | 2010[33] | USA | Alcohol use | 21 | None |
| *Huang | 2014[13] | USA | Alcohol use | 26.6 | Negative |
| *LaGasse | 2011[61] | USA | Alcohol | 21.6 | None |
| *Navarro | 2020[109] | Ireland | Alcohol use | 8.8-29.3 | None |
| ***Maternal drugs*** | | | | | |
| *Seipel | 2013[68] | USA | Drugs use | 11-13 | None |
| *Shankaran | 2010[33] | USA | Drugs use | 21 | None |
| *LaGasse | 2011[61] | USA | Cocaine | 21.6 | Positive |
| ***Diet*** | | | | | |
| *Fernández-Barrés | 2016[25] | Spain | Maternal caloric intake<br>Mediterranean diet | 12.4 | None<br>None |

| | | | | | |
|---|---|---|---|---|---|
| *Callanan | 2021[119] | Ireland | Low GI diet | 6.5 | None |
| Chen | 2019a[113] | Ireland | Diet, higher GI;<br>Diet, higher GL (glycaemic load);<br>Diet, insulinemic index (II) | 5.0 | None |
| *Donahue | 2011[114] | USA | Diet, more fish/fatty acid/DHA+EPA intake | 9.4 | Negative |
| Hakola | 2017[115] | Finland | Fatty acid intake | 2.0 - 10.0 | Positive, negative and none |
| Hu | 2020[116] | USA | Diet, fast food pattern;<br>Diet, processed food pattern | 16.4 | Positive;<br>None |
| Kadawathagedara | 2018[117] | Norway | Acrylamide intake | 0.4-1.6 | Positive |
| Klebanoff | 2015[118] | USA | Serum paraxantine | 6.5 | None |
| Mizutani | 2007[108] | Japan | Maternal breakfast habit (skipping) | 2.7 | Positive |
| *Navarro | 2020[109] | Ireland | Maternal diet | 8.8-29.3 | None |
| Suzuki | 2009[111] | Japan | Diet (skipping breakfast) | 3 | Positive |
| ***Physical activity*** | | | | | |
| *Fernández-Barrés | 2016[25] | Spain | Physical activity | 12.4 | None |
| *Navarro | 2020[109] | Ireland | Maternal physical activity | 8.8-29.3 | None |
| ***Stress*** | | | | | |
| Bryl | 2022[162] | Poland | Maternal stress | 5.0 | None |
| *Ingstrup | 2012[173] | Denmark | Maternal distress;<br>Anxious;<br>Depressed;<br>Stressed;<br>Worried;<br>Lack of support | 9.9 | None;<br>None;<br>None;<br>None;<br>Positive;<br>None |
| *Kjaer | 2019[27] | USA | Depression | 37.0 | Negative |
| ***Caffeine*** | | | | | |
| Li | 2015[175] | USA | Caffeine | ? | Positive |
| ***Sleep duration*** | | | | | |
| Mizutani | 2007[108] | Japan | Maternal sleep duration >8h | 2.7 | Negative |
| Suzuki | 2009[111] | Japan | Sleep duration (longer) | 3 | Negative |
| ***Fetal musical education*** | | | | | |
| *Zhou | 2011[40] | China | Fetal musical education (lower) | 3.5 | Positive |
| **Physical** | | | | | |
| ***Paternal BMI or overweight/obesity*** | | | | | |
| *Chivers | 2012[102] | Australia | Paternal BMI at birth | 7.8 | Positive |
| *Whitaker | 1998[189] | USA | Paternal obesity | 20 | Positive |
| ***Maternal BMI or ovesrweight/obesity*** | | | | | |

| | | | | | |
|---|---|---|---|---|---|
| *Lavin | 2018[81] | Vietnam | Maternal overweight/obesity | 4.1 | Positive |
| *Wallby | 2017[120] | Sweden | Maternal BMI;<br>Maternal overweight/obesity; | 1.9 | Positive<br>Positive |
| ***Gestational weight gain continuous*** | | | | | |
| *Chen | 2017[63] | Taiwan | Gestational weight gain | 14.7 | Positive |
| Diesel | 2015b[69] | USA | Gestational weight gain | 21.0 | Positive |
| *Fernández-Barrés | 2016[25] | Spain | Gestational weight gain | 12.4 | None |
| *Fisch | 1975[21] | USA | Gestational weight gain | 5.5 | None |
| Groth | 2017[70] | USA | Gestational weight gain | 21.4 | Positive |
| Hivert | 2016[71] | USA | Gestational weight gain | 12.0 | Positive |
| *Hunt | 2022[72] | USA | Gestational weight gain | 13.1 | Positive |
| *Layte | 2014[28] | Ireland | Gestational weight gain | ? | Positive |
| *Li | 2011[62] | USA | Gestational weight gain | 18.0 | Positive |
| *Seipel | 2013[68] | USA | Gestational weight gain | 11-13 | Positive (only 10y) |
| Oken | 2007[73] | USA | Gestational weight gain | 9 | Positive |
| Oken | 2008[74] | USA | Gestational weight gain | 6.5 | Positive |
| Oken | 2009[75] | USA | Gestational weight gain | 10 | Positive (only women with prepregnancy BMI 25-30) |
| *Wang | 2022[39] | China | Gestational weight gain | 10.8 | Positive |
| *Rooney | 2011[53] | USA | Gestational weight gain | 22.6-29.6 | Positive |
| Widen | 2016[76] | USA | Gestational weight gain | 22 | Positive |
| ***Excessive gestational weight gain*** | | | | | |
| Diesel | 2015a[163] | USA | Excessive gestational weight gain | 16.4 | Positive |
| Ensenauer | 2013[164] | Germany | Excessive gestational weight gain | 2.4 | Positive |
| Hu | 2019[94] | USA | Excessive gestational weight gain | 16.6 | Positive |
| Liu | 2019[177] | USA | Excessive gestational weight gain | 11.0 | None |
| Ouyang | 2016[51] | USA | Gestational weight gain (excessive) | 6.4 | Positive |
| ***Inadequate gestational weight gain*** | | | | | |
| *Liu | 2019[177] | USA | Inadequate gestational weight gain | 11.0 | None |
| *Seipel | 2013[68] | USA | Gestational weight loss | 11-13 | Positive (only at 10y) |
| Ouyang | 2016[51] | USA | Gestational weight gain (inadequate) | 6.4 | Negative |
| ***Maternal lipids (standard pregnancy screening)*** | | | | | |
| Thaware | 2018[183] | UK | Lipids | 11.1 | None |
| **Environmental** | | | | | |
| Guo | 2020a[167] | China | Urinary bisphenol A | 15.8 | Positive |
| Guo | 2020b[168] | China | Polybrominated diphenyl ethers (6 metabolites) | 8.8 | 5 None, 1 negative (BDE-154) |

| | | | | | |
|---|---|---|---|---|---|
| Vafeiadi | 2015[184] | Greece | Persistent organic pollutants | 6.9 | Positive |
| Vafeiadi | 2016[185] | Greece | Bisphenol A exposure | 13.3 | Positive |
| Warner | 2013[187] | USA | Pesticide exposure (Dichlorodiphenyltrichloroethane& Dichlorodiphenyldichloroethylene) | 35.6 | None |
| **Early infancy** | | | | | |
| **Feeding patterns** | | | | | |
| ***Early solid food introduction*** | | | | | |
| *Brophy | 2009[24] | UK | Early solid food introduction (<3 months) | 5.8 | Positive |
| *Gittner | 2013[166] | USA | Timing of solid food introduction | - | None |
| Gooze | 2011[144] | USA | Timing of solid food introduction <3 months | 17.6 | Negative |
| Huh | 2011[171] | USA | Timing of solid food introduction (among breastfed; <4mnth)<br>Timing of solid food introduction (among formulafed; <4mnth) | 9.0 | None<br><br>Positive |
| Moss | 2014[128] | USA | Timing of solid food introduction | 14.3 | None |
| *Reilly | 2005[16] | UK | Introduction to solid food | 8.6 | None |
| *Ventura | 2020[14] | USA | Introduction to complementary foods 4- <6 months<br>Introduction to complementary foods > 6 months | 10.9 | None |
| *Von Kries | 2002[38] | Germany | Introduction to solid foods < 4 months | 2.2-8.5 | None |
| *Zhou | 2011[40] | China | Solid food introduction (<4 mnth) | 3.5 | Positive |
| ***Breastfeeding (yes/no)*** | | | | | |
| *Chiasson | 2016[41] | USA | Breastfeeding (partial or exclusive) | 9.9 | Negative |
| Ehrenthal | 2016[124] | USA | Breastfeeding | 13.4 | Negative |
| *Fernández-Barrés | 2016[25] | Spain | Breastfeeding | 12.4 | None |
| *Gittner | 2013[166] | USA | Breastfeeding | - | None |
| Gooze | 2011[144] | USA | Breastfeeding | 17.6 | Positive |
| *Hawkins | 2019[79] | USA | Breastfeeding | 12.4 – 13.2 | Negative |
| *Heerman | 2019[64] | USA | Breastfeeding | 34.5 | None |
| Hildebrand | 2022[125] | USA | Breastfeeding | 13.0 | Negative |
| *Huang | 2014[13] | USA | Breastfeeding | 26.6 | Negative |
| *Hunt | 2022[72] | USA | Exclusive breastfeeding 6 months vs not exclusively breastfed for 6 months | 13.1 | Negative |
| Huus | 2008[172] | Sweden | Exclusive breastfeeding < 4months vs not exclusively breastfed for 4 months. | 4.3 | None |
| Jing | 2014[146] | China | Breastfeeding | 4.0 | None |
| Jwa | 2014[126] | Japan | Exclusive breastfeeding vs formula feeding | 1.6 – 2.4 | Negative |
| *Kjaer | 2019[27] | USA | Exclusive breastfeeding at 4-6 wks | 37.0 | None |
| *Li | 2011[62] | USA | Breastfeeding | 18.0 | Negative |

| | | | | | |
|---|---|---|---|---|---|
| Mayer-Davis | 2006[138] | USA | Breastfeeding | 6.7 | None |
| Metzger | 2010[127] | USA | Breastfeeding | - | Negative |
| Michels | 2007[147] | USA | Breastfeeding | 21.9 | None |
| Moss | 2014[128] | USA | Breastfeeding | 14.3 | Negative |
| *O'Connor | 2020[31] | USA | Breastfeeding until 6 months (vs none) | 17 | None |
| Park | 2015[129] | Korea | Mixed feeding vs predominantly breast fed | 9.8 | Positive |
| Pattison | 2019[130] | USA | Breastfeeding | 10.0 | Negative |
| *Reilly | 2005[16] | UK | Breastfeeding | 8.6 | None |
| *Seipel | 2013[68] | USA | Breastfeeding | 11-13 | Negative |
| *Shehadeh | 2008[131] | Israel | Breastfeeding (vs combined and formula) | - | Negative |
| *Shi | 2013[34] | Canada | Exclusive breastfeeding for 6 months vs no breastfeeding | 13 | Negative |
| *Toschke | 2002[56] | Czech Republic | Breastfeeding | 3.8 | Negative |
| Van Rossem | 2011[132] | USA | Breastfeeding | 3.4-9.2 | Negative |
| *Von Kries | 2002[38] | Germany | Breastfeeding | 2.2-8.5 | Negative |
| *Wang | 2017[133] | USA | Breastfeeding at 1 month | 18.4 | Negative |
| Yamakawa | 2013[134] | Japan | Partial or exclusive breastfeeding | 2 | Negative |
| ***Breastfeeding (duration)*** | | | | | |
| Burdette | 2007[135] | USA | Breastfeeding > 4months | 18 | Negative |
| *Donkor | 2017[103] | Norway | Breastfeeding duration < 4 months | 3.5 | Positive |
| *Fernández-Barrés | 2016[25] | Spain | Breastfeeding duration | 12.4 | None |
| *Frondelius | 2018[105] | Sweden | Breastfeeding duration | 3.0 | None |
| Gooze | 2011[144] | USA | Breastfeeding duration | 17.6 | None |
| Grube | 2015[136] | Germany | Breastfeeding duration >4 months | 6.4 | Negative |
| *Heerman | 2019[64] | USA | Breastfeeding duration | 34.5 | None |
| Hildebrand | 2022[125] | USA | Breastfeeding duration >6 months | 13.0 | Negative |
| Hummel | 2021[137] | USA; Finland;<br>Germany;<br>Sweden | Any breastfeeding >6 months;<br>Exclusive breastfeeding >3 months | 5.3 | Negative;<br>Negative |
| Izadi | 2013[145] | Iran | Breastfeeding duration | ? | None |
| Jing | 2014[146] | China | Breastfeeding duration | 4.0 | None |
| *Layte | 2014[28] | Ireland | Breastfeeding duration >6 months | ? | Negative |
| Martin | 2013[142] | Belarus | Breastfeeding duration | 5.2 | None |
| Martin | 2017[143] | Belarus | Exclusive Breastfeeding > 3 months | 4.4 | None |
| Mayer-Davis | 2006[138] | USA | Breastfeeding duration > 9 months | 6.7 | Negative |
| *McCrory | 2012[65] | Ireland | Breastfeeding duration >13 weeks | 6.6 | Negative |
| Michels | 2007[147] | USA | Breastfeeding duration | 21.9 | None |
| Morovic | 2019[139] | Croatia | Breastfeeding < 6 months (vs >6 months) | 12.2 | Positive |
| *Ochoa | 2007[148] | Spain | Breast feeding > 3 months | - | None |
| Pattison | 2019[130] | USA | Breastfeeding duration | 10.0 | None |

| | | | | | |
|---|---|---|---|---|---|
| *Reynolds | 2014[100] | Ireland | Breastfeeding >11 weeks | 6.6 | Negative |
| Shields | 2006[149] | Australia | Breastfeeding duration | 4.6-6.9 | None |
| *Skledar | 2015[35] | Croatia | Breastfeeding duration | 17.8 | None |
| Tambalis | 2018[140] | Greece | Breastfeeding duration >1 month | ? | Negative |
| Toschke | 2007[141] | UK | Breastfeeding duration | 6.0 | Negative |
| Van Rossem | 2011[132] | USA | Breastfeeding duration | 3.4-9.2 | Negative |
| *Ventura | 2020[14] | USA | Breastfeeding duration >16 weeks | 10.9 | None |
| *Wallby | 2017[120] | Sweden | Breastfeeding duration | 1.9 | Negative |
| *Wang | 2017[133] | USA | Breastfeeding > 6 months | 18.4 | None |
| *Wojcicki | 2015[15] | USA | Breastfeeding (in months continuously) | 24.9 | Negative |
| ***Milk formula type*** | | | | | |
| Jwa | 2014[126] | Japan | Mixed feeding vs formula feeding | 1.6 – 2.4 | Negative |
| Mayer-Davis | 2006[138] | USA | Breastfeeding; predominantly breast; predominantly formula; formula only | 6.7 | None |
| Weber | 2014[188] | Belgium; Germany; Italy; Poland; Spain | High protein formula vs low-protein formula | 6.4 | Positive |
| **Infant anthropometrics** | | | | | |
| ***BMI/weight*** | | | | | |
| *Huus | 2007[22] | Sweden | High BMI 1 year (>95$^{th}$ percentile) | 4.3 | Positive |
| *Taveras | 2009[181] | USA | Weight for length at 6 months | 9 | Positive |
| ***BMI/weight change*** | | | | | |
| Aris | 2017[150] | Singapore | BMI change | 7.5 | Positive |
| Aris | 2018[151] | USA; Belarus | Weight change | USA: 12.3; Belarus: 5.0 | Positive |
| *Eid | 1970[60] | UK | Excessive postnatal weight gain | 7.7 | Positive |
| *Goodell | 2009[152] | USA | Rapid postnatal weight gain | 18.0 | Positive |
| *Kato | 2014[47] | Japan | Height change; Weight change | 5.6-8.4 | None; Positive |
| *LaGasse | 2011[61] | USA | Early weight gain | 21.6 | Positive |
| *Li | 2011[62] | USA | Rapid infant weight gain | 18.0 | Positive |
| Liu | 2017[153] | USA | Infant BMI trajectory: high as compared to low | 11.0 | Positive |
| *Monteiro | 2003[67] | Brazil | Weight/height gain in first 24 months | 7.7 | Positive |
| *O'Connor | 2020[31] | USA | Weight gain in first 24 months | 17 | Positive |
| *Rooney | 2011[53] | USA | Weight gain in first 4 months | 22.6-29.6 | Positive |
| *Rotevatn | 2021[54] | Danmark | Infant weight gain (rapid) | 2.4 | Positive |
| *Roy | 2015[154] | USA | >= 90$^{th}$ percentile of weight checks | 17 | Positive |
| Smego | 2016[155] | USA | Weight for length in first 24 months | 4.5-12 | Positive |
| *Taveras | 2011[156] | USA | Number of crossed major weight-for-length percentiles | 13.9 | Positive |

| | | | | | |
|---|---|---|---|---|---|
| *Ventura | 2020[14] | USA | Weight for age in first year | 10.9 | Positive |
| ***NICU admission*** | | | | | |
| *Reynolds | 2014[100] | Ireland | NICU admission | 6.6 | None |

**Figure S1-3.** Bias assessment of the included studies.

**Figure 1a**. Risk of Bias assessment. Preconception Case-Control studies

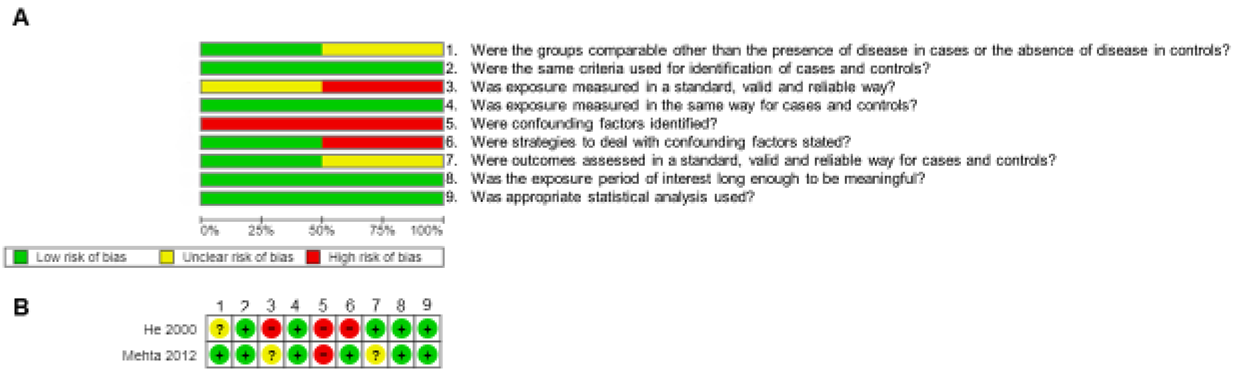

**Figure 1b**. Risk of Bias assessment. Preconception Prospective cohort studies

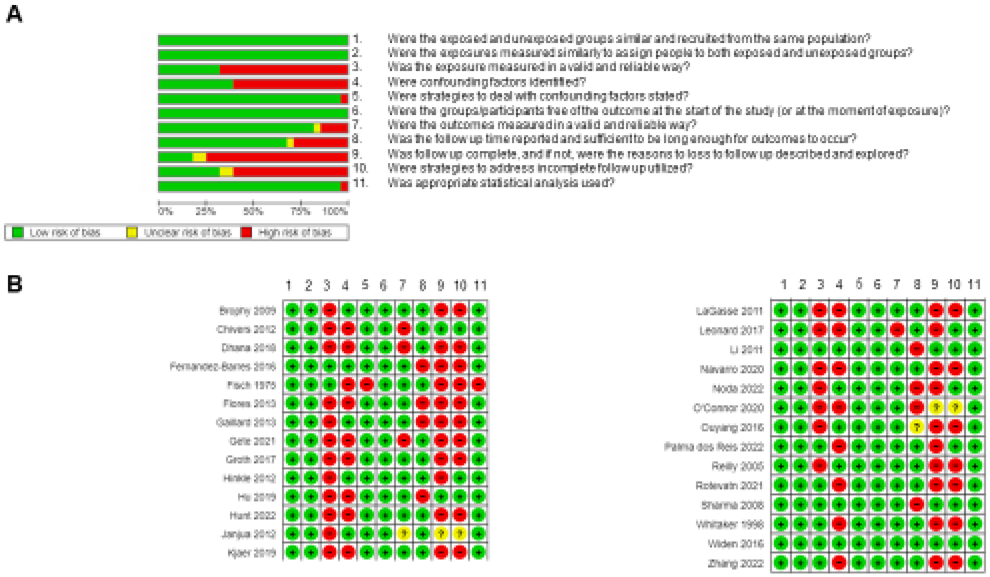

**Figure 1c**. Risk of Assessment. Preconception
Retrospective cohort studies

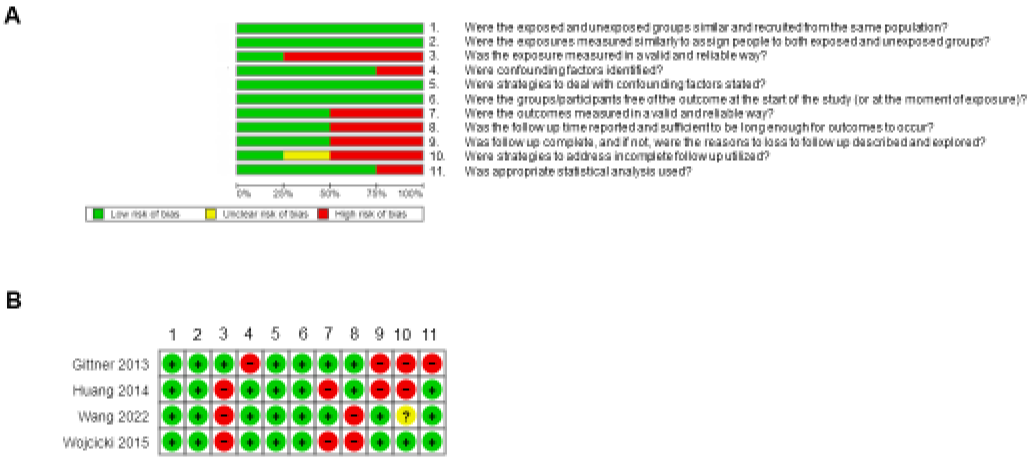

**Figure 2a**. Risk of Bias Assessment. Pregnancy and birth Case-Control studies

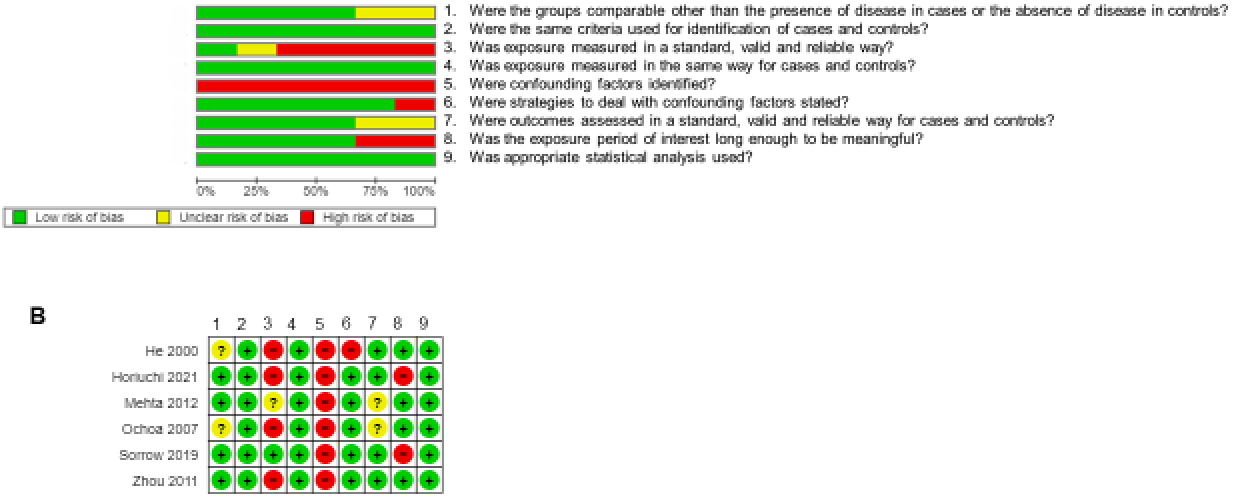

**Figure 2b**. Risk of Bias Assessment.
Pregnancy and birth RCT studies

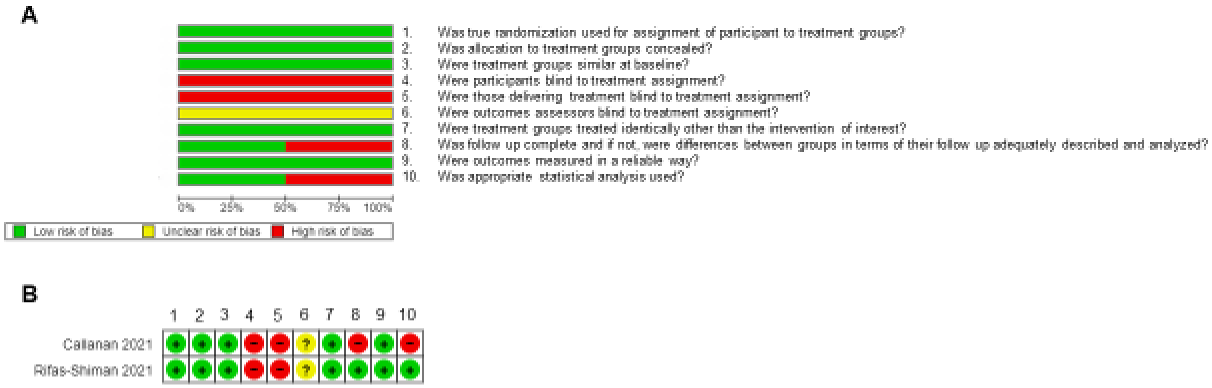

**Figure 2c**. Risk of Bias Assessment. Pregnancy and birth Prospective cohort studies

**A**

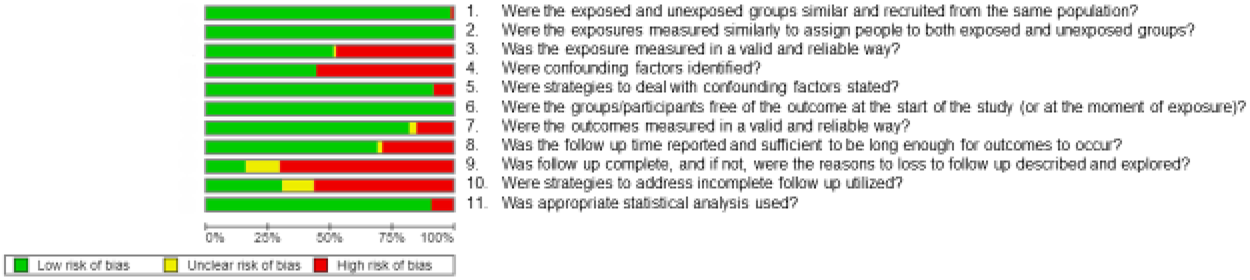

**Figure 2c**. Risk of Bias Assessment. Pregnancy and birth Prospective cohort studies

**CONTINUED**

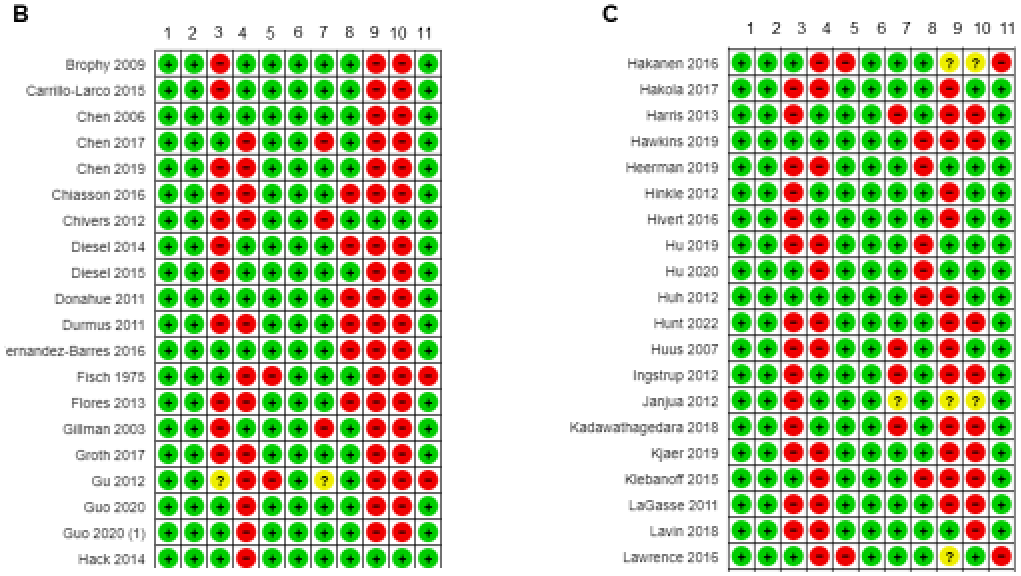

**Figure 2c**. Risk of Bias Assessment. Pregnancy and birth Prospective cohort studies

**CONTINUED**

D

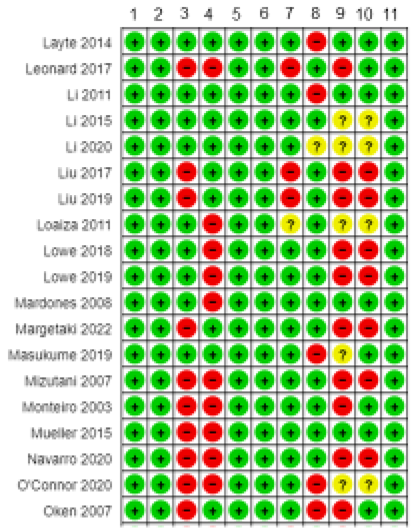


E

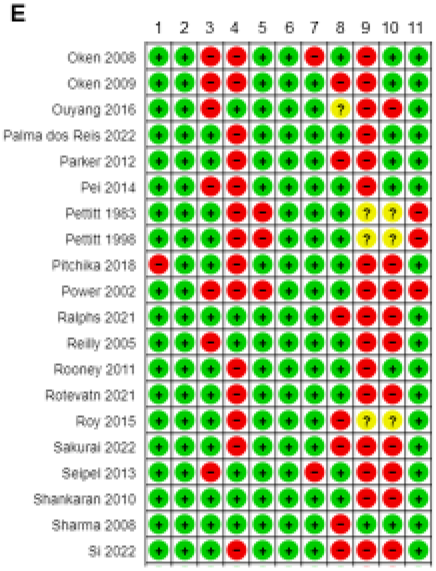

**Figure 2c**. Risk of Bias Assessment. Pregnancy and birth Prospective cohort studies

**CONTINUED**

F

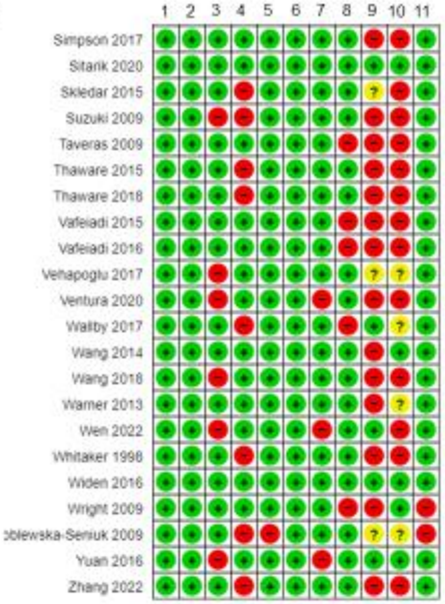

**Figure 2d**. Risk of Bias Assessment. Pregnancy and birth Retrospective cohort studies

**A**

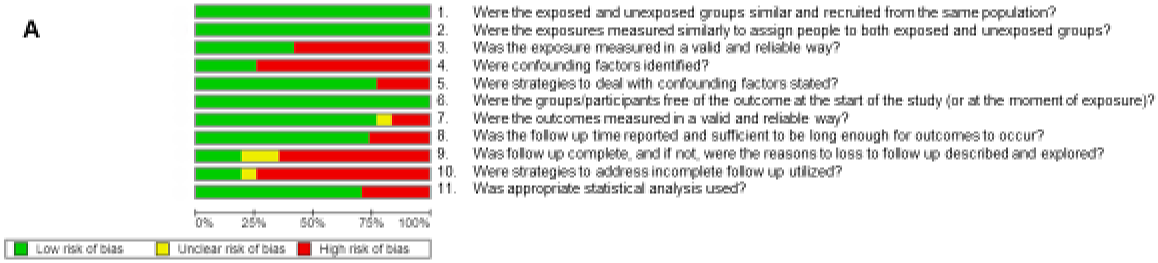

**Figure 2e**. Risk of Bias Assessment. Pregnancy and birth Retrospective cohort studies

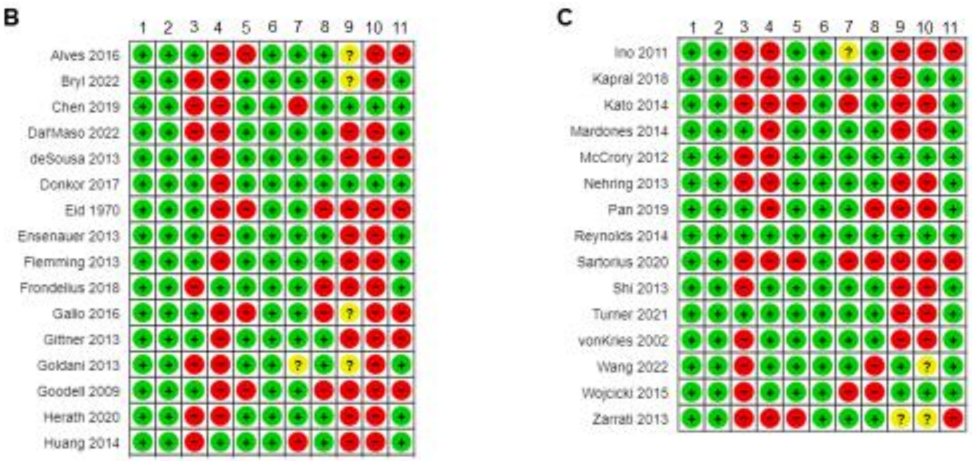

**Figure 3a**. Risk of Bias Assessment. Early Infancy Case-Control studies

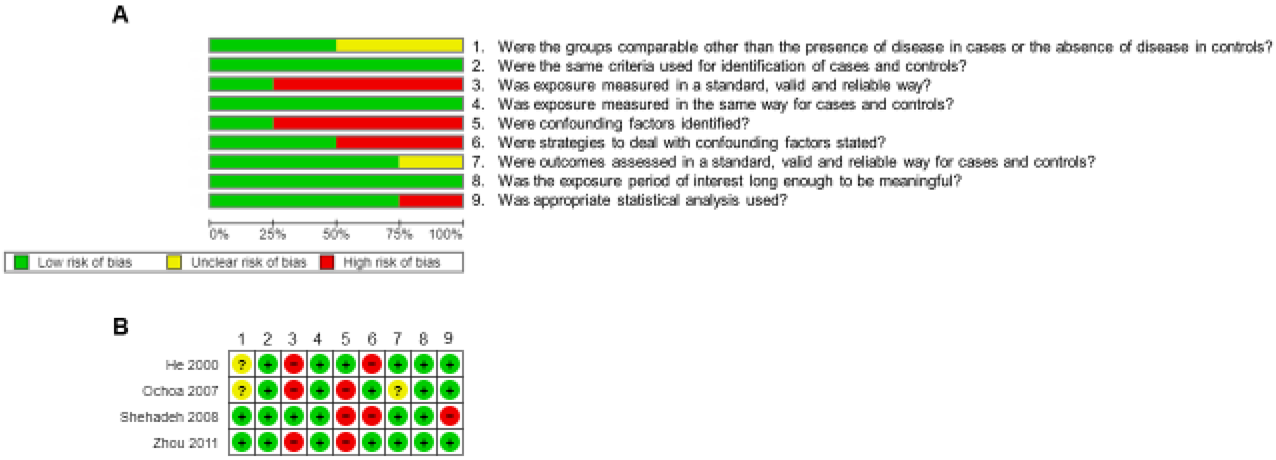

**Figure 3b**. Risk of Bias Assessment. Early Infancy RCT studies

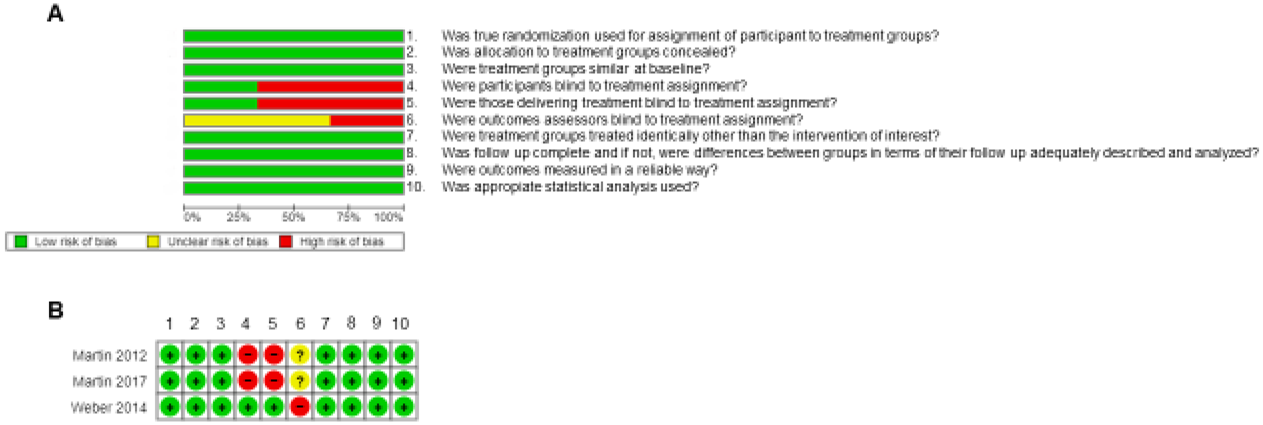

**Figure 3c**. Risk of Bias Assessment. Early Infancy Prospective cohort studies

**CONTINUED**

**A**

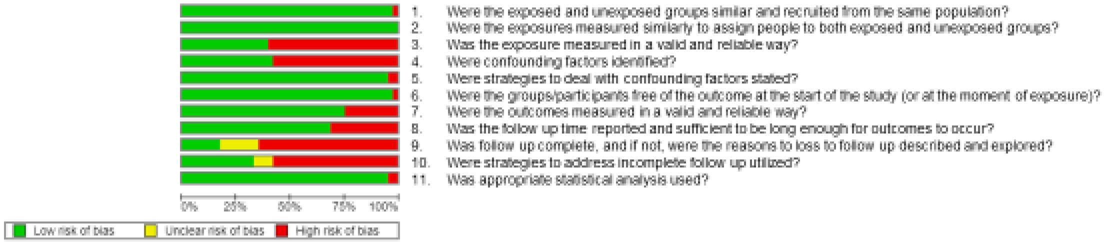

**Figure 3c**. Risk of Bias Assessment. Early Infancy Prospective cohort studies

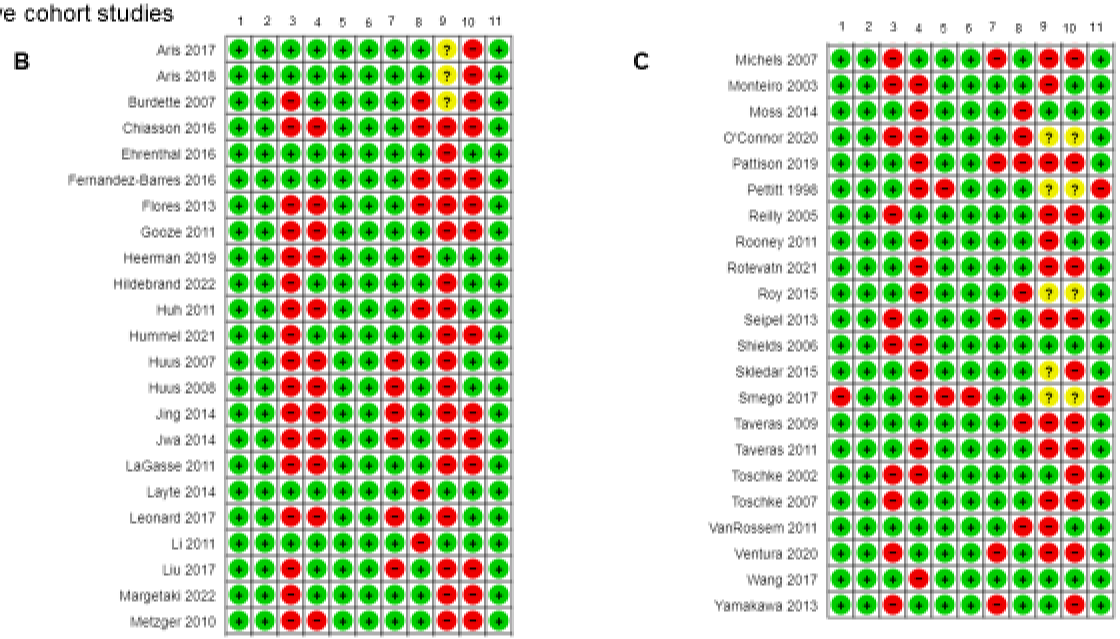

**Figure 3d**. Risk of Bias graph and summary of Early Infancy Retrospective cohort studies

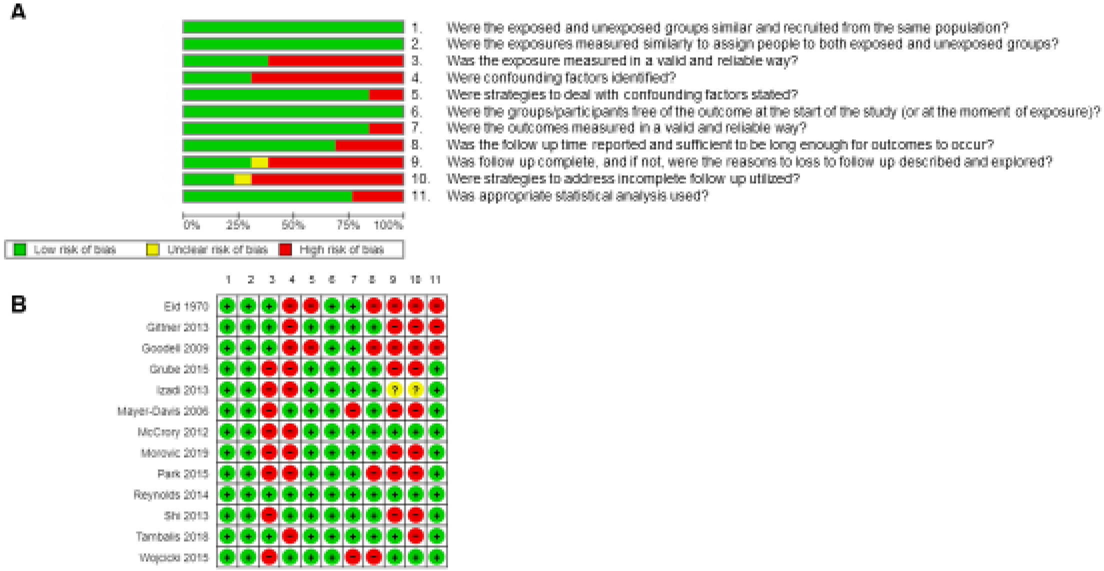

**Figure S4. Quality assessment of the 24 consistently associated risk factors with childhood obesity***

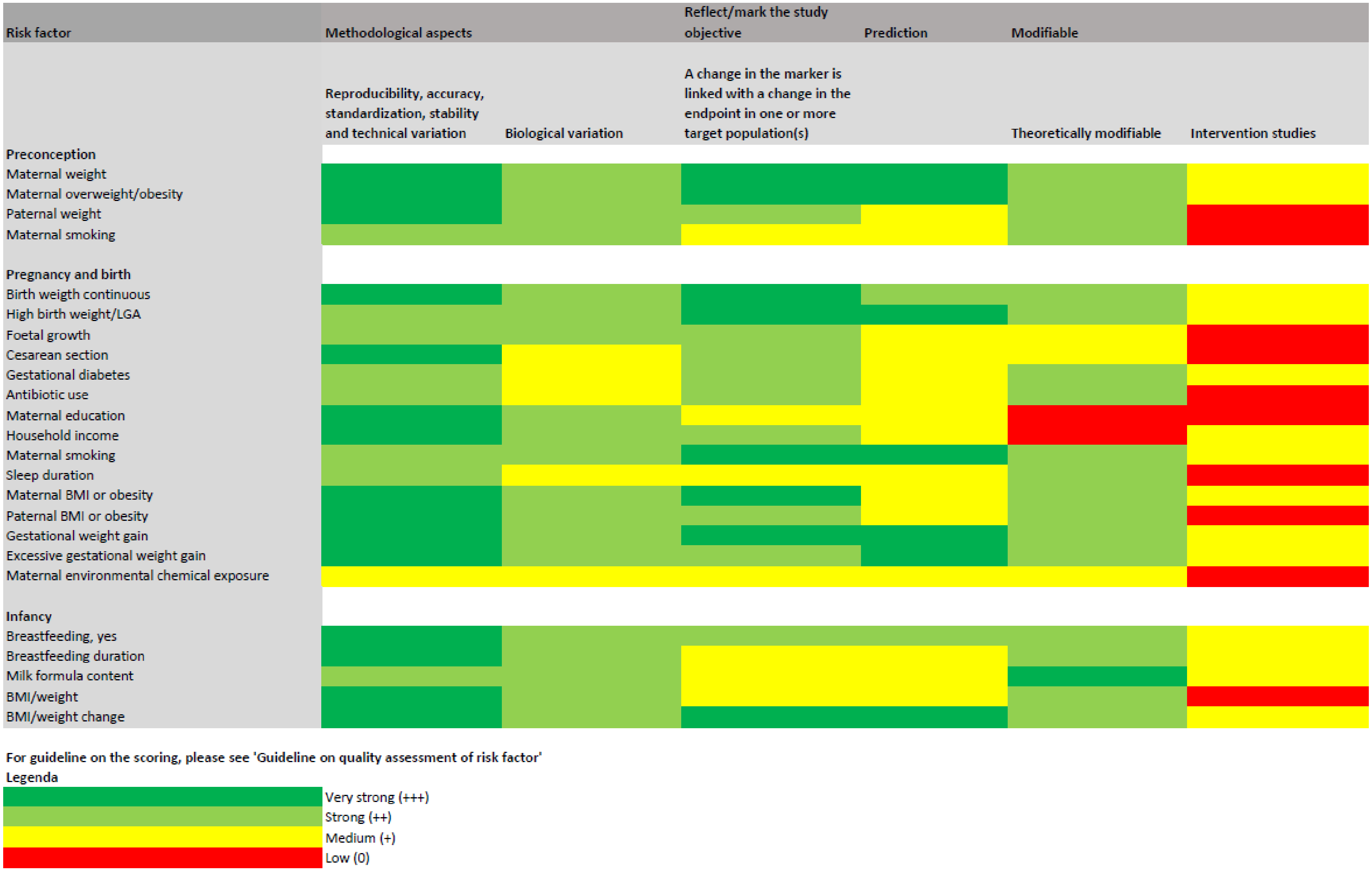


*Scores based on table S1
